# How Have Global Scientists Responded to Tackling COVID-19?

**DOI:** 10.1101/2022.08.16.22278871

**Authors:** Longbing Cao, Wenfeng Hou

## Abstract

Since the outbreak of COVID-19, the global scientific communities across almost all countries have made urgent, intensive, and continuous effort on understanding, fighting and modeling the COVID-19 pandemic. COVID-19 research turns out to be the first overwhelming global scientific reaction to significant global crises and threats. This literature analysis report collects and summarizes the profiles, trends, quality and impact of this global scientific response. It collects and analyzes 346,267 scientific references in English, involving researchers from 189 countries and regions in 27 subject areas. The report generates a picture of how global scientists have responded to COVID-19 between Jan 2020 and Mar 2022 in terms of their publication quantity, impact, focused major problems, and research areas and methods over country/region, discipline and time and collaboratively. The report also captures broad-reaching distributions and trends of modeling COVID-19 by AI, data science, analytics, shallow and deep machine learning, epidemic modeling, applied mathematics, and social science methods, etc. We further show the correlations between publication quality and quantity and economic status and COVID-19 infections globally and in major countries and regions. The literature analysis results of this global scientific response to COVID-19 present a comprehensive global, regional and subject-specific picture of the significant cross-disciplinary, cross-country, cross-problem, and cross-technique profiles and differences of the COVID-19 publication quantity and quality. The report also discloses significant imbalances in the COVID-19 research across countries/regions, subject areas, problems, topics, methods, research collaborations, and economic statuses. We share the source and analytical data of this global literature analysis for further research on this unprecedented and future crises.

**More Information about COVID-19 Modeling:** *The COVID-19 global scientists response dataset:* The COVID-19 global scientists response dataset supporting this report can be found in Kaggle at: https://www.kaggle.com/datasets/datascienceslab/covid19-global-scientists-response, Dataset doi: 10.34740/kaggle/dsv/4077649.

*More information about COVID-19 modeling:* Interested readers may refer to the following review on modeling COVID-19 which complements this global literature analysis: Longbing Cao and Qing Liu, COVID-19 Modeling: A Review, 1-103, 2022 (version 2). Accessible at medRxiv: https://www.medrxiv.org/content/10.1101/2022.08.22.22279022v1. More information about COVID-19 modeling, including the analytical results of this report and data and review on COVID-19 can be found at: https://datasciences.org/covid19-modeling.

*Citation of this report:* Longbing Cao and Wenfeng Hou. How have global scientists responded to tackling COVID-19? Full technical report, https://doi.org/10.1101/2022.08.16.22278871, Data Science Lab, University of Technology Sydney, 2022.

## 1 Introduction

The COVID-19[**Elsken19**] pandemic^1^^2^ has caused unprecedented impact on almost every aspect of our life, work, study, travel, and entertainment and has reshaped our economy, society, and globalization. Perhaps, for the first time in the human scientific history, almost all countries, institutions, disciplines and scientists globally have been involved more or less in reacting to this emergent, unprecedented, significant threat and crisis to humans in a very unprecedented, immediate, intensive and continuous manner. An important research agenda is to review this mostly intensive global scientific effort ([**Cao-Liu’22**]) and to create a COVID-19 publishing profile of this globalized COVID-19 research. In particular, as quantifying COVID-19 is essential for understanding and managing the pandemic, it is important to understand what modeling techniques including in mathematics, epidemiology, AI, data science, machine learning, deep learning, and social science etc. have been applied and where they have played their part in modeling COVID-19^3^ ([**Cao-Liu’22, Cao’22**]).

### 1.1 Review objectives and questions

Accordingly, we set up four major review objectives in this project to understand how global scientists have responded to fighting the COVID-19 pandemic. By collecting and analyzing all literature on COVID-19, our first objective is to draw a global COVID-19 publishing picture of their quantity, quality (in terms of publication impact metrics), mainly concerned problems, and mostly applied techniques, etc. The second objective is to understand the differences between countries, disciplines, problems, and techniques in both publishing quantity and quality. The third objective is to explore how major countries including the US, China, G20 and OECD countries and regions perform and differ from each other in terms of their research interests and publication impact. Lastly, we aim to explore the correlation between COVID-19 publication and GDP per capita of a country and between publication and infected and death case numbers in major countries. We then design various research questions for this exploration.

First, to generate an overview of the global COVID-19 research quantity and quality, we aim to identify answers from the literature analysis to the following questions.

- What is the global statistics of the COVID-19 publication quantity and quality (impact)?
- What is the global statistics of the COVID-19 publication quality across major disciplines?
- How has each country published on COVID-19 quantitatively and what is their impact in terms of major publication impact metrics?
- What are the mostly concerned keywords globally?
- What is the global distribution of the cumulative publication impact in terms of combining major publication impact metrics?
- What is the global publication impact distribution in terms of various impact metrics?
- Who are the top-10 published countries and their research impact?
- What is the disciplinary research impact of the top-10 published countries?
- How have global scientists collaborate with each other in fighting the pandemic?

These are expected to capture the overall profile and trends of this global scientific exploration and provide statistics about how the COVID-19 publication quantity and quality differ between countries and their high-performing countries in studying the COVID-19.

Second, to quantify how G20 and OECD countries and regions have performed in tackling COVID-19, we explore the indication to the following questions:

- What are the publication number and impact of G20 countries and regions?
- What is the disciplinary publication impact of G20 countries and regions?
- What is the publication-averaged mean publication impact of G20 countries and regions?
- What are the publication number and mean publication impact of OECD countries and regions?

Third, we aim to understand how various modeling techniques ([**Cao-Liu’22**]), including applied mathematics, AI and data science (including shallow and deep learning and analytics) ([**Cao’22**]), epidemic modeling, and other methods such as simulation, have contributed to quantifying COVID-19 in terms of their trending effects. We thus explore the following questions:

- What are the main keywords concerned in modeling COVID-19 globally?
- What are the main modeling keywords concerned in different disciplines?
- What are the main keywords concerned in modeling COVID-19 in the two major economic and technical powers USA and China respectively?
- What are the top-50 concerned problems and their publication impact?
- What are the top-50 concerned problems and their modeling techniques?
- What are the top-50 modeling techniques and their publication impact?
- What are top-10 monthly concerned problems in modeling COVID-19?
- What are the top-10 problems concerned in the top-10 published countries?
- What are the top-10 modeling techniques applied by the top-10 published countries?
- What are the top-10 problems and techniques concerned by the top-10 published countries?

Lastly, we aim to understand the relations between the COVID-19 research outcomes and a country’s coronavirus infections, deaths, and their economy by exploring the following questions:

- How does global first-authored countries’ GDP per capita correlate with their publications and cumulative publication impact?
- How does a G20 country’s GDP correlate with their publication quantity and impact?
- How does an OECD country’s GDP correlate with their publication quantity and impact?
- How does global monthly publications correlate with the global monthly infections and deaths in major disciplines?
- How does a G20 country’s coronavirus infections and deaths correlate with their publications?
- How does an OECD country’s coronavirus infections and deaths correlate with their publications?

### 1.2 Review methods

To obtain answers and indications to the above research questions, we developed crawling programs to acquire, preprocess, and analyze the literature on COVID-19 from major digital libraries and archival systems. Below, we explain how we extracted and processed the COVID-19 publications.

First, the references, with mostly published ones and partially preprints, were collected from the COVID-19 Open Research Dataset (CORD-19) in SemanticScholar from Jan 2020 to Mar 2022, as discussed in Section 2.1. We further collected the supplementary information about the collected publications, including their publishing dates, and author country information. This information is collected and verified through mapping the CORD-19 publications with those listed in Web of Science, Researchgate, WHO, Crossref, medRxiv, arXiv, and the US National Library of Medicine. More details are in Section 2.2. In addition, we collected the publication impact metrics of journals and conferences which have published the collected publications from Google Scholar, Web of Science’s Journal Citation Reports, and Scopus. More details are available in Section 2.3. These materials are matched and processed by programs and verified manually to form the global COVID-19 publication source data.

Then, we preprocessed the publication data including filtering all publications before 2020 and not in English, and extracted country information and publication date available from their publications. More details are in Sections 3.2 and 3.3.

In all publications, we extracted keywords related to COVID-19 problems and techniques of handling such problems from their titles and abstracts by Apache OpenNLP^4^. To extract the COVID-19 modeling keywords of each publication, after generating the embeddings of the extracted keywords and a list of predefined domain-specified keywords on modeling by SciBert, those extracted keywords with highly similar embeddings to that of the domain-specified modeling keywords were selected as the final modeling keywords of each publication. More details are in Section 3.5. To extract the COVID-19 modelled problem keywords of each publication, we applied PositionRank based on Pagerank and domain-specified synonym and expression mapping to extract the modelled problems in each publication. More details are in Section 3.4. Consequently, we obtained the keywords reflecting the COVID-19 modeling techniques and modelled problems of all publications to highlight the publication profile of COVID-19 modeling.

In addition, we categorize each publication to its mostly relevant disciplines by focusing on medical science, computer science (broadly refer to IT, computing, informatics, applied statistics), and social science. The disciplinary categorization is generated by matching each publication’s publishing venue to the disciplinary information available in the Web of Science, SJR^5^, and Research.com^6^. The disciplinary information about social science is mapped to that in Social Science Research Network (SSRN)^7^. We thus obtained the disciplinary categorization of each modeling publication. More details are in Section 3.6.

Lastly, we further extracted all publications with modeling as their focus. This was done by mapping the extracted publication keywords to a list of domain-specified modeling-sensitive words and expressions. More details are in Section 3.7.

As a result, we identified 346,267 publications, including 26,420 preprints. 186,650 are from medical science publications, 13,959 from computer science, and 32,128 from social science. 51,071 publications including 7,150 focus on and are related to general COVID-19 modeling research, where 21,785 are from medical science, 6,142 from computer science, and 3,890 from social science.

### 1.3 Publication analysis methods

Our focus in this report to undertake the publication analysis driven by the aforementioned review questions in terms of four aspects: (1) global publication profile in Chapter 5, (2) G20 and OECD publication profile in Chapter 6, (3) publication profile on modeling COVID-19 in Chapter 7, and (4) the correlations between publications, economy and infections in Chapter 8.

First, we applied and created various evaluation measures to quantify the target variables and their analytical results. We involve major publication impact metrics to quantify the impact of publications. They are H5-index, Impact Factor, CiteScore, SNIP, and SJR, introduced in Section 4.1. We further generated several measures, including the composite indicator in Section 4.2 that integrates H5-index, Impact Factor, CiteScore, SNIP, and SJR, the correlation coefficient between publications and GDP per capita of a country in Section 4.4.

Second, the above four aspects of the COVID-19 publishing profiles are generated by descriptive, contrastive, multi-dimensional, ranking, linkage, sequential, and correlative analytics of the COVID-19 publication quantity, quality and trending effects across countries and regions, disciplines, concerned problems, and applied techniques, in particular on COVID-19 modeling.

- *Descriptive analytics* collects the statistics of publications in terms of factors such as publication number and mean, the statistics of publication impact metrics such as mean, maximum, and minimum values, for example, the statistics and distributions of global publications in Chapter 5.
- *Contrastive analysis* provides comparison between target publication variables, such as first-authored country, publication quantity, publication impact metrics, and publication disciplines, e.g., G20 country’s publication impact in computer science in Section 6.2.
- *Multi-dimensional analysis* presents the results in terms of multiple variables, e.g., producing a joint view of relations between top-50 problems and their publication impact, and between top-10 modeling methods, top-10 problems in top-10 published countries as in Section 7.14, and G20 country’s mean publication impact in Section 6.5.
- *Ranking analysis* is used to generate top-K list in terms of target variables, for example, top-10 published countries in Section 5.10, and top-50 problems in Section 7.8.
- *Linkage analysis* is applied to identify the connectivity between target variables, e.g., the research collaborations between countries in Section 5.12.
- *Sequential analysis* captures temporal patterns and dynamics of a target variable, for example, top-10 monthly concerned problems in modeling COVID-19 in Section 7.11.
- *Correlation analysis* explores the relationships and coefficients between two to multiple factors, for example, the correlations between publications and GDP per capita in G20 countries in Section 8.2.

Lastly, various visualization techniques are applied to present the analytics and their results. They include choropleth world map^8^ (e.g., Figure 2), multi-dimensional visualization^9^ (e.g., Figure 15), radar chart^10^ (e.g., Figure 19), and contour map^11^ (e.g., Figure 28).

**Figure 1:**
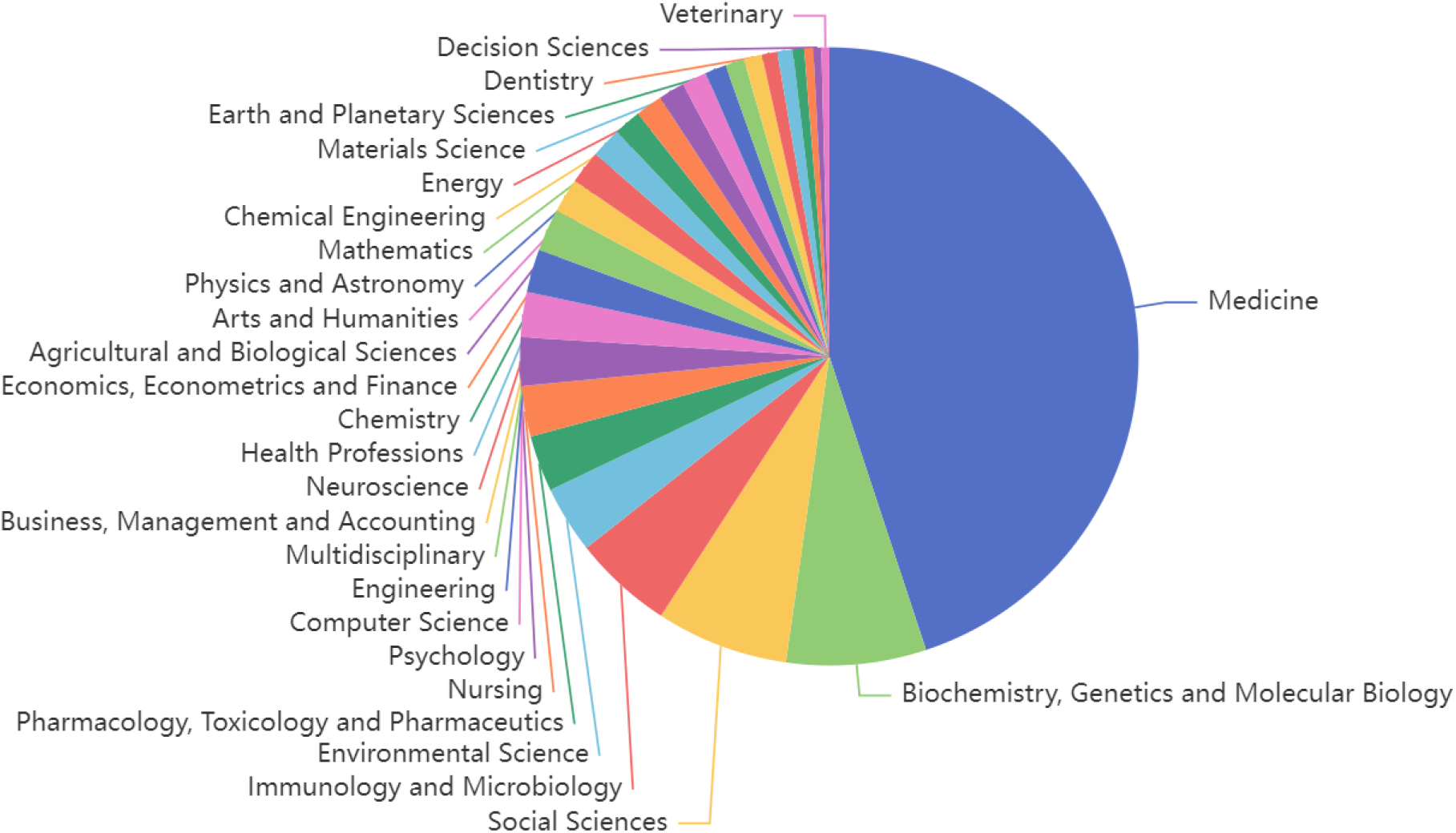
Disciplinary distribution of COVID-19 publications

**Figure 2:**
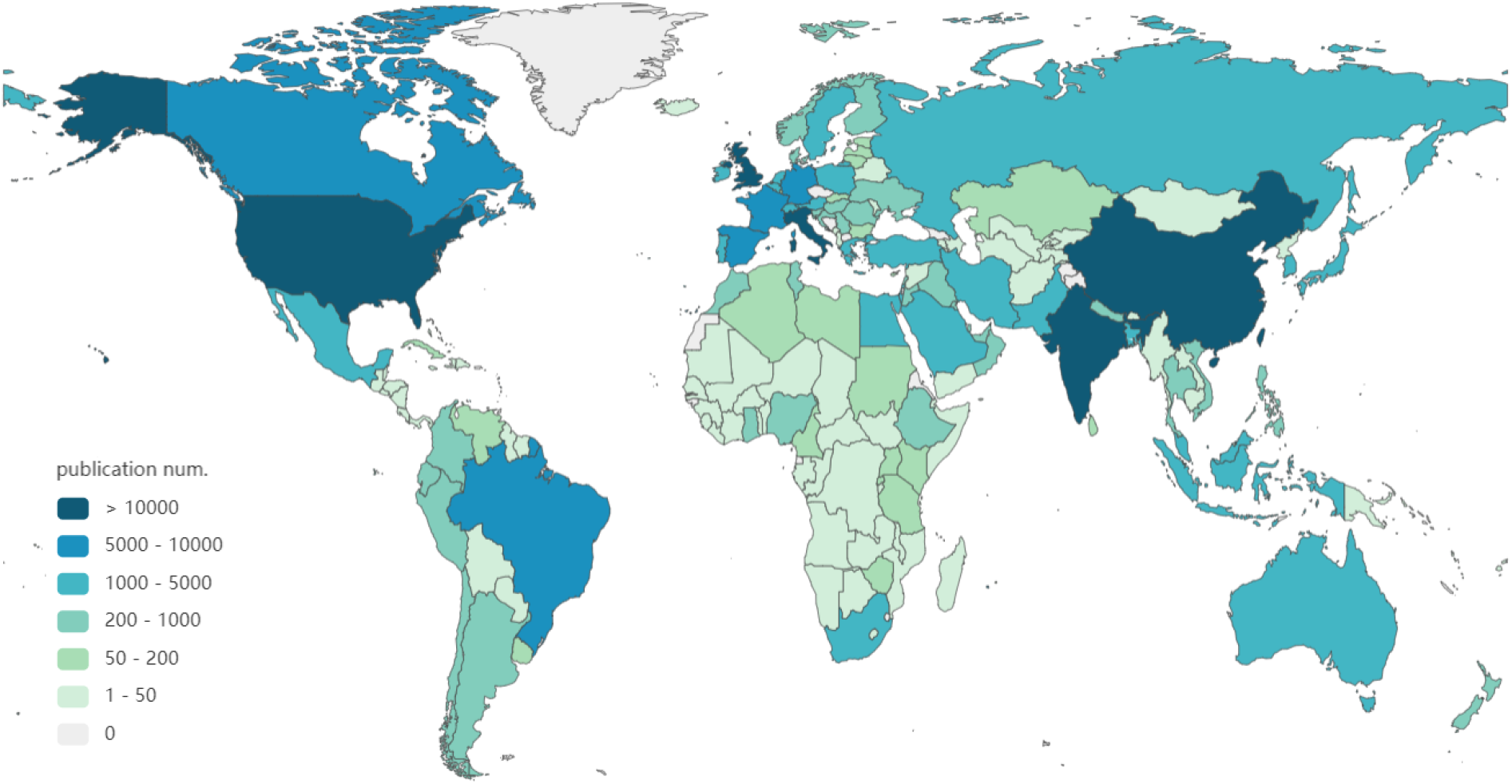
Global publication distribution

In addition to the visualization tools applied above, we present the results in Python or R programming. These include: word cloud (e.g., Figure 3), linkage analysis (e.g., Figure 12), and heat map (e.g., Figure 27)

**Figure 3:**
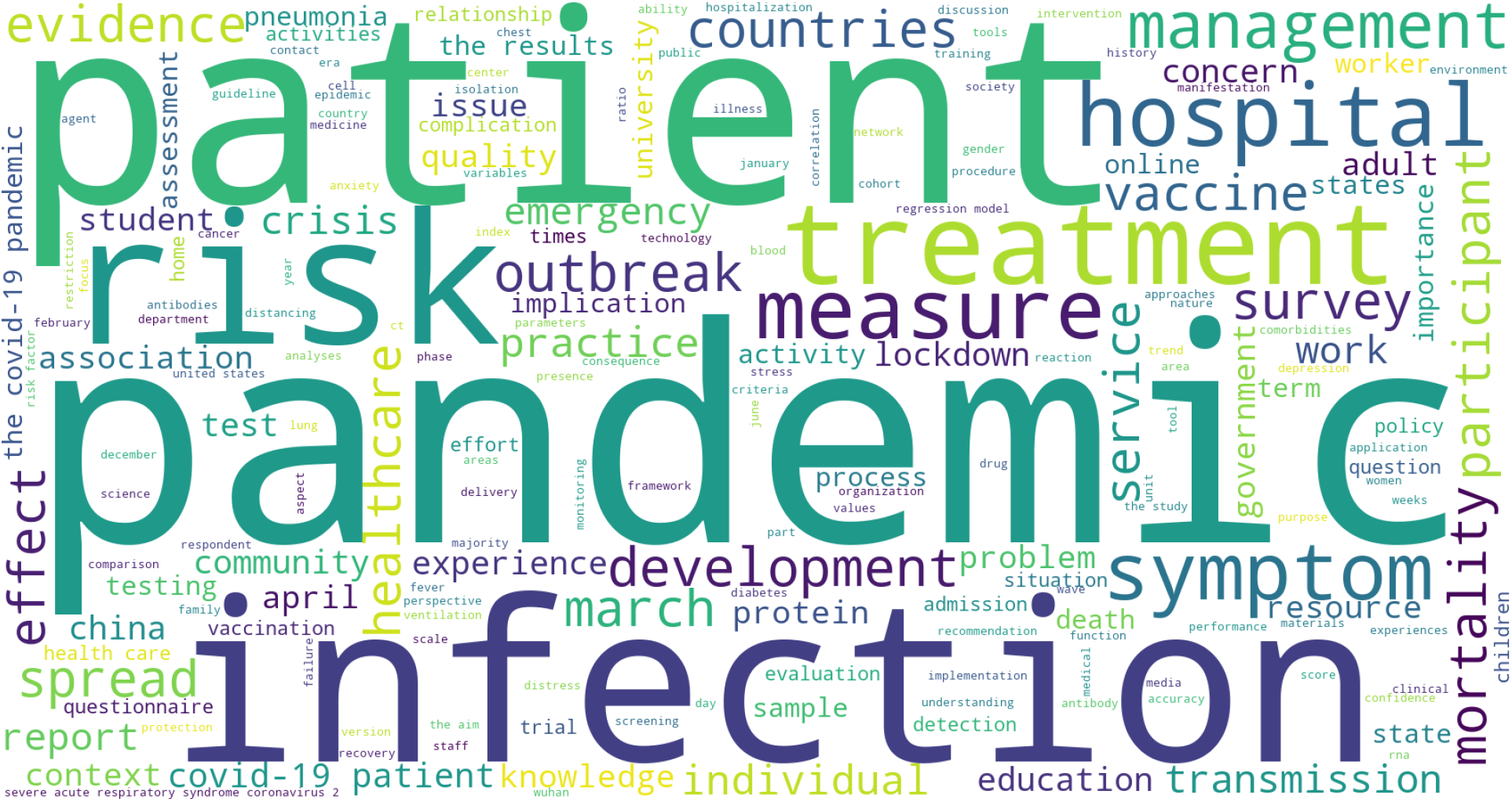
Global top-200 word cloud of all publications

### 1.4 Findings

In particular, this project identifies the following publishing profiles and findings from analyzing the global scientific response to the COVID-19 pandemic:

- Scientists from 189 countries and regions and 175 first-authored countries and regions have contributed to the research on COVID-19, although their research capacity and quality are highly divided.
- 27 subject areas have been involved in studying COVID-19 globally, where medicine, biochemistry/genetics/molecular biology, social science, immunology/microbiology, and environmental science rank the top five subject areas contributing the most publications.
- The US, China, Italy, the UK, and India lead the number of publications, and their cumulative publication composite indices of H5-index, impact factor, CiteScore, SNIP and SJR also rank top-5 globally. However, of the top-20 publishing countries, Netherlands, the UK, France, China and Switzerland lead the worlds in terms of their publication’s mean composite indices.
- The US, China, the UK, Italy and India are the top-5 mostly collaborative countries, where the US appears to be at the center point of global collaboration on COVID-19 research. The US has the highest collaborations with China, the vice versa.
- Of all publications, pandemic, patient, infection, risk, treatment are the top-5 keywords mostly concerned globally.
- Mental health, second wave, lock down, vaccine, and anxiety are the five mostly concerned problems concerned in global research.
- Regression, machine learning and deep learning, simulation, multivariate statistics, artificial intelligence, and statistical model are the mostly applied techniques in quantifying and modeling COVID-19.
- In terms of quantifying COVID-19, in medical science, regression, multivariate statistics, simulation, statistical modeling, and machine learning are mostly applied. In computer science, machine learning, simulation, deep learning, artificial intelligence, and regression are mostly applied. In contrast, social science favors regression, structural equation models, machine learning, simulation, and artificial intelligence.
- In G20 countries and regions, the US, EU, China, the UK, and India rank top-5 in terms of publication number in computer science. In social science, the US, EU, the UK, China and Australia take the lead. In contrast, the US, EU, China, the UK, and India take the lead in medical science.
- The top-5 countries with the highest coefficient between the total publication number and GDP per capita and between the collective composite indicator and GDP per capita are Germany, France, Canada, Spain, and the UK. In contrast, India and China rank the lowest of the top-10 countries, indicating most productive in the COVID-19 research in the context of their GDP per capita.
- In G20 countries and regions, Argentina, South Korea, Japan, Australia and Germany rank the top-5 in terms of their correlation between publication number and GDP per capita; while Argentina, Russia, Japan, South Korea and Australia rank the top-5 in terms of their publication number and collective composite indicator. In contrast, India, China and the US mark the lowest three, showing their higher productivity in terms of their GDP per capita.
- In OECD countries and regions, Iceland, Luxembourg, Estonia rank top 3 in terms of their publication number and GDP per capita; in contrast, US, Turkey and Italy rank have the lowest correlation between their publication number and GDP per capita, i.e., showing relatively higher productivity. The same trends hold for the correlation between publication number and collective composite indicator.
- In the top-20 countries, the top-5 countries with the highest log-normalized correlations between their publication number and new infection case number are Netherlands, Poland, Brazil, France, and Switzerland.
- With regard to the log-normalized correlation between publication number and death number, the top-5 countries are regions are Brazil, Poland, Iran, India and France.

There are significant imbalances in the global COVID-19 research. Mostly developed countries and regions may not necessarily contribute to the most impactful research outcomes, instead countries with low economic status may be more productive. Many underdeveloped countries and regions, including in the African region, suffer from less concentration and productivity in studying COVID-19. In addition, the subject area imbalance and topical imbalance appear across countries and disciplines. With regard to modeling COVID-19, classic methods still play a dominant role in particular in medical and social science. More advanced AI and data science have not been widely applied across disciplines and countries.

## 2 Publication collection

Here, we briefly introduce the collection of COVID-19 research publications, their supplementary information, and their publication impact metrics.

### 2.1 COVID-19 open research data acquisition

The COVID-19 Open Research Dataset (CORD-19) ([**Wang’20**]) was downloaded from SemanticScholar ^12^. The latest CORD-19 data was collected up to 9th March 2022. CORD-19 is a growing resource of scientific publications on COVID-19 and its related historical research on coronavirus^13^.

### 2.2 Supplemental information collection

As the author’s country, affiliation, and the publication date of each publication in SemanticScholar are missing in the CORD-19 dataset, we crawled this additional information from the websites below, cross-checked and refined them for each publication:

- Clarivate’s Web of Science^14^
- ResearchGate^15^
- The US National Library of Medicine ^16^, called PubMed or PMC source below.
- The World Health Organization’s COVID-19 information source^17^
- Crossref^18^
- medRxiv^19^
- arXiv^20^

The Python source codes for crawling the above data are in the folders crawler/WOS_crawler.py, WHO_crawler.py, RG_crawler.py, PMD_keyword_crawler.py, medrxiv_crawler.py, and cross-ref_crawler_by_id.py, respectively.

### 2.3 Collection of publication impact metrics

For each publication, we collected the research impact metrics of their publication venue (journal or conference), including H5-index, Impact Factor, CiteScore, SNIP, and SJR for those available from the following repositories:

- H5-index: each journal or conference publication’s H5-index was crawled from the Google Scholar website^21^, the crawl date is 8th March 2022.
- Impact Factor: was obtained from the Web of Science’s Journal Citation Reports^22^, the data was lastly updated on 1st July 2021 (i.e., the 2020 journal impact factor).
- CiteScore, SNIP and SJR: were obtained from the Scopus website^23^, updated to October 2021.

The collected publication impact data of each publication in this COVID-19 global scientist response dataset can be found in Kaggle file at https://www.kaggle.com/datasets/datascienceslab/covid19-global-scientists-response.

## 3 Publication processing

Here, we briefly introduce the processing of the collected publications. The processing consists of preprocessing the collected publications, extracting author’s country information, extracting publication date, extracting COVID-19 problem keywords, extracting COVID-19 modeling technique keywords, categorizing the disciplines of each publication, and extracting those publications focusing on modeling COVID-19.

The keywords describing each publication were extracted from the title and abstract of each publication. We categorize keywords into two groups: one on COVID-19 problems or topics of interest from a domain perspective (e.g., epidemiological, medical and social perspective, called *problem keywords* in this report), and the other on the techniques for modeling COVID-19 problems (we call them *modeling keywords* in this report). The problem keywords and modeling keywords were extracted by different approaches, explained in Sections 3.4 and 3.5, respectively.

### 3.1 Preprocessing publication data

In the CORD-19 dataset, some publications may be somehow related to COVID-19 but were published before 2020, we thus removed them. In addition, we focus on the publications in English and thus removed those non-English publications collected from the CORD-19 dataset. Finally, as the CORD-19 publications were collected from multiple sources, there are duplicated ones, which were removed as well.

The source code for the data preprocessing of the global publications can be found in data_processing/pre_process.py.

### 3.2 Extracting country information

Here, we collect the author’s country information related to each publication if it is available. The author’s country information of each publication (if available) was extracted from the data crawled from the Web of Science, ResearchGate, the US National Library of Medicine, WHO, Crossref, and medRxiv. However, the country information extracted from these websites is inconsistent and has quality issues. For example, some publications only have affiliations or city information, which have to be transformed to obtain their country information. Hence, an institution-country mapping file is created to map an institution to its country.

The source code for processing author’s country information can be found in the file: *data processing/country_process*.*py*. The extracted country information of each publication in this COVID-19 global scientist response dataset can be found in Kaggle file at https://www.kaggle.com/datasets/datascienceslab/covid19-global-scientists-response.

### 3.3 Extracting publication date

The publishing date of a publication was extracted from the data crawled from the Web of Science, ResearchGate, the US National Library of Medicine, WHO, Crossref, and medRxiv. All dates were transformed to the format of ‘yyyy-mm-dd’.

The source code for extracting and processing publication date can be found in the file: *data processing/publish date process*.*py*. The extracted publication date of each publica-tion in this COVID-19 global scientist response dataset can be found in Kaggle file at https://www.kaggle.com/datasets/datascienceslab/covid19-global-scientists-response.

### 3.4 Extracting problem keywords

Here, we collect the keywords describing the main COVID-19 problems and topics addressed in all publications. We call them COVID-19 “problem keywords.”

We first collected the candidate COVID-19 problems-related keywords from three sources:

- **Keywords provided by publications** In the Web of Science (WOS) and PubMed (PMD) websites, some publications have keywords provided by authors or other tools (e.g., publishers). We crawled these keywords as well as each publication’s supplementary information (including author’s affiliation, country, and publication date). The keywords are regarded as the candidate problem keywords. The related source code for collecting these keywords can be found in crawler/PMD_keyword_crawler.py, WOS_crawler.py.
- **Keywords extracted by PositionRank** COVID-19 problems-related words were further extracted from the titles and abstracts of all publications. We used PositionRank ^24^ ([**Corina’17**]) to conduct the text segmentation of titles and abstracts. The position rank captures both the positions of words in the title and abstract of each publication and the word occurrence frequencies in the title and abstract. Then, the PositionRank is calculated by the position-biased PageRank, which captures the co-occurrence word’s position information. As the topic of a publication usually occurs in the very beginning of its title/abstract, PositionRank is thus suitable to extract problem-related keywords. We selected top-5 keywords as the candidate problem keywords for each publication. The source code can be in /keyword_extraction/keyword_position_rank/main_process.py.
- **Domain-driven keywords** In some publications, no specific keywords were provided, where their synonyms and other expressions were used instead. For example, for a publication with the title “A novel model to predict severe COVID-19 and mortality using an artificial intelligence algorithm,” we know it is about “mortality prediction’ although the phrase “mortality prediction” does not appear in the title. To find out such specific topics of those publications, we used a domain-driven method to work out their relevant topics. First, we created a mapping list of expressions/patterns to a specific topic, for example, two lists for the problem-related keyword “mortality prediction” as follows: *{forecasting, forecast, forecasts, predict, prediction, estimate, estimation, time series, modeling, modelling*, and *model}* and *{death rate, mortality rate, fatality rate, death case, death, fatality*, and *mortality}*, If a publication’s title and abstract contain one of the keywords in both lists, then the phrase “mortality prediction” would be added as the domain-driven problem keyword to this publication. The source code can be found in data_processing/domain_process.py.

After we obtained the keywords generated by the above three approaches, all keywords for each publication were combined to form this publication’s candidate problem keywords.

The file containing the problem keywords of each publication in this COVID-19 global scientist response dataset can be found in Kaggle file at https://www.kaggle.com/datasets/datascienceslab/covid19-global-scientists-response.

### 3.5 Extracting modeling keywords

Here, we explain how the modeling-specific keywords of all publications were generated in this project. First, we extracted the keywords and phrases from the title and abstract of each extracted publication. However, those extracted terms are not all related to modeling. To select modeling-specific terms, we calculate the cosine similarity between each term and the embedding (mean value) of all predefined modeling keywords in Appendix 10.2. We then sorted the similarity scores of all terms in the descending order. We further manually selected those terms with high scores. Below, we introduce them in detail.

- We used Apache OpenNLP to extract keyword and key phrases from the titles and abstracts of all publications. To narrow down the search scope, we only chose those terms with their length no greater than three. Those extracted terms are regarded as the basic keywords of each publication.
- To obtain the embedding of a keyword, we first collected a list of predefined modeling-related keywords per domain knowledge and prior information, such as machine learning, deep learning, statistical model, compartmental model, epidemic modeling, and other modeling techniques and methods. These predefined modeling keywords can be found in Appendix 10.2: List of predefined modeling keywords. Then we applied SciBert ([**Beltagy’19**]) to generate the embeddings of these predefined keywords. SciBert^25^ is a pretrained language model based on BERT, which was trained on a large corpus of scientific publications. We then obtain the embedding of each keyword by SciBert. After that, an average embedding value was calculated upon the embeddings of all keywords, which is called *predefined modeling keyword embedding*.
- For each term extracted from a publication, we further generated its embedding by SciBert, forming the *extracted keyword embedding*.
- Then, for each extracted keyword embedding from its publication, we applied the cosine similarity function to calculate the similarity between this embedding and the predefined modeling keyword embedding. We further sorted the extracted keywords based on this similarity score. Those extracted keywords with high similarity score are chosen as the modeling keywords of each publication.

The source code can be found in “data_processig\keyword_process.py.” The file collecting the modeling technical keywords of each publication in this COVID-19 global scientist response dataset can be found in Kaggle file at https://www.kaggle.com/datasets/datascienceslab/covid19-global-scientists-response.

### 3.6 Extracting disciplinary categorization

Here, in all publications, we extract those publications associated with three major disciplines: medical science, computer science, and social science. Here, *medical science* refers to anything related to medicine, medical science, health science, and epidemiology, etc. *Computer science* groups any computing-related areas, including applied mathematics, IT, computing, and informatics. *Social science* publications were collected from SSRN.

The definition of each discipline is based on the categorization collected from three websites: Web of Science, SJR^26^, and Research.com^27^. This disciplinary categorization can be found in Appendix 10.1: List of disciplinary categorization. In each category, there are a list of journals (and conferences). Based on this categorization, each publication is assigned to its related disciplines. As a journal may involve two disciplines, some publications may thus appear in two disciplines. As the disciplinary sources are from different websites, below, we introduce how we categorized a publication per the above different sources.

- **Web of Science**: we crawled a publication’s disciplinary category from the WOS website, based on Appendix 10.1: List of disciplinary categorization. We added the publication to its related disciplinary categories. For example, if a publication’s category falls in Cell Biology based on the categorization, we then add this publication to the discipline of medical science.
- **SJR**: In the SJR website, a journal (or conference) is labeled by one to multiple categories. Each category has a collection of journals and conferences. We crawled the sources (i.e., the names of journals and conferences) of publications from ResearchGate, the US National Library of Medicine, WHO, Crossref, and medRxiv. According to these sources of each publication, we added the publication to its categorized disciplines.
- **Research.com**: previously also called Guide2Research, was a research portal initially for computer science and is now for broad disciplines. It lists a large number of computer science sources (journals and conferences). Based on a publication’s sources obtained from ResearchGate, the US National Library of Medicine, WHO, Crossref and medRxiv, we added the publication to its mostly relevant disciplines.

Based on the disciplinary files generated per the above three sources, we combined the data from these three sources to generate the unified disciplinary files of publications.

The source code can be found in “data_processing\domain_process.py.” The generated disciplinary information of each publication in this COVID-19 global scientist response dataset can be found in Kaggle file at https://www.kaggle.com/datasets/datascienceslab/covid19-global-scientists-response.

### 3.7 Extracting modeling publications

Here, we extracted those modeling-specific publications from the entire publication set. The modeling publications are those containing at least one of the modeling keywords and expressions.

The file “Modeling related words and expressions.txt” collected the modeling words and expressions.

### 3.8 Extracting global GDP and population data

Here, we collected the Gross Domestic Product (GDP) and population data of each country globally from the World Population Review^28^.

Accordingly, we calculate the GDP per Capita for each country, as shown in Section 4.3.

### 3.9 Extracting COVID-19 infection cases

Here, we extract the number of infected cases and the number of deaths in each country. The case numbers of all countries were collected from Our World in Data^29^.

This case data will be used to analyze the correlation between COVID-19 publications and the case numbers for each country or region.

## 4 Measurement

Here, we introduce the measures and metrics applied for analyzing or evaluating the COVID-19 publications. They include publication impact metrics, composite indicator, and correlation coefficient.

### 4.1 Publication impact metrics

To quantify the quality and impact of each publication and the publications associated with a target variable, e.g., a country, we consider the following publication impact metrics: H5-index, Impact Factor, CiteScore, SNIP, and SJR. We consider their original values, mean values, and normalized values, respectively.

#### Original impact metrics

Here, we introduce the five original impact metrics.

- H5-index: created by Google, “is the largest H-index for articles published in a publication journal or conference in the last 5 complete years, which had at least h citations for each article”, available at Google Scholar^30^.
- Impact Factor: i.e., journal impact factor (JIF or IF), created by the Web of Science announced in its annual Journal Citation Reports (JCR), “is calculated by dividing the number of current year citations to the source items published in that journal during the previous two years”^31^, available at annual JCR reports^32^.
- CiteScore: created by Elsevier, “calculates the citations from all documents in year one to all documents indexed on Scopus published in the prior three years for a publication”^33^, available from the Scopus Journal Metrics^34^.
- Source Normalized Impact per Paper: i.e, SNIP, “measures the contextual citation impact by weighting citations based on the total number of citations in a subject field”^35^, available from the Scopus Journal Metrics^36^.
- SCImago Journal Rank: i.e., SJR, “is based on the concept of a transfer of prestige between journals via their citation links. Drawing on a similar approach to the Google PageRank algorithm - which assumes that important websites are linked to from other important websites - SJR weights each incoming citation to a journal by the SJR of the citing journal, with a citation from a high-SJR source counting for more than a citation from a low-SJR source”^37^, available from the Scopus Journal Metrics^38^.

#### Mean publication impact metrics

We further calculate the *mean publication impact metrics* of each publication in terms of the five original impact metrics: H5-index, Impact Factor, CiteScore, SNIP, and SJR, respectively. The *mean publication impact metrics* measures the publication-averaged impact metric of all publications in terms of a target variable, e.g., a discipline, or a country. For example, Section 5.1.5 reports the mean publication impact in three major disciplines, respectively. Sections 6.5 and 6.6 report the mean publication impact of G20 and OECD countries and regions, respectively.

#### Normalized publication impact metrics

To better compare the relative publication impact of all publications in terms of an impact metric, we also generate its relative publication impact metric by normalizing the values of publications. As the impact values from different countries are usually nonlinear and highly skewed, we take the following log normalization function scaled to the maximum log normalization for better comparison and visualization effects, which generates *log-normalized impact metrics*.

For example, to log-scale H5-index (H5) to obtain its log-normalized relative H5-index 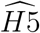 of each publication *i*,

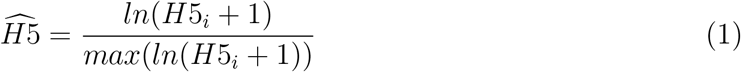

where *max* is to extract the maximum log value of H5-index over all publications. Further, to obtain the log-scaled H5-index for a country, we sum its values of all publications from that country to obtain its cumulative log-scaled H5-index. This result is then applied to compare the publication impact between countries, as shown in Section 5.10.

Similarly, we log normalize the Impact Factor, CiteScore, SNIP, SJR, and CI (to-be-introduced in Section 4.2) values of all publications to obtain their relative log-normalized impact metrics.

In addition, to analyze the overall impact of all publications (e.g., of a country or discipline), we further introduce the *log-normalized cumulative impact metrics*. For example, to calculate the log-normalized cumulative H5-index (*sum*(*H*5_*i*_)) of a country *i* as follows:

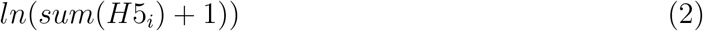

These log-normalized cumulative impact metrics are applied to analyze the publication impact in Sections 6.1, 6.2, 6.3, and 6.4.

We also introduce the *log-normalized publication number* as follows for a target variable, e.g., a country or discipline.

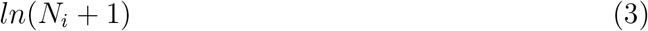

Here, *N*_*i*_ refers to the number of publications of the target variable, e.g., country *i*.

This log-normalized cumulative publication number is applied in Sections 6.1, 6.2, 6.3, and 6.4 to analyze the relative impact of G20 and OECD countries on the COVID-19 research.

### 4.2 Composite indicator (CI)

Here, we propose three collective impact metrics to measure the overall impact of each publication, the mean collective impact of a publication, and the cumulative impact of all publications selected per a criterion (e.g., a country or a discipline), respectively. They are called *publication composite indicator, cumulative composite indicator*, and *mean composite indicator*.

#### Publication composite indicator

In our work, we create the *Publication Composite Indicator* (CI, *C*_*i*_, which is also generally called composite indicator for simplicity), which is defined below, as one of the research impact metrics to measure the collective impact of a publication in terms of five vendor-created individual major impact metrics: H5-index (H5), Impact Factor (IF), CiteScore (CS), SNIP (SN), and SJR (SJR).

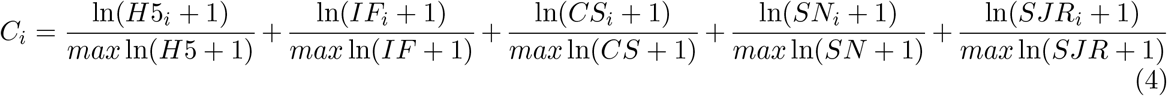

where *i* refers to each publication, *max* is the operation of extracting the maximum value. The H5-index data was obtained from Google Scholar, impact factor values were from the Web of Science, the CiteScore, SNIP and SJR were obtained from the Scopus sources.

#### Cumulative composite indicator

Given a grouping criterion such as a country or a discipline, all of their publications can be selected. After calculating the composite indicator of each of their publications, we can then sum the publication composite indicator to obtain the cumulative composite indicator *𝒞*_*i*_.

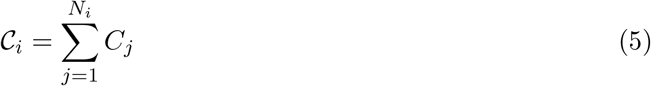

Here, *i* refers to a given target, e.g., a country *i, N*_*i*_ refers to the number of total publications with the information about composite indicator available from the target variable, *C*_*j*_ is the composite indicator of each publication *j* of *N*_*i*_.

The cumulative composite indicator is applied in analyzing the global publication impact such as in Section 5.4 (see Figure 4), and analyzing the correlation between publication composite impact and GDP for a country as in Section 8.1 (see Figures 32 and 33).

**Figure 4:**
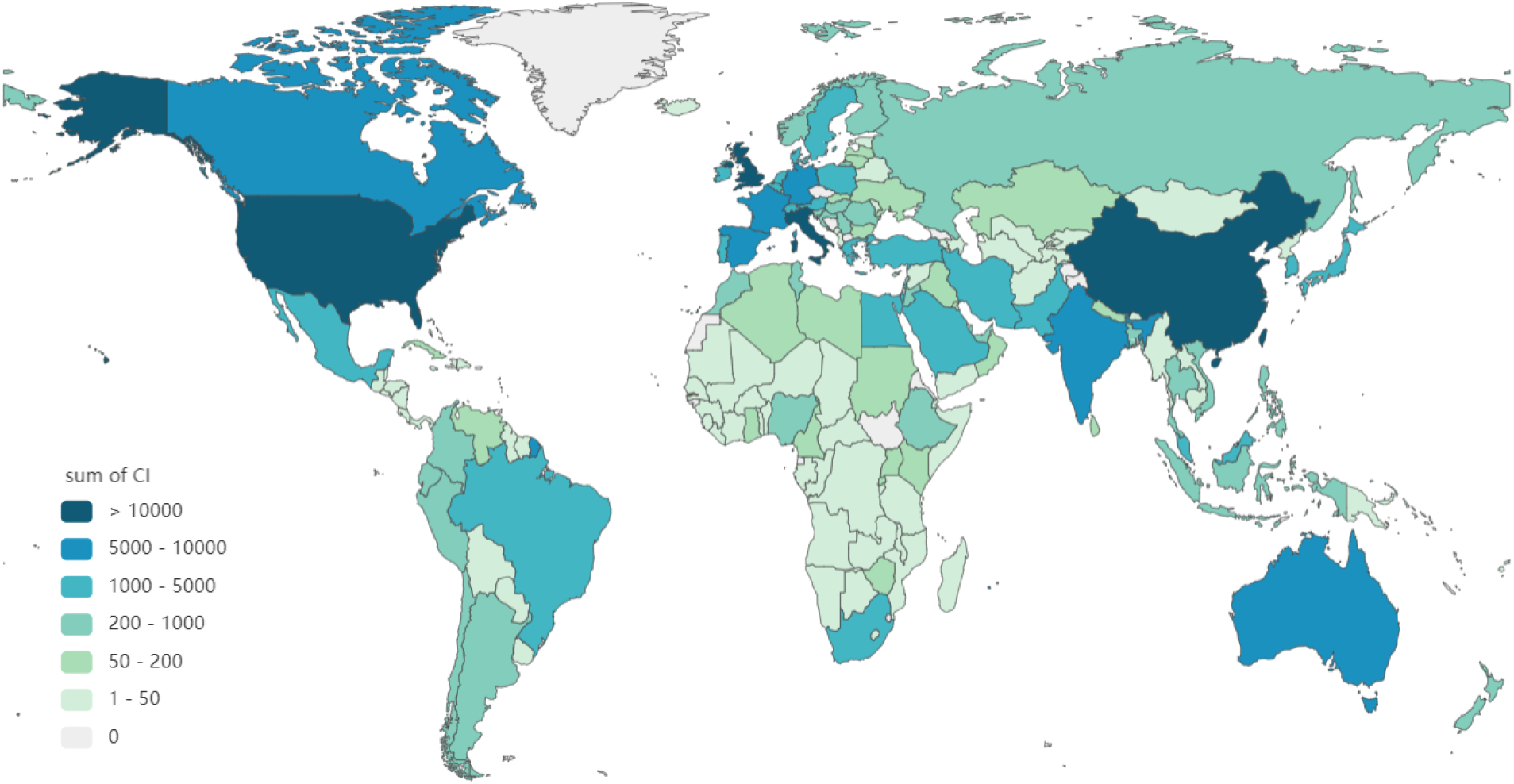
Global publication impact distribution per cumulative Composite Indicator (CI)

#### Mean composite indicator

With the cumulative composite indicator *𝒞*_*i*_ and the number of total publications *N*_*i*_ from a target variable e.g., a country *i* with the information for calculating the composite indicator, we can then calculate the *mean composite indicator* 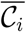 as follows:

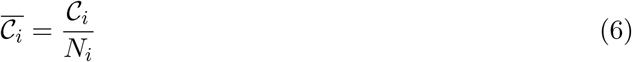

Examples of using the mean composite indicator are the analysis of the top-10 published country’s research impact in Section 5.10 (as in Figure 10) and the top-10 published country’s disciplinary research impact in Section 5.11 (as in Figure 11), and the analysis of the G20 country’s mean publication impact in Section 6.5 (see Figure 19).

### 4.3 GDP per capita

Here, we calculate the Gross Domestic Product (GDP) per Capita (GDPPC, 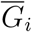) as follows, which measures a country’s economic output per person.

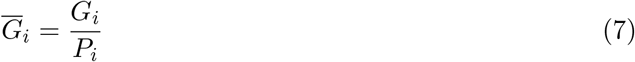

Here, *G*_*i*_ is the gross domestic product of country or region *i*, and *P*_*i*_ is the number of total population of that country or region. *G*_*i*_ and *P*_*i*_ are collected per the tool introduced in Section 3.8.

Examples of using GDPPC are the analysis of the correlation between COVID-19 publications and the GDP per capita of of all countries in Section 8.1 (see Figures 32 and 33) and the correlation between publications and GDP per capita in G20 countries and regions in Section 8.2 (see Table 28).

### 4.4 Publication-GDP correlation coefficient

We further calculate the correlation coefficient between a country’s publication metrics (e.g., publication number, and CI) and their economic metrics (e.g., GDP per capita) or COVID-19 infection metrics (e.g., infected case number, death number) of the country. Specifically, the *publication correlation coefficient* is defined for a first-authored country in terms of their total publication number and the cumulative composite indicator of the publications with another variable such as GDP per capita in that country.

Further, three publication-GDP coefficients are calculated to measure the correlations between the publication number (N) and the GDP per capita (G) and between the composite indicator (CI) and the GDP per capita (G) for each country, respectively.

First, the *absolute similarity* between a publication metric and the GDP per capita of a country is measured by *absolute publication-GDP coefficient* (*Ab_corr*) *ρ*. Accordingly, we define the *absolute publication number-GDP coefficient ρ*_*n*_ below:

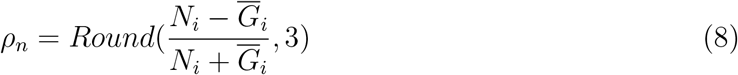

where *N*_*i*_ refers to the publication number of country *i*, and 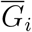 refers to the GDP per capita of the country.

Similarly, we define the *absolute publication composite impact-GDP coefficient* (*Ab_corr*) *ρ*_*c*_ as follows:

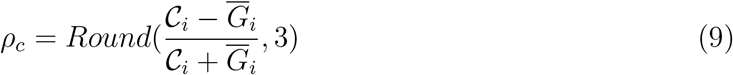

where *𝒞*_*i*_ refers to the cumulative composite indicator value of all publications from country *i*, and 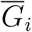 refers to the GDP per capita of the country.

Both *ρ*_*n*_ and *ρ*_*c*_ fall in the value range [*−*1, 1] symmetric about 0 with no correlation. The magnitude of values reflect more positive or negative correlation between the comparison objects, i.e., more strongly co-move in the same or different direction.

Second, the maximum-normalized coefficient between the publication metrics and GDP per capita is measured by the *relative publication-GDP coefficient* (*Re_corr*) 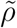, which normalizes the publications and GDP per their respective maximum values. Accordingly, we define the *relative publication number-GDP coefficient* 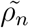 below:

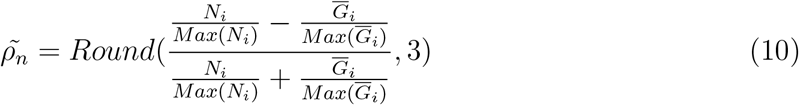

where *N*_*i*_ refers to the publication number of country *i*, 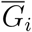 refers to the GDP per capital of the country, *Max*(*N*_*i*_) refers to the maximum value of publication number over all countries, *Max* 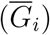 refers to the maximum value of GDP per capita over all countries.

Similarly, we define the *relative publication composite impact-GDP coefficient* (*Re_corr*) 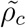 as follows:

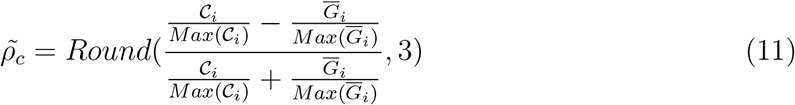

where *𝒞*_*i*_ refers to the cumulative composite indicator of country *i*, 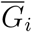 refers to the GDP per capita of the country, *Max*(*𝒞*_*i*_) refers to the maximum value of composite indicator over all countries, *Max* 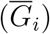 refers to the maximum value of GDP per capita over all countries.

Both 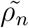 and 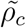 fall in the value range [*−*1, 1] symmetric about 0 with no correlation. The magnitude of values reflect more positive or negative correlation between the comparison objects, i.e., more strongly co-move in the same or different direction.

Third, we further measure the *dispersion-deviation coefficient De_corr* between two comparison objects in terms of their dispersion/deviation from the mean of the cohort. Accordingly, we define the *publication-GDP dispersion coefficient* 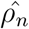 between publication number *N*_*i*_ and GDP per capita *G*_*i*_ as follows:

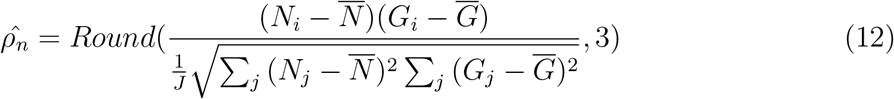

where 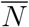 refers to the mean of publication number of all *J* countries and regions; 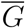 refers to the mean of GDP per capita of all countries and regions; and *N*_*j*_ and *G*_*j*_ refer to the publication number and GDP per capita of country/region *j*, respectively.

Similarly, we define the *dispersion coefficient between publication composite impact and GDP* 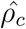 as follows:

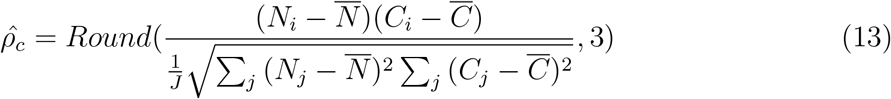

where 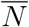 refers to the mean of publication number of all *J* countries and regions; 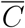 refers to the mean of composite indicator of all countries and regions; and *N*_*j*_ and *C*_*j*_ refer to the publication number and composite indicator of country/region *j*, respectively.

Both 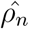 and 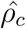 fall in the value range [*−∞*, +*∞*] symmetric about 0 with no correlation. The magnitude of values reflect more positive or negative correlation between the comparison objects, i.e., more strongly co-move in the same or different direction.

### 4.5 Publication-COVID-19 infection correlation coefficient

Here, we introduce measurement to quantify the correlation between the COVID-19 publications and the COVID-19 infections. Since the infections and deaths are very high for many countries, the value ranges of publications and infections and deaths are very divided, and their values across countries and regions are also highly divided and incomparable, it would be difficult to directly analyze their correlation on the original values. It is thus unsuitable to follow the same approach as in Section 4.4 (i.e., Equations (8) to (11)) to measure the publication-infection correlations.

Instead, we natural-log-normalize the number of publications and the number of infections and then apply them to Equation (8) to measure the correlation between COVID-19 publications and infections and deaths respectively. We thus obtain the *log-normalized publication-infection coefficient ρ*_*i*_:

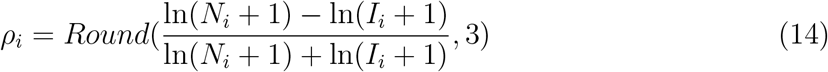

where ln *N*_*i*_ is the log-normalized value of publications of a target variable *i* (e.g., for a country, a month, or a discipline), ln *I*_*i*_ refers to the log-normalized value of the infections of a target variable *i*.

Accordingly, we also obtain the *log-normalized publication-death coefficient ρ*_*d*_:

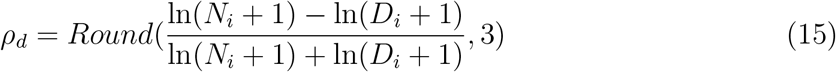

where ln *N*_*i*_ is the log-normalized value of publications of a target variable *i* (e.g., for a country, a month, or a discipline), ln *D*_*i*_ refers to the log-normalized value of the deaths of a target variable *i*.

Lastly, we further measure the *dispersion-deviation coefficient De_corr* between publications and infections/deaths for each country/region in terms of their dispersion/deviation from the mean of the cohort. Accordingly, we define the *log-normalized publication-infection dispersion coefficient* 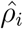 between publication number *N*_*i*_ and log-normalized infection number ln *N*_*i*_ + 1) as follows:

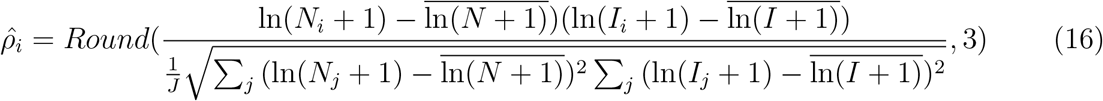

where 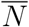 refers to the mean of publication number of all *J* countries and regions; 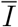 refers to the mean of infections of all countries and regions; and *N*_*j*_ and *I*_*j*_ refer to the publication number and infections of country/region *j*, respectively.

Similarly, the *log-normalized publication-death dispersion coefficient* 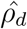 between publication number *N*_*i*_ and log-normalized infection number ln(*D*_*i*_ + 1) as follows:

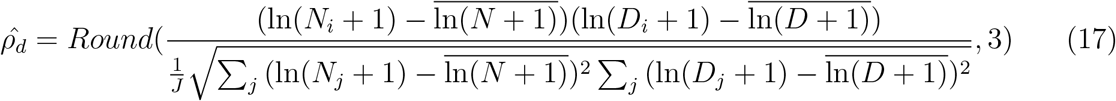

where 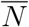 refers to the mean of publication number of all *J* countries and regions; 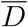 refers to the mean of deaths of all countries and regions; and *N*_*j*_ and *D*_*j*_ refer to the publication number and deaths of country/region *j*, respectively.

Both 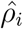 and 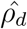 fall in the value range [*−∞*, +*∞*] symmetric about 0 with no correlation. The magnitude of values reflect more positive or negative correlation between the comparison objects, i.e., more strongly co-move in the same or different direction.

## 5 Global publication profile

Following the review objectives and questions and the first review objective of generating an overview of the global COVID-19 research profile, as discussed in Section 1.1, here we present the results of identified global COVID-19 publication profile.

### 5.1 Global publication overview

Here, we present an overview of the global publications on COVID-19, produced by all first-authored countries, including the results of total publications, disciplinary publications, publication statistics with publication dates and author countries, the statistics of publication impact, and the mean publication impact of major disciplines.

#### 5.1.1 Total publications

Here, we summarize the count of all publications on COVID-19 from all disciplines and by all published countries. The collected publications comprise official publications and preprints.

We further extracted those on COVID-19 modeling, called *modeling publications* in this report. Modeling publications are those with modeling keywords, as shown in Appendix 10.1, appearing in their titles and abstracts.

Table 1 shows the overall statistics of COVID-19 publications. In total, up to 9 March 2022, there have been 319,847 published references and 26,420 preprints available online, amounting to 346,267 references collected on COVID-19. These publications involve authors from 189 countries.

**Table 1:**
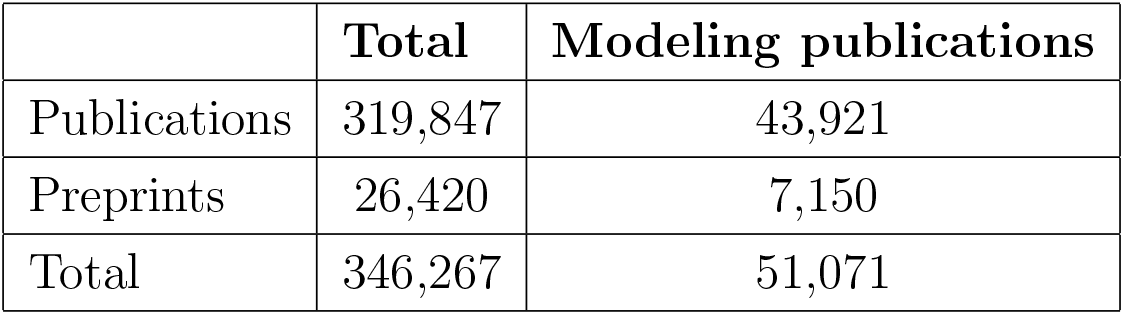
Overall statistics of publications

#### 5.1.2 Disciplinary publications

Here, we report the statistics of the disciplinary publications. All publications are segmented into disciplinary areas (subjects) by using the subject categorization listed in the Scimago Journal Country Rank^39^. There are 233,387 publications in 27 subject areas contributed to the COVID-19 research, which include ‘Agricultural and Biological Sciences’, ‘Arts and Humanities’, ‘Biochemistry, Genetics and Molecular Biology’, ‘Business, Management and Accounting’, ‘Chemical Engineering’, ‘Chemistry’, ‘Computer Science’, ‘Decision Sciences’, ‘Dentistry’, ‘Earth and Planetary Sciences’, ‘Economics, Econometrics and Finance’, ‘Energy’, ‘Engineering’, ‘Environmental Science’, ‘Health Professions’, ‘Immunology and Microbiology’, ‘Materials Science’, ‘Mathematics’, ‘Medicine’, ‘Multidisciplinary’, ‘Neuroscience’, ‘Nursing’, ‘Pharmacology, Toxicology and Pharmaceutics’, ‘Physics and Astronomy’, ‘Psychology’, ‘Social Sciences’ and ‘Veterinary.’ There are 86,460 publications without clear alignment with their subject areas. Figure 1 shows the subject distribution of the COVID-19 publications. According to the pie chart, ‘Medicine’ occupies the largest percentage in all the publications, followed by subjects ‘Biochemistry, Genetics and Molecular Biology’, ‘Social Sciences’ and ‘Immunology and Microbiology.’ Table 2 shows the detailed data of the subject distribution of COVID-19 publications.

**Table 2:** Disciplinary distribution of COVID-19 publications

| **Subject areas** | **Publication number** | **Pub percentage** |
| --- | --- | --- |
| Medicine | 164,082 | 5.13 |
| Biochemistry, Genetics and Molecular Biology | 26,689 | 0.834 |
| Social Sciences | 25,231 | 0.789 |
| Immunology and Microbiology | 19,042 | 0.595 |
| Environmental Science | 12,837 | 0.401 |
| Pharmacology, Toxicology and Pharmaceutics | 10,736 | 0.336 |
| Nursing | 9,653 | 0.302 |
| Psychology | 9,242 | 0.289 |
| Computer Science | 8,605 | 0.269 |
| Engineering | 8,215 | 0.257 |
| Multidisciplinary | 8,125 | 0.254 |
| Business, Management and Accounting | 6,545 | 0.205 |
| Neuroscience | 6,358 | 0.199 |
| Health Professions | 5,794 | 0.181 |
| Chemistry | 5,223 | 0.163 |
| Economics, Econometrics and Finance | 5,124 | 0.16 |
| Agricultural and Biological Sciences | 4,993 | 0.156 |
| Arts and Humanities | 4,761 | 0.149 |
| Physics and Astronomy | 4,019 | 0.126 |
| Mathematics | 3,629 | 0.113 |
| Chemical Engineering | 3,476 | 0.109 |
| Energy | 2,936 | 0.092 |
| Materials Science | 2,929 | 0.092 |
| Earth and Planetary Sciences | 2,169 | 0.068 |
| Dentistry | 1,723 | 0.054 |
| Decision Sciences | 1,533 | 0.048 |
| Veterinary | 1,484 | 0.046 |

Further, we categorize those publications in terms of their relevance to the three major disciplines: computer science, medical science, and social sciences.

Table 3 shows the results of all collected publications labelled to these three major disciplines: computer science, medical science, and social sciences. Of all 346,267 publications from all disciplines, there are 13,959 associated with computer science, 186,650 associated with medical science, and 32,128 associated with social science.

**Table 3:** Distribution of COVID-19 publications in major disciplines

|  | **Overall** | **Modeling publications** |
| --- | --- | --- |
| Medical science | 186,650 | 21,785 |
| Computer science | 13,959 | 6,142 |
| Social science | 32,128 | 3,890 |

#### 5.1.3 Publications with publication date and first-authored country

Here, we report the publications with publication dates and first-authored country information, which were collected per the methods introduced in Sections 3.3 and 3.2.

Of 189 countries and regions associated with the tot publications, there are 175 first-authored countries appearing in the publications, which spread all five continents. Of 319,847 published references, there are 272,060 with publication dates.

Table 4 shows the number of publications with publication dates and first-author’s country information in those published literature (i.e., excluding those preprints), respectively.

**Table 4:** Publications with publication date and first-authored country

|  | **Overall** |
| --- | --- |
| with publication date | 272,060 out of 319,847 published references |
| with first-authored country | 211,178 out of 319,847 published references |

#### 5.1.4 Statistics of publication impact

Here, we present the statistics of the publication impact in terms of major publication impact metrics: H5-index, Impact Factor, CiteScore, SNIP, and SJR. The values of these impact metrics for the collected publications were collected per the introduction in Section 2.3.

Over all 346,267 published and preprint publications, 243,597 are with H5-index, 199,284 are with Impact Factor, 250,998 are with CiteScore, 246,640 are with SNIP, and 247,403 are with SJR. There are 50,792 references without any of the impact metrics H5-index, Impact Factor, CiteScore, SNIP, and SJR.

Table 5 shows the statistics of these impact metrics of all collected publications with the impact metrics issued on their publication venues. Num_pub refer to the number of publications, max, min, mean and median refer to the statistics of original impact metrics values associated with each publication.

**Table 5:** Statistics of publication impact metrics of all publications

|  | Valid Num_pub | Max score | Min score | Mean | Median |
| --- | --- | --- | --- | --- | --- |
| H5-index | 243,597 | 414 | 1 | 53.8 | 34 |
| Impact Factor | 199,284 | 508.7 | 0.018 | 5.55 | 2.49 |
| CiteScore | 250,998 | 463.2 | 0.1 | 8 | 3.4 |
| SNIP | 246,640 | 143.645 | 0.001 | 1.83 | 1.34 |
| SJR | 247,403 | 62.937 | 0.1 | 1.66 | 0.97 |

#### 5.1.5 Mean publication impact in major disciplines

Here, we present the mean publication impact of publications from the three main disciplines: computer science, medical science, and social science. The disciplinary publications were collected in Section 5.1.2, their impact metrics were collected by the method introduced in Section 2.3. Here, we only count those references with at least one of the five impact metrics available on their publication venues.

Table 6 shows the paper-averaged score of the five publication impact metrics: H5-index, Impact Factor, CiteScore, SNIP, and SJR, in three disciplines: CS - computer science, MS - medical science, and SS - social science. We call the paper-averaged score *mean publication impact*. In the table, *Num_pub* refer to the number of publications, *Mean_H*5 refers to the mean H5-index of each publication, *Mean_IF* refers to the mean Impact Factor of each publication, *Mean_CS* refers to the mean CiteScore of each publication, *Mean_SN* refers to the mean SNIP of each publication, and *Mean_SJR* refers to the mean SJR of each publication.

**Table 6:** Mean publication impact score in three disciplines

|  | H5-index |  | Impact Factor |  | CiteScore |  | SNIP |  | SJR |  |
| --- | --- | --- | --- | --- | --- | --- | --- | --- | --- | --- |
|  | Num<br>_pub | Mean<br>_H5 | Num<br>_pub | Mean<br>_IF | Num<br>_pub | Mean<br>_CS | Num<br>_pub | Mean<br>_SN | Num<br>_pub | Mean<br>_SJR |
| MS | 171,019 | 61.86 | 151,290 | 7 | 174,086 | 6.97 | 172,052 | 1.74 | 173,493 | 1.72 |
| CS | 11,741 | 60.65 | 9,081 | 4.47 | 12,669 | 5.18 | 12,542 | 1.56 | 12,557 | 0.91 |
| SS | 27,414 | 41.65 | 19,650 | 3.53 | 29,219 | 3.45 | 28,919 | 1.37 | 29,062 | 0.85 |

### 5.2 Global publication distribution

Here, we report the distribution of global publications on COVID-19. The *global publication distribution* refers to the publication number distribution of first-authored countries of all publications with country information. The first-authored countries were crawled from the Web of Science, ResearchGate, WHO, Crossref, and the US National Library of Medicine, as explained in Section 3.2.

Figure 2 shows the publication distribution of 175 first-authored countries, where the publication of each first-authored country is plotted to the World Map. We further extracted the top-20 first-authored countries in terms of their publication number in Table 7. The top-20 countries are the US, China, Italy, the UK, India, Spain, Canada, Germany, France, Brazil, Iran, Australia, Turkey, Japan, Saudi Arabia, South Korea, Netherlands, Poland, Switzerland, and Pakistan.

**Table 7:** Global publication distribution

| Country/Region | Publication number | Percentage of publications |
| --- | --- | --- |
| EU | 48,140 | 15.05% |
| G20 | 182,097 | 56.93% |
| OECD | 139,588 | 43.64% |
| US | 49,203 | 15.38% |
| China | 21,693 | 6.78% |
| Italy | 15,029 | 4.70% |
| UK | 14,475 | 4.53% |
| India | 13,044 | 4.08% |
| Spain | 6,372 | 1.99% |
| Canada | 6,093 | 1.90% |
| Germany | 5,857 | 1.83% |
| France | 5,196 | 1.62% |
| Brazil | 5,024 | 1.57% |
| Iran | 4,914 | 1.54% |
| Australia | 4,719 | 1.48% |
| Turkey | 4,600 | 1.44% |
| Japan | 3,678 | 1.15% |
| Saudi Arabia | 2,792 | 0.87% |
| South Korea | 2,696 | 0.84% |
| Netherlands | 2,123 | 0.66% |
| Poland | 2,012 | 0.63% |
| Switzerland | 1,880 | 0.59% |
| Pakistan | 1,797 | 0.56% |

The data of the global publication distribution can be found in the file named “Global publication distribution.xlsx.”

The US alone contributed to 49,203 references, amounting to 15.38% of the total published COVID-19 scientific references. China made the second largest contribution, amounting to 21,693 i.e., 6.78%, while EU together contributed to 48,140 publications, amounting to 15.05%. G20 countries and regions contributed to 182,097 publications, amounting to 56.93%. OECD countries and regions contributed to 139,588 publications, amounting to 43.64%.

### 5.3 Word cloud of global publications

Here, we present the most frequent keywords extracted from the NLP analysis of all global publications and present them in word cloud^40^.

Figure 3 shows the word cloud of the top-200 words appeared in all publications. The source code for generating the word cloud can be found in the data processing/word cloud.py. The full results can be found in “Word cloud of global publications.xlsx.”

The top-20 keywords appearing in the global publications are: ‘pandemic’, ‘patient’, ‘infection’, ‘risk’, ‘treatment’, ‘hospital’, ‘measure’, ‘symptom’, ‘management’, ‘development’, ‘outbreak’, ‘mortality’, ‘countries’, ‘survey’, ‘march’, ‘spread’, ‘participant’, ‘effect’, ‘evidence’ and ‘healthcare.’

### 5.4 Global publication impact distribution per Composite Indicator

Here, we present the global publication impact distribution by world map in terms of the cumulative composite indicator value of each first-authored country. The definition of composite indicator is given in Section 4.2. Only those publications with at least one metric value available from the five metrics: H5-index, Impact Factor, CiteScore, SNIP and SJR are counted here. Accordingly, for each country, at least one of their first-authored publications must have the CI value in order to be included in the world map visualization.

Figure 4 shows the sum of each publication’s composite indicator (CI) value of each first-authored country for all countries with first-authored publications. 174 countries are with CI values, while one country (Vanuatu) missing the CI value as there is no CI with some of their publications. The definition of Composite Indicator can be found in Section 4.2. The full results of all countries can be found in “Global research impact distribution per Composite Indicator (CI).xlsx.”

Further, the top 20 countries in terms of the sum of their publication’s CI values are listed in Table 8, where the US, China, the UK, Italy and India mark the top 5.

**Table 8:**
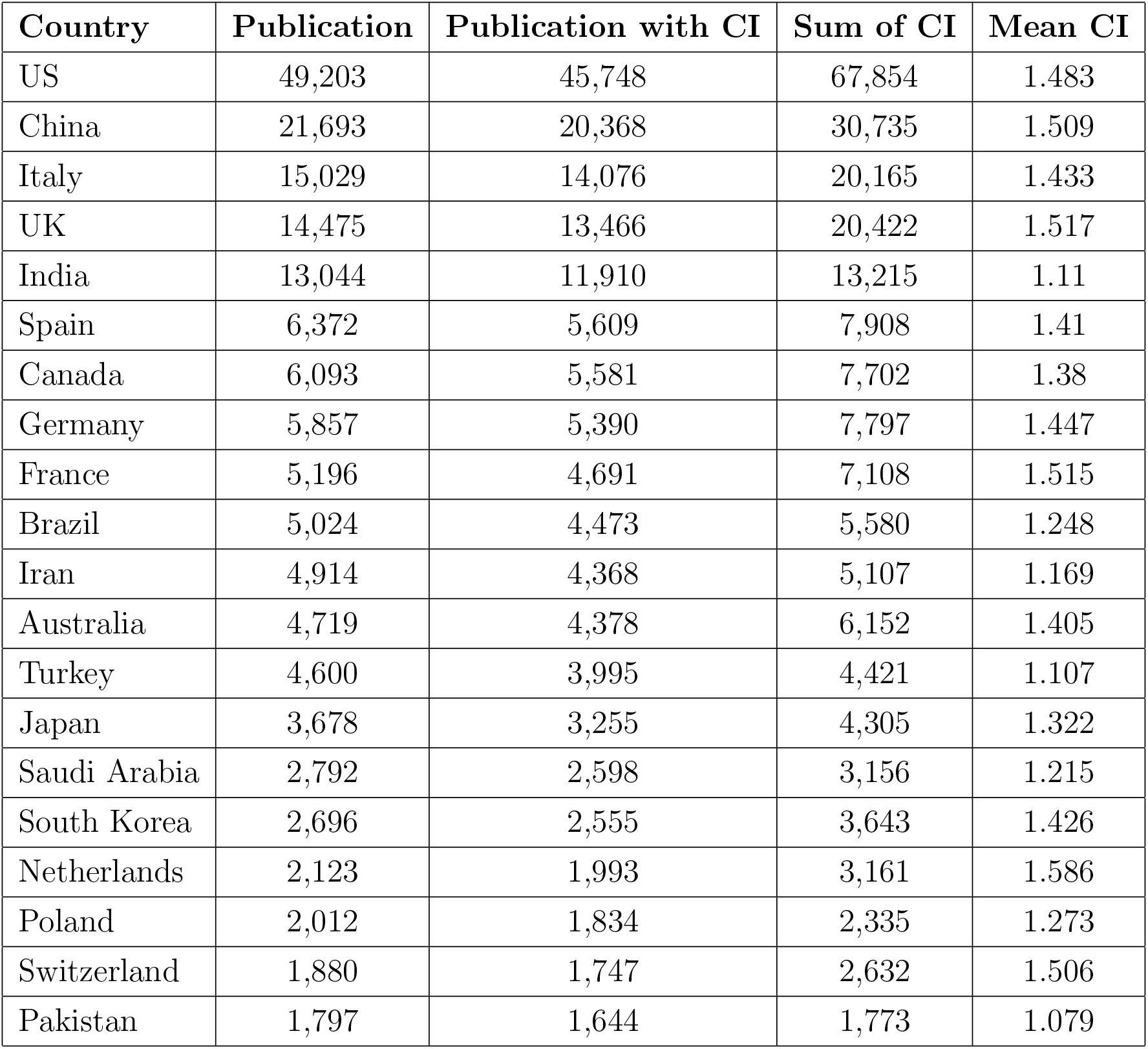
Top-20 countries with the highest cumulative Composite Indicator (CI)

| **Country** | **Publication** | **Publication with CI** | **Sum of CI** | **Mean CI** |
| --- | --- | --- | --- | --- |
| US | 49,203 | 45,748 | 67,854 | 1.483 |
| China | 21,693 | 20,368 | 30,735 | 1.509 |
| Italy | 15,029 | 14,076 | 20,165 | 1.433 |
| UK | 14,475 | 13,466 | 20,422 | 1.517 |
| India | 13,044 | 11,910 | 13,215 | 1.11 |
| Spain | 6,372 | 5,609 | 7,908 | 1.41 |
| Canada | 6,093 | 5,581 | 7,702 | 1.38 |
| Germany | 5,857 | 5,390 | 7,797 | 1.447 |
| France | 5,196 | 4,691 | 7,108 | 1.515 |
| Brazil | 5,024 | 4,473 | 5,580 | 1.248 |
| Iran | 4,914 | 4,368 | 5,107 | 1.169 |
| Australia | 4,719 | 4,378 | 6,152 | 1.405 |
| Turkey | 4,600 | 3,995 | 4,421 | 1.107 |
| Japan | 3,678 | 3,255 | 4,305 | 1.322 |
| Saudi Arabia | 2,792 | 2,598 | 3,156 | 1.215 |
| South Korea | 2,696 | 2,555 | 3,643 | 1.426 |
| Netherlands | 2,123 | 1,993 | 3,161 | 1.586 |
| Poland | 2,012 | 1,834 | 2,335 | 1.273 |
| Switzerland | 1,880 | 1,747 | 2,632 | 1.506 |
| Pakistan | 1,797 | 1,644 | 1,773 | 1.079 |

### 5.5 Global publication impact distribution per H5-index

Here, we present the distribution of the global publication impact in terms of the cumulative H5-index values of each first-authored country’s publications in world map. Only those publications with H5-index values are counted here.

Figure 5 shows the sum of the H5-index values of all publications for each first-authored country over all countries with the H5-index values on their publications. The H5-index values of journal and conference publications were extracted from Google Scholar, as shown in Section 2.3. 173 countries except Guinea-bissau and Vanuatu are included in this result. The full results can be found in “Global research impact distribution per H5-index.xlsx.”

**Figure 5:**
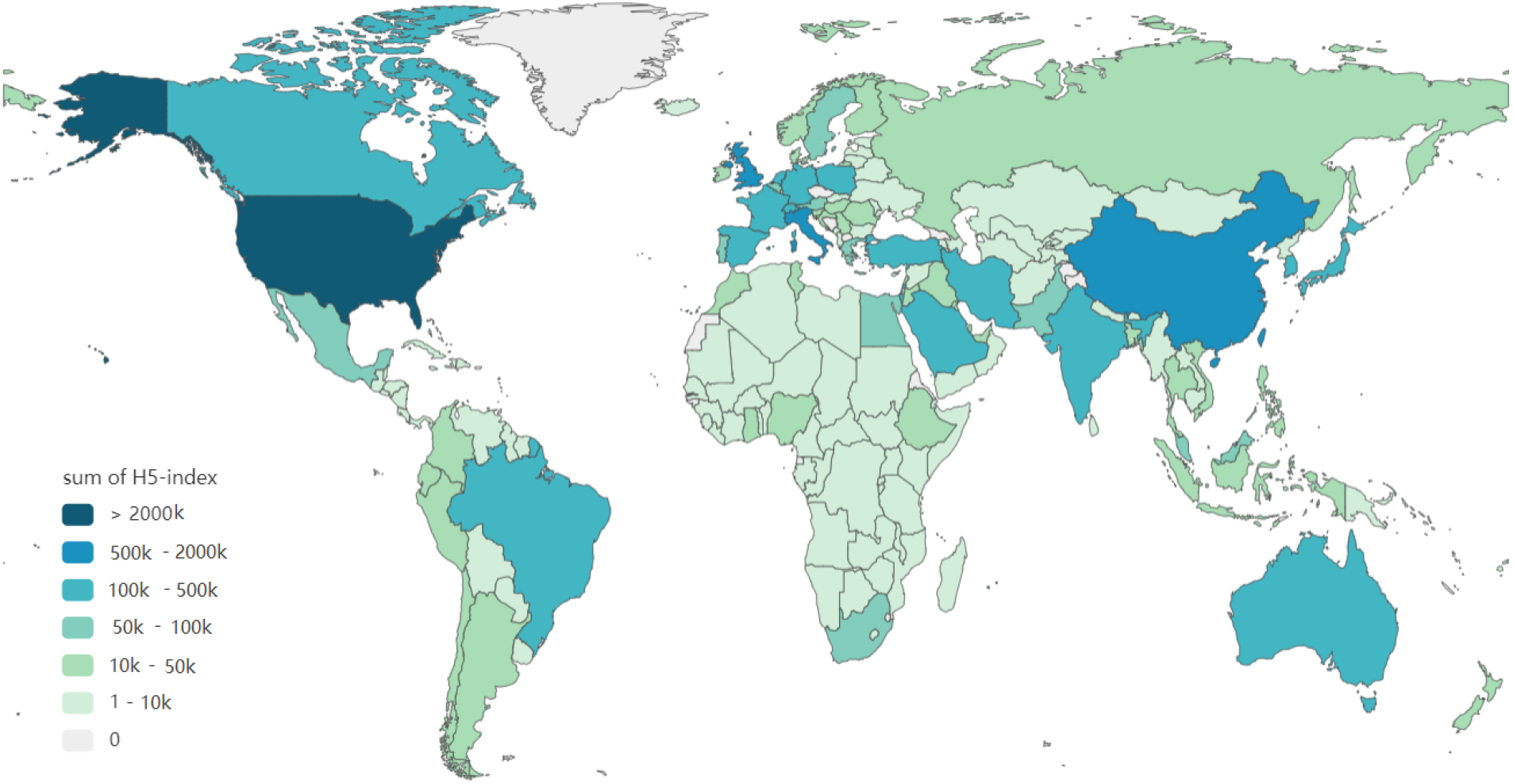
Global publication impact distribution per cumulative H5-index

The top-10 countries in terms of their cumulative H5-index are the US, China, the UK, Italy, India, Germany, Spain, Canada, France and Australia.

### 5.6 Global publication impact distribution per Impact Factor

Here, the distribution of the global publication impact in terms of the cumulative impact factor values of all first-authored publications from each country is presented in world map. Only those publications with the impact factor values on their publication venues are counted here.

Figure 6 shows the sum of the impact factor values of all publications for each first-authored country over all countries with their data available. The impact factor values of journal publications were obtained from Web of Science, as shown in Section 2.3. There are 173 countries except S. Sudan and Vanuatu displayed in the figure. The full results can be found in “Global research impact distribution per Impact Factor.xlsx.”

**Figure 6:**
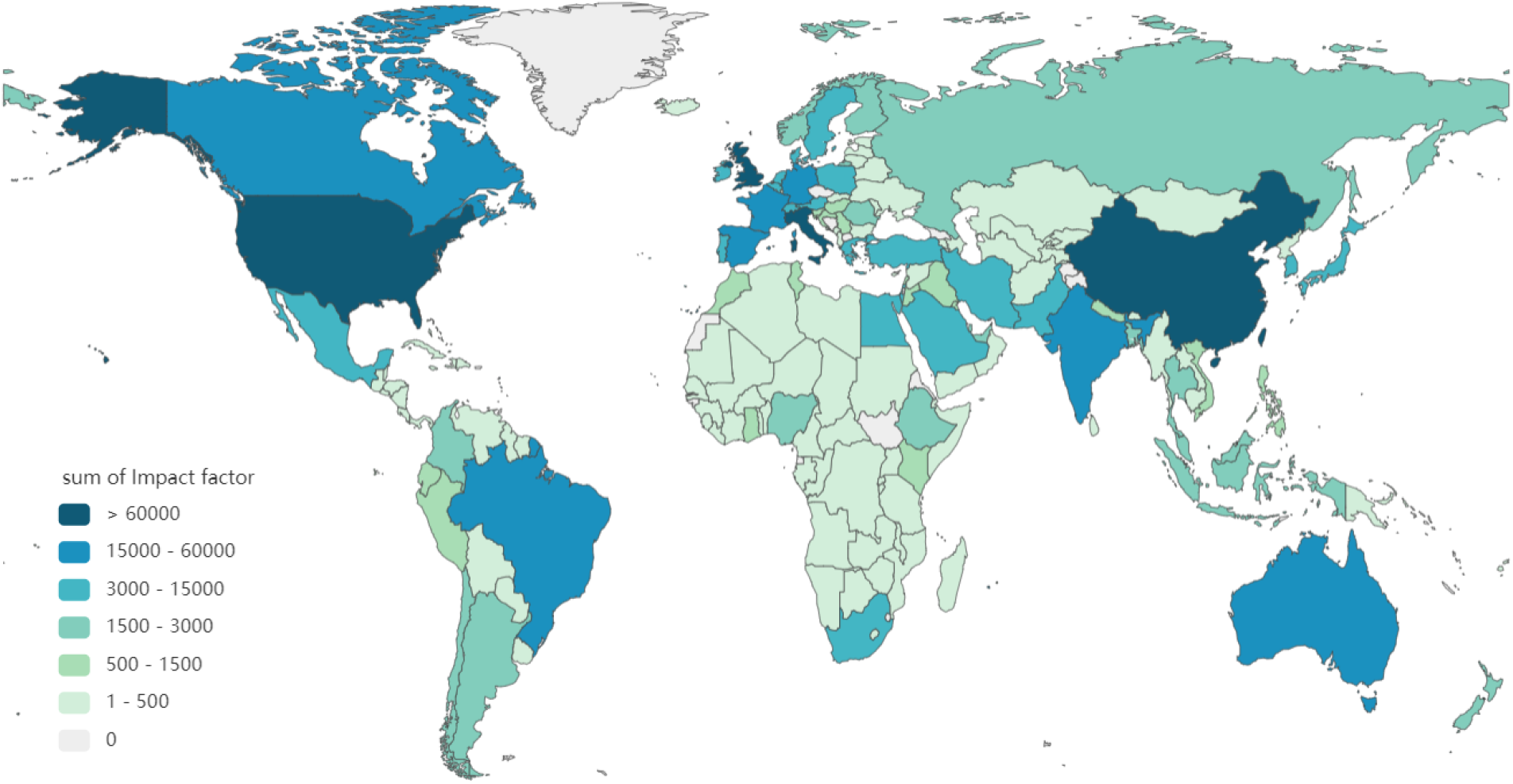
Global publication impact distribution per cumulative Impact Factor

The top-10 countries in terms of their cumulative impact factor are the US, China, the UK, Italy, Germany, Canada, France, India, Spain and Australia.

### 5.7 Global publication impact distribution per CiteScore

Here, the distribution of the global publication impact in terms of the cumulative CiteScore values of all first-authored publications from each country is presented in world map. Only those publications with the CiteScore values on their publication venues are counted here.

Figure 7 presents the sum of the CiteScore values of all publications for each first-authored country over all countries. The CiteScore values of journal and conference publications were obtained from Scopus, as shown in Section 2.3. 174 countries except Vanuatu are displayed here. The full results can be found in “Global research impact distribution per CiteScore.xlsx.”

**Figure 7:**
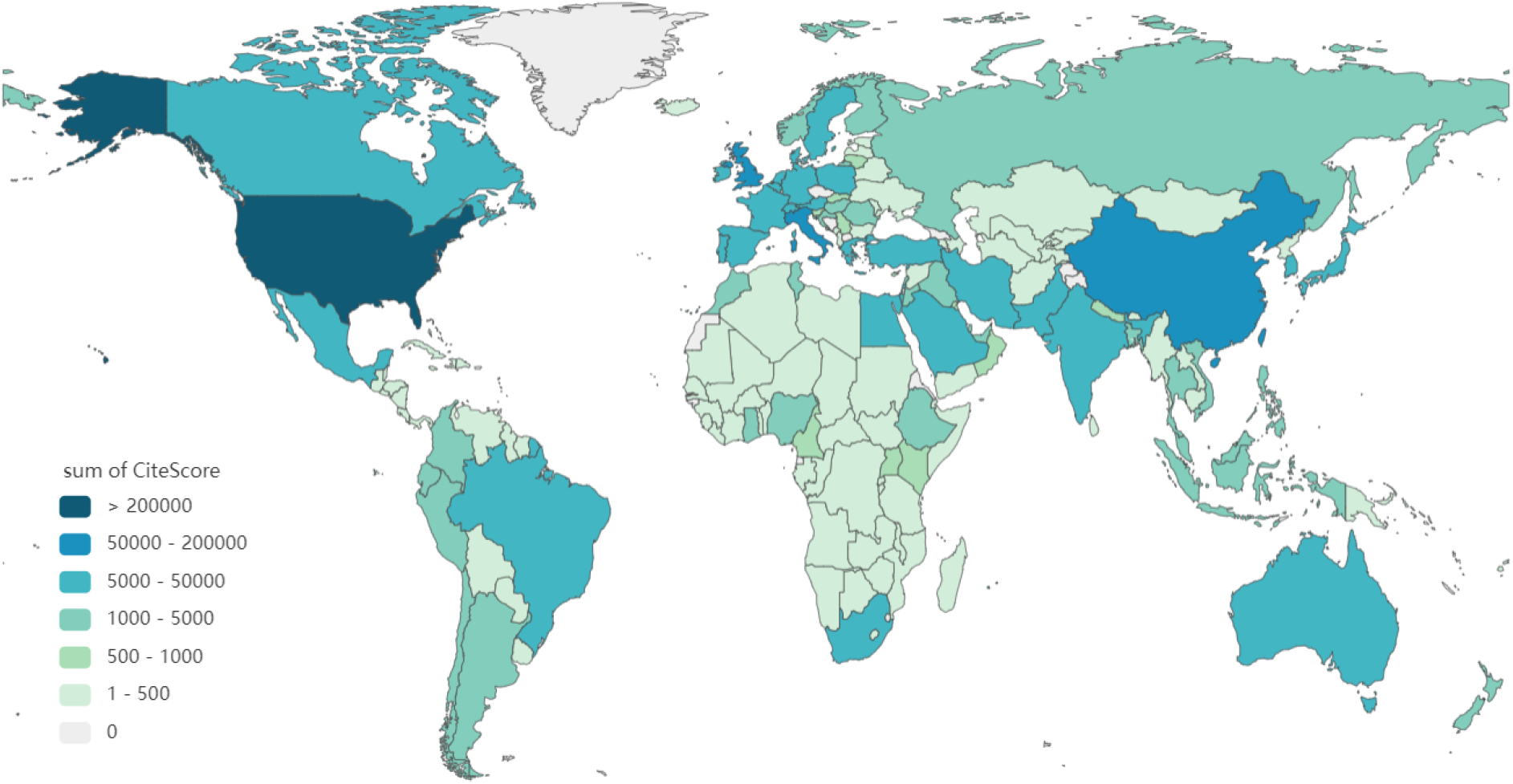
Global publication impact distribution per cumulative CiteScore

The top-10 countries in terms of their cumulative CiteScore are the US, China, the UK, Italy, India, Germany, France, Canada, Spain and Australia.

### 5.8 Global publication impact distribution per SNIP

Here, the distribution of the global publication impact in terms of the cumulative SNIP values of all first-authored publications from each country is presented in world map. Only those publications with the SNIP values on their publication venues are counted here.

Figure 8 presents the sum of the SNIP values of all publications for each first-authored country over all countries with data available. The SNIP values of journal and conference publications were obtained from Scopus, as shown in Section 2.3. 174 countries except Vanuatu are displayed in the figure. The full results can be found in “Global publication impact distribution per SNIP.xlsx.”

**Figure 8:**
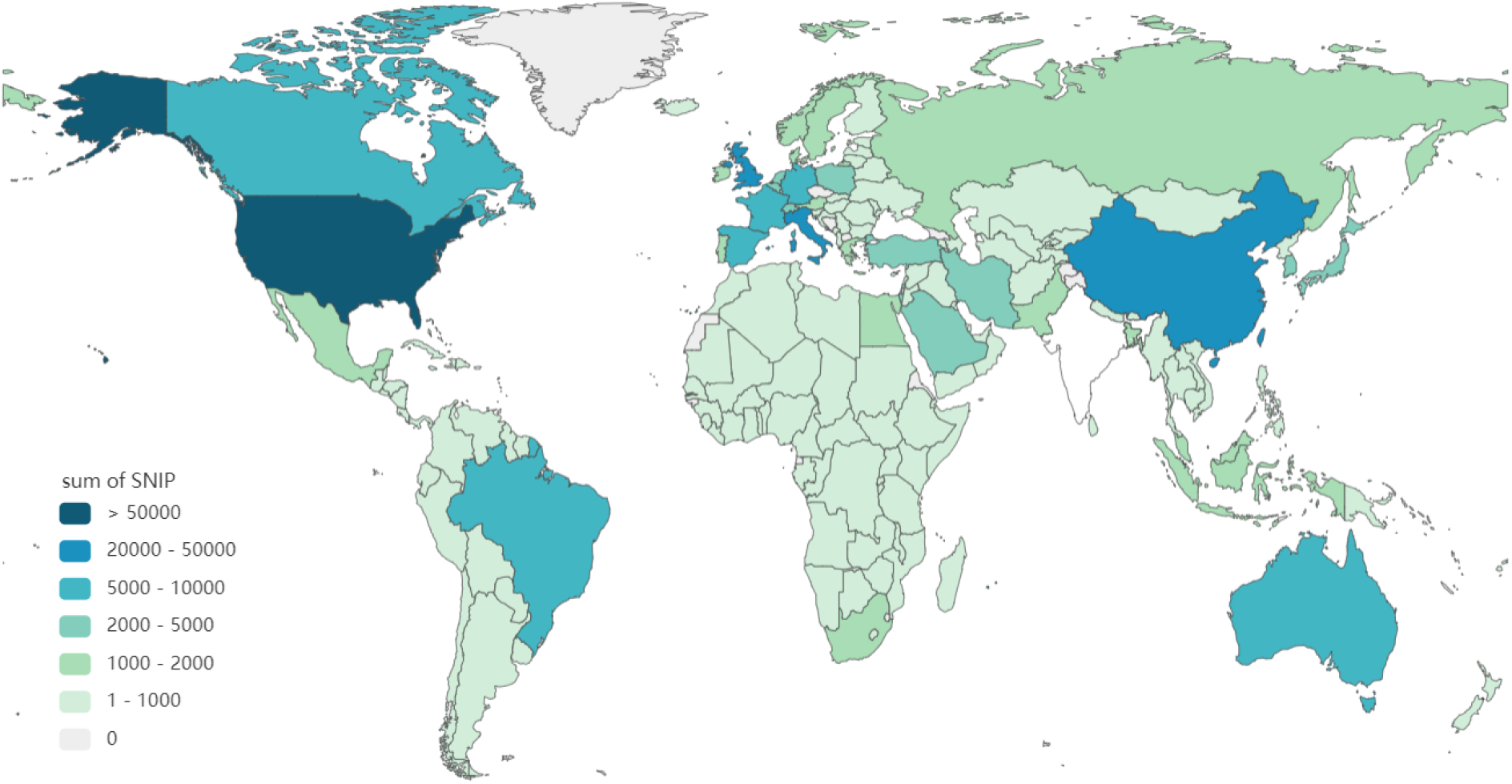
Global publication impact distribution per cumulative SNIP

The top-10 countries in terms of their cumulative SNIP are the US, China, the UK, Italy, India, Canada, Germany, Spain, France and Australia.

### 5.9 Global publication impact distribution per SJR

Here, the distribution of the global publication impact in terms of the cumulative SJR values of all first-authored publications from each country is presented in world map. Only those publications with the SJR values on their publication venues are counted here.

Figure 9 displays the sum of the SJR values of all publications for each first-authored country over all countries in world map. The SJR values of journal and conference publications were obtained from Scopus, as shown in Section 2.3. 173 countries except Andorra and Vanuatu are displayed in this result. The full results can be found in “Global publication impact distribution per SJR.xlsx.”

**Figure 9:**
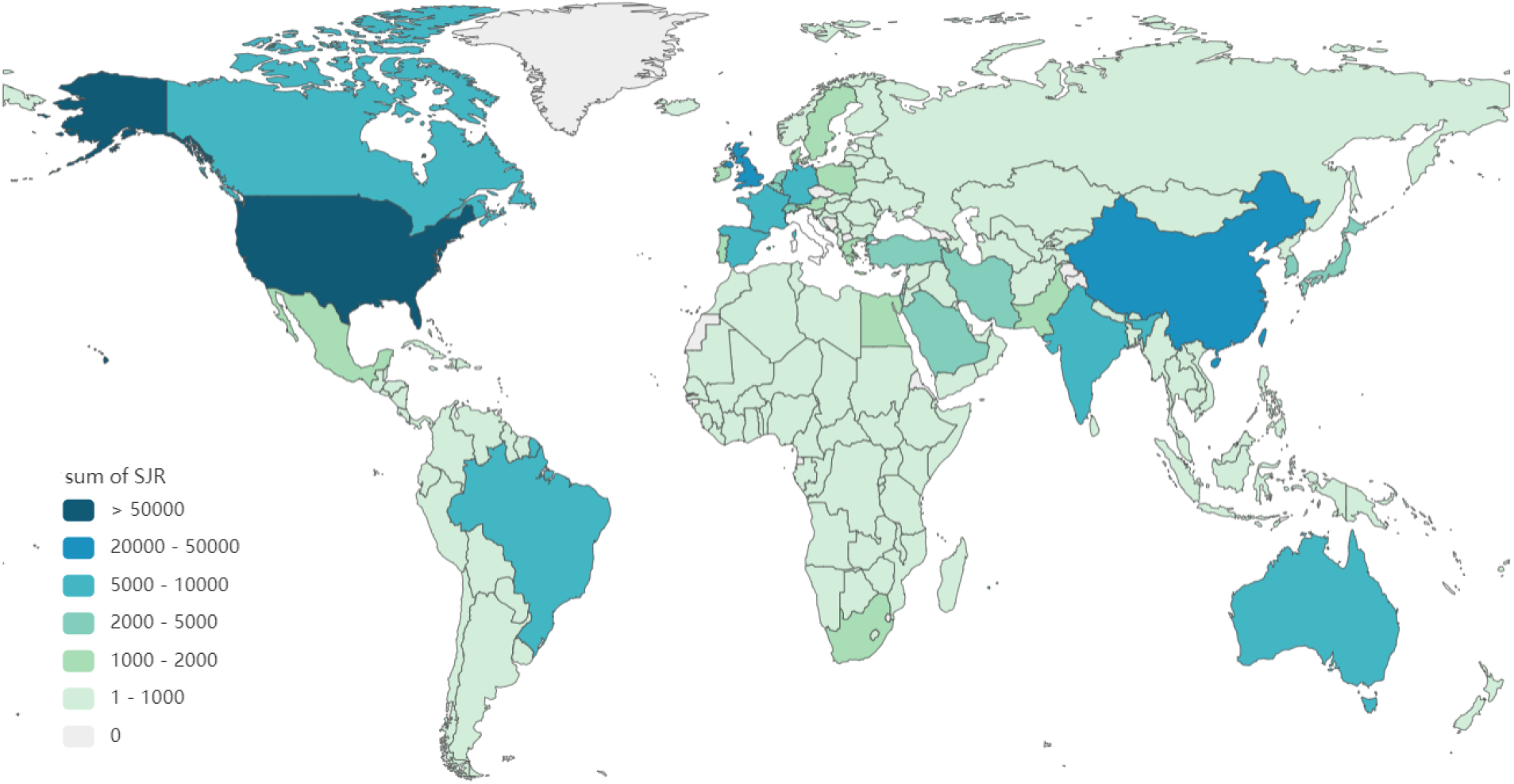
Global publication impact distribution per cumulative SJR

The top-10 countries in terms of their cumulative SJR are the US, China, the UK, Italy, Germany, India, France, Canada, Spain and Australia.

### 5.10 Top-10 published country’s research impact

Here, we present the research impact of the top-10 first-authored countries with the highest number of publications. The publication impact metrics are H5-index, Impact Factor, CiteScore, SNIP, SJR, and CI. We calculate their cumulative values for each country. To make the impact metrics of publications comparable between countries, we normalize each impact metric and then sum its values of all publications from each first-authored country as the overall publication impact of this country per the impact metric. The log normalization of these impact metrics is introduced in Section 4.1.

Figure 10 shows the top-10 mostly published country’s research impact. There are two Y axes, the right Y axis with line graphs stands for the publication-averaged mean of the log-normalized impact metrics of the top-10 countries, the log-normalized impact metrics of each impact metric is calculated per Equation (1). The left Y axis refers to the cumulative log-normalized impact metric of each country, presented in six colored bar charts for the sum of log-normalization of H5-index, Impact Factor, CiteScore, SNIP, SJR and CI, respectively.

**Figure 10:**
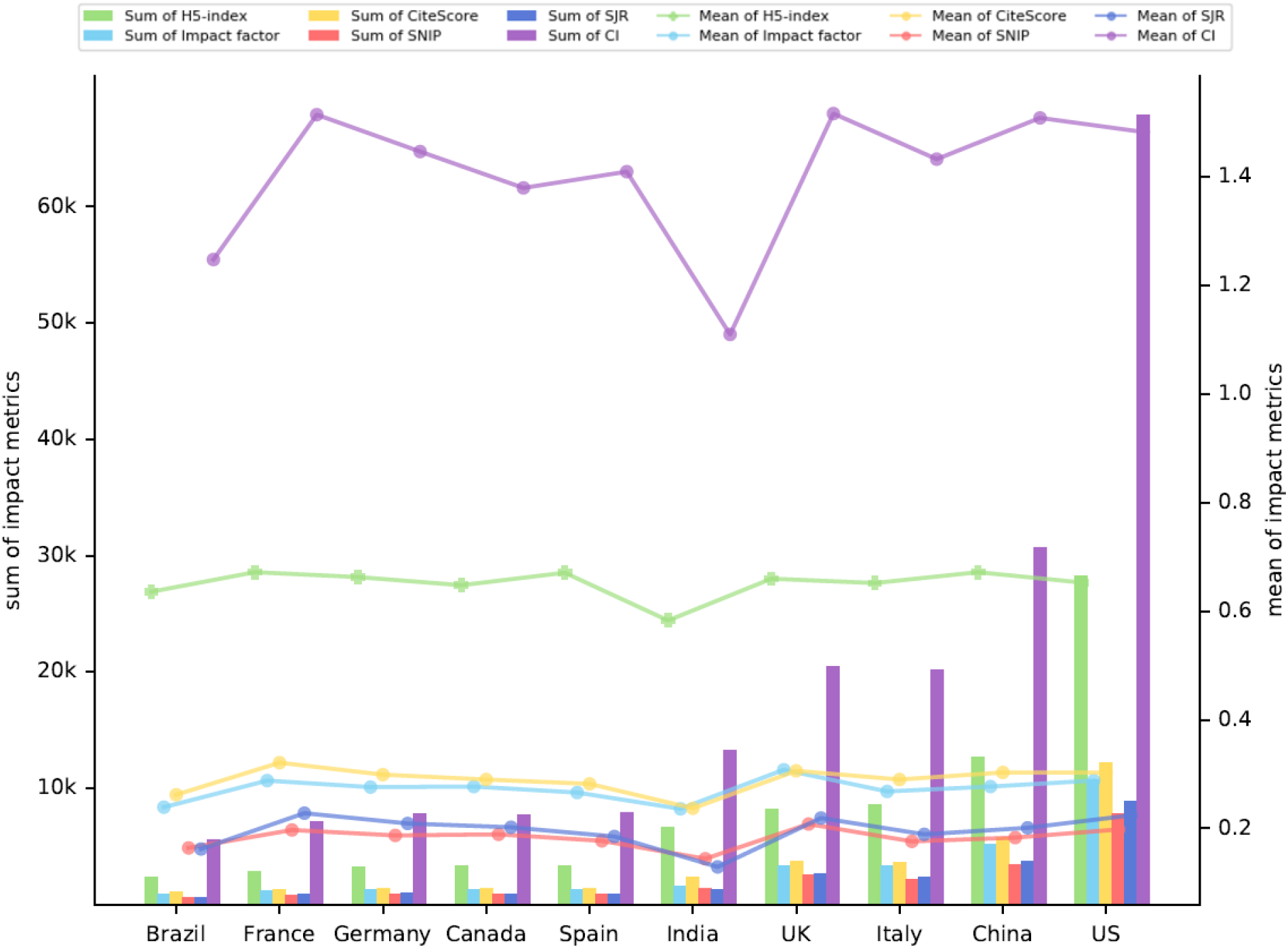
Top-10 mostly published country’s research impact

The full details can be found in “Global research impact.xlsx.”

The top-10 mostly published countries are the US, China, Italy, the UK, India, Spain, Canada, Germany, France, and Brazil. In terms of the cumulative (sum of) H5-index, the US, China, the UK, Italy and India rank the top-5 respectively; the US, China, the UK, Italy and Germany rank the top-5 in terms of sum and mean of Impact Factor; the US, China, the UK, Italy and India rank top-5 in terms of sum and mean of CiteScore; the US, China, the UK, Italy and India rank top-5 in terms of sum and mean of SNIP; and the US, China, the UK, Italy and Germany rank top-5 in terms of sum and mean of SJR. With regard to the cumulative composite indicator (CI), the US, China, the UK, Italy and India rank top 5, in contrast, the UK, France, China, the US and Germany rank top 5 in terms of mean CI.

### 5.11 Top-10 published country’s disciplinary research impact

Similar to the presentation in Section 5.10, here, we present the results of the top-10 published country’s publication impact in terms of three major disciplines: computer science, social science, and medical science. The visualization settings are the same as that in Section 5.10.

Figure 11 shows the publication research impact of Top-10 countries in terms of their disciplinary publications on COVID-19. The full details can be found in “Global disciplinary research impact.xlsx.”

**Figure 11:**
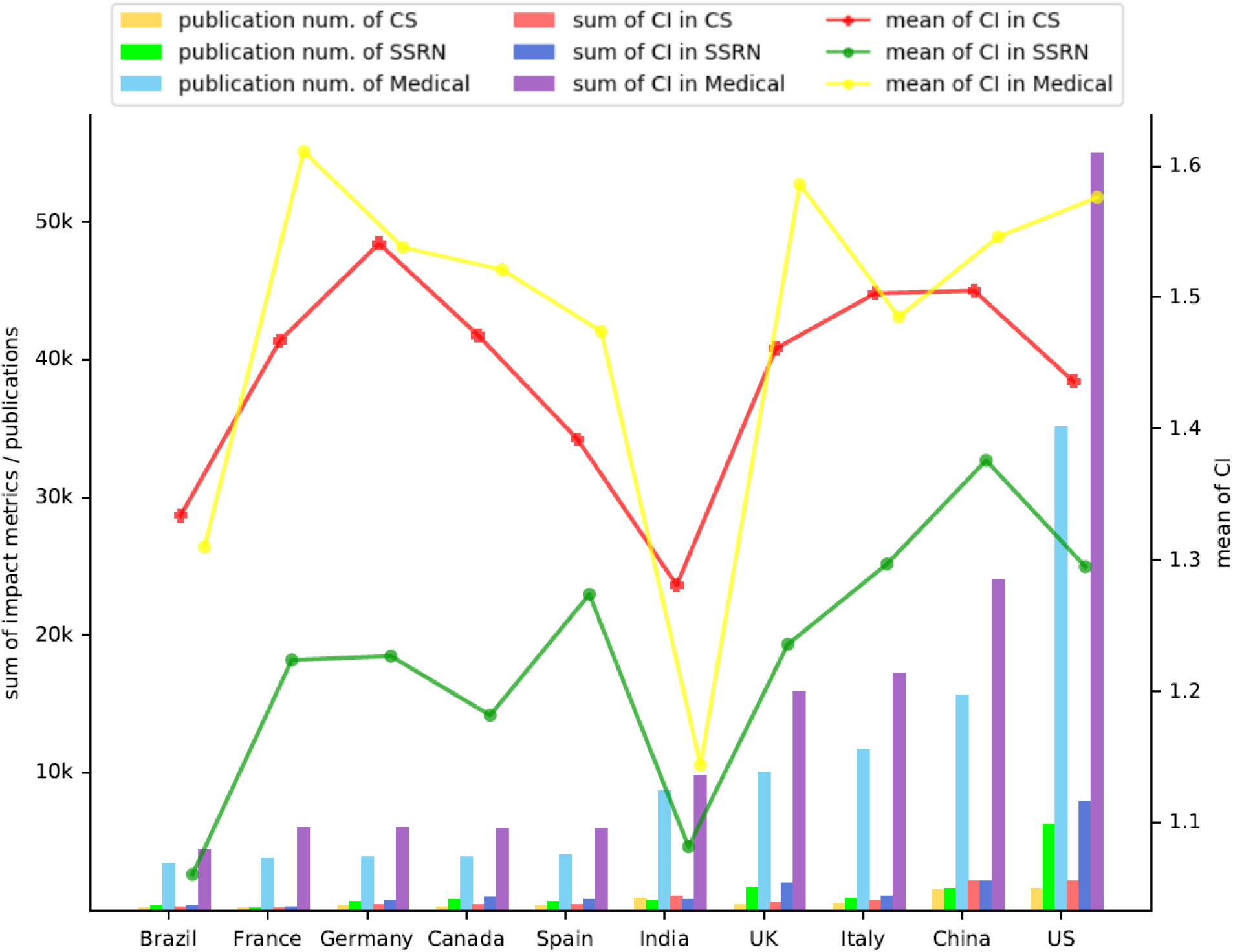
Top-10 mostly published country’s disciplinary research impact

### 5.12 Global research collaborations

Here, we discuss the research collaborations between countries on COVID-19. Centered by the first-authored country of each publication, each two-author pairwise collaboration shows a two-author pair from different countries. We thus obtain the country-collaboration pair of N countries in the form of 1-2, 1-3, …, 1-N, if a publication has authors from N different countries. The country of the first author’s affiliation is regarded as the publication’s host country, which must be available from the publication data.

In the global publications, there are 189 countries formed research collaborations on COVID-19. Figures 12, 13 and 14 show some snapshots of such research collaborations with collaboration times greater than 50, 200, and 500, respectively. Here, “greater than N” (N = 50, 200, 500) refers to those jointly authored publications with at least N co-author pairs from different countries, i.e., N collaborative publications.

**Figure 12:**
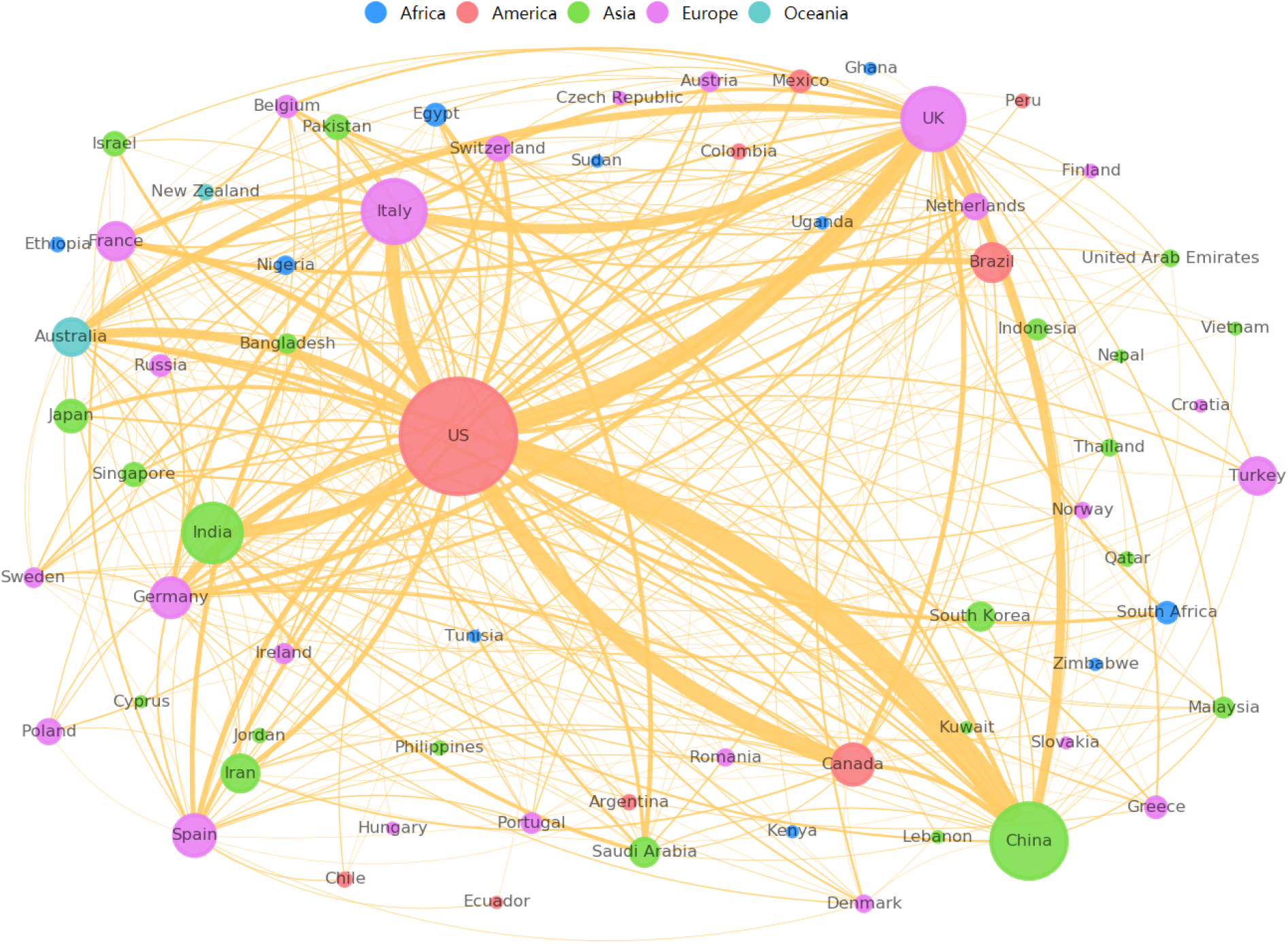
Cross-country COVID-19 research collaboration with joint publications >= 50 (the networking graph shows 67 countries have collaborative publications >= 50.)

**Figure 13:**
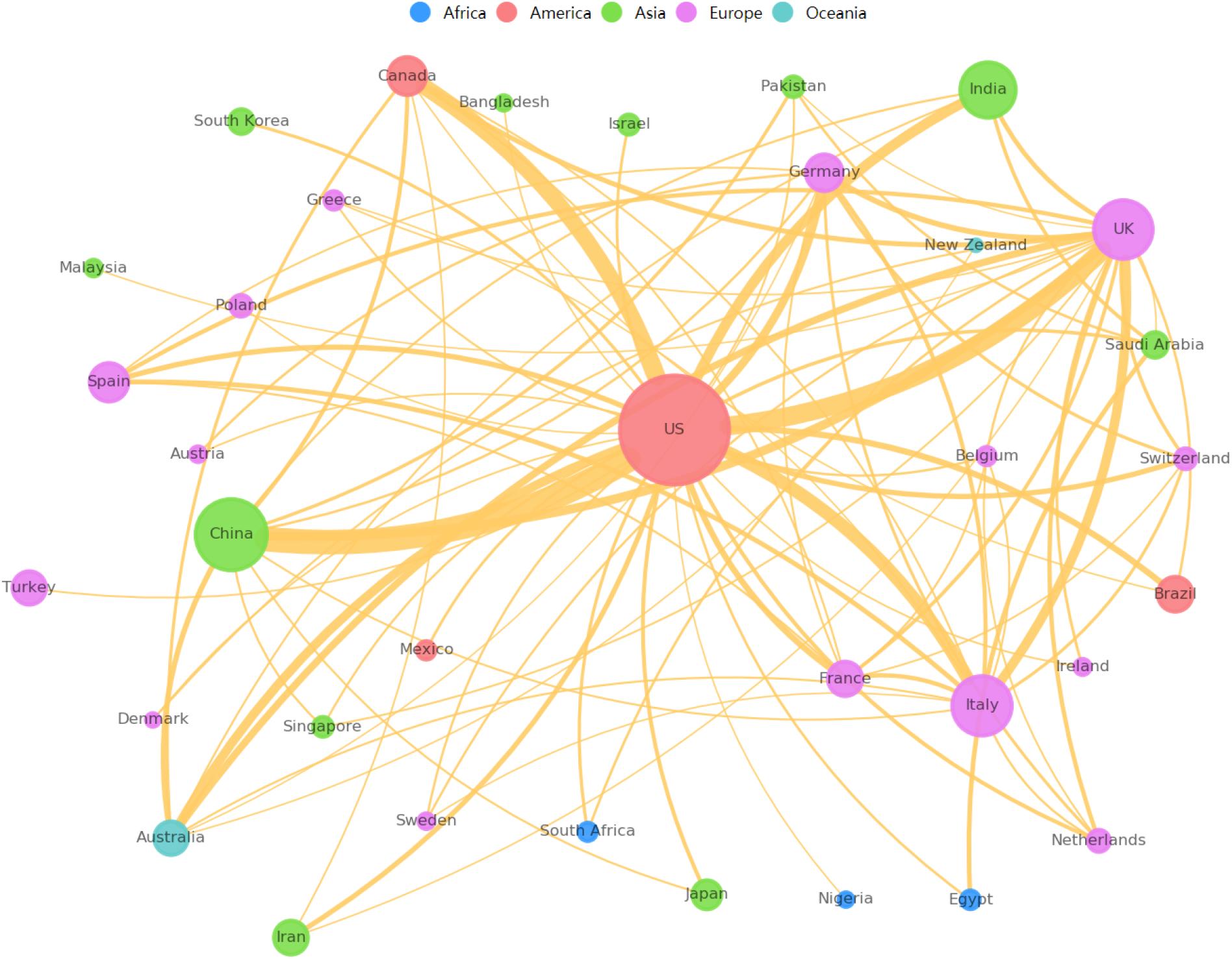
Cross-country COVID-19 research collaborations >= 200 (the networking graph shows 35 countries have collaboration publications >= 200.)

**Figure 14:**
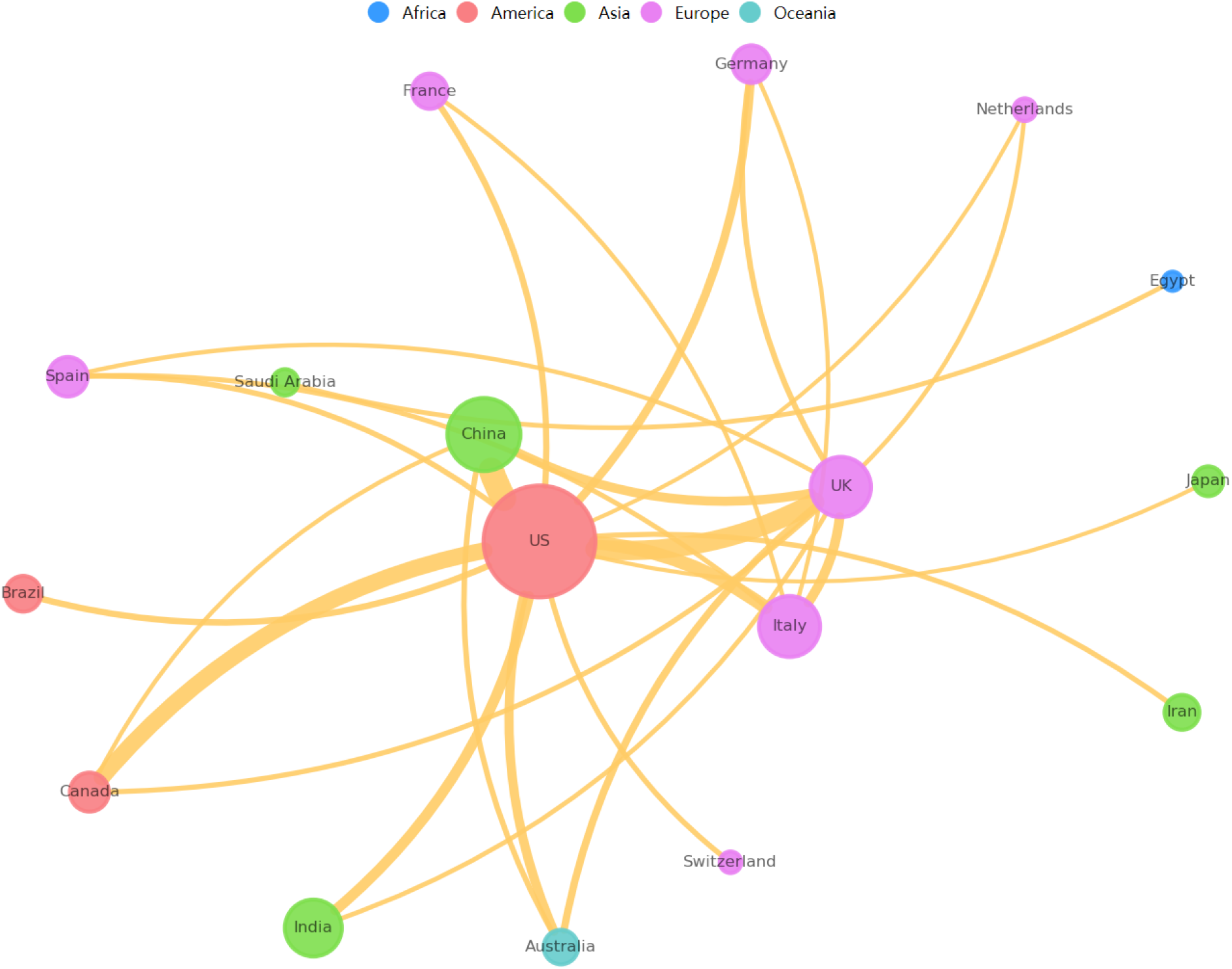
Cross-country COVID-19 research collaborations >= 500 (the networking graph shows 17 countries have collaboration publications >= 500.

In the network graph, different colors stand for countries from different continents. We only consider the pairwise collaborations between the first author and another author from a different country in a publication. Coauthoring from the same country is ignored, we also ignore the pairwise collaborations between non-first authors, such as the second-third author pair, the third-fourth author pair even if they are from different countries. The edge thickness represents the joint publication number (i.e., collaboration times) between countries. The node size stands for a country’s collaborative publication number.

The research collaboration results show that the US, China, the UK, Italy and India are the top-5 mostly collaborative countries on COVID-19. The US appears to be the center point of the global COVID-19 research collaboration network among their top-5 collaboration countries. Of all publications, the US had 3,356 collaborations with China, 2,570 with UK, 1,788 with Italy and 1,397 with India, respectively. The results can be found in sheet named ‘Collab. without author’s order’ in the file ‘Global research collaborations.xlsx.’

Similarly, of the 1397 publications with UK as the first authored country, 1200 collaborations with the Canada, 1,044 with China and 718 with Italy, as the US’ top-5 collaborative countries. Of all publications, China had 2,312 collaborations with the US, 868 with UK, 538 with Australia and 368 with Canada, respectively, as China’s top-5 research collaborators on COVID-19. The results can be found in the sheet named ‘Collab. with author’s order’ of the file ‘Global research collaborations.xlsx.’

## 6 G20 and OECD publication profile

G20 countries and regions^41^ contributed 182,097 publications to studying the COVID-19 pandemic in total. In contrast, 38 OECD countries and regions^42^ contributed 139,588 publications. Here, we analyze their publication impact in terms of all of their publications, disciplinary publications, and publication-averaged impact, respectively.

### 6.1 G20 country’s overall publication impact

Here, we analyze the G20 country’s COVID-19 research quality in terms of the five impact metrics of all of their publications in each country. We collected the publications of each G20 country, and then calculated their impact metrics for each country.

Figure 15 shows the log-normalized cumulative values of H5-index, Impact Factor, and CiteScore of each country in terms of three dimensions. The X axis stands for the logarithm of a country *i*’s sum of CiteScore of all of their publications *N*_*i*_, which is calculated by *ln*(*sum*(*CiteScore*_*i*_ +1)). Similarly, Y axis stands for the logarithm of a country’s sum of Impact Factor values of all publications, which is represented by *ln*(*sum*(*Impactfactor*_*i*_ + 1)). Z axis stands for the logarithm of a country’s sum of H5-index values of all publications, which is represented by *ln*(*sum*(*H*5 *− index*_*i*_ + 1)). The ball size stands for the logarithm value of a country’s publication number *N*_*i*_, which is represented by *ln*(*sum*(*N*_*i*_ + 1)). The data can be found in “G20 country’s overall research impact.xlsx.”

**Figure 15:**
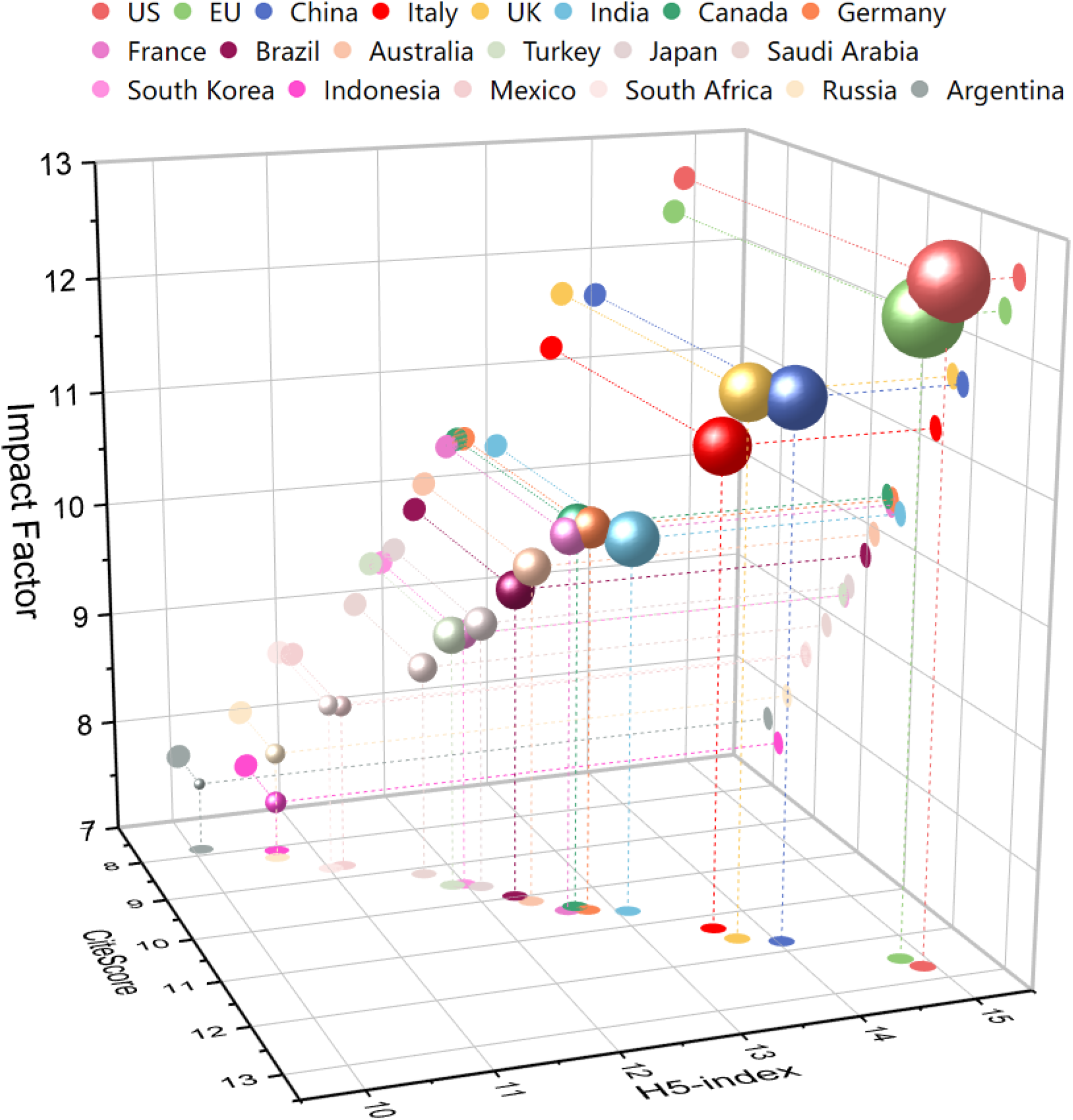
G20 country’s overall COVID-19 research impact

The results show that the US, EU, China, Italy and the UK made the top-5 impact in the COVID-19 research. The top-5 countries in terms of H5-index are the US, EU, China, the UK, and Italy. The top-5 countries in terms of Impact Factor are the US, EU, China, the UK and Italy. The US, EU, China, the UK and Italy rank the top-5 countries in terms of CiteScore.

### 6.2 G20 country’s publication impact in computer science

Here, we analyze the research impact of G20 country’s publications in computer science. Taking the same analysis approach in Section 6.1, we collected those computer science-related publications for each country and then calculated their log-normalized cumulative impact metrics.

Figure 16 shows the results of the G20 country’s COVID-19 research impact in terms of the three impact metrics: CiteScore, Impact Factor, and H5-index in the discipline of computer science. The meaning of each axis is the same as in Section 6.1. The data can be found in “G20 country’s research impact in computer science.xlsx.”

**Figure 16:**
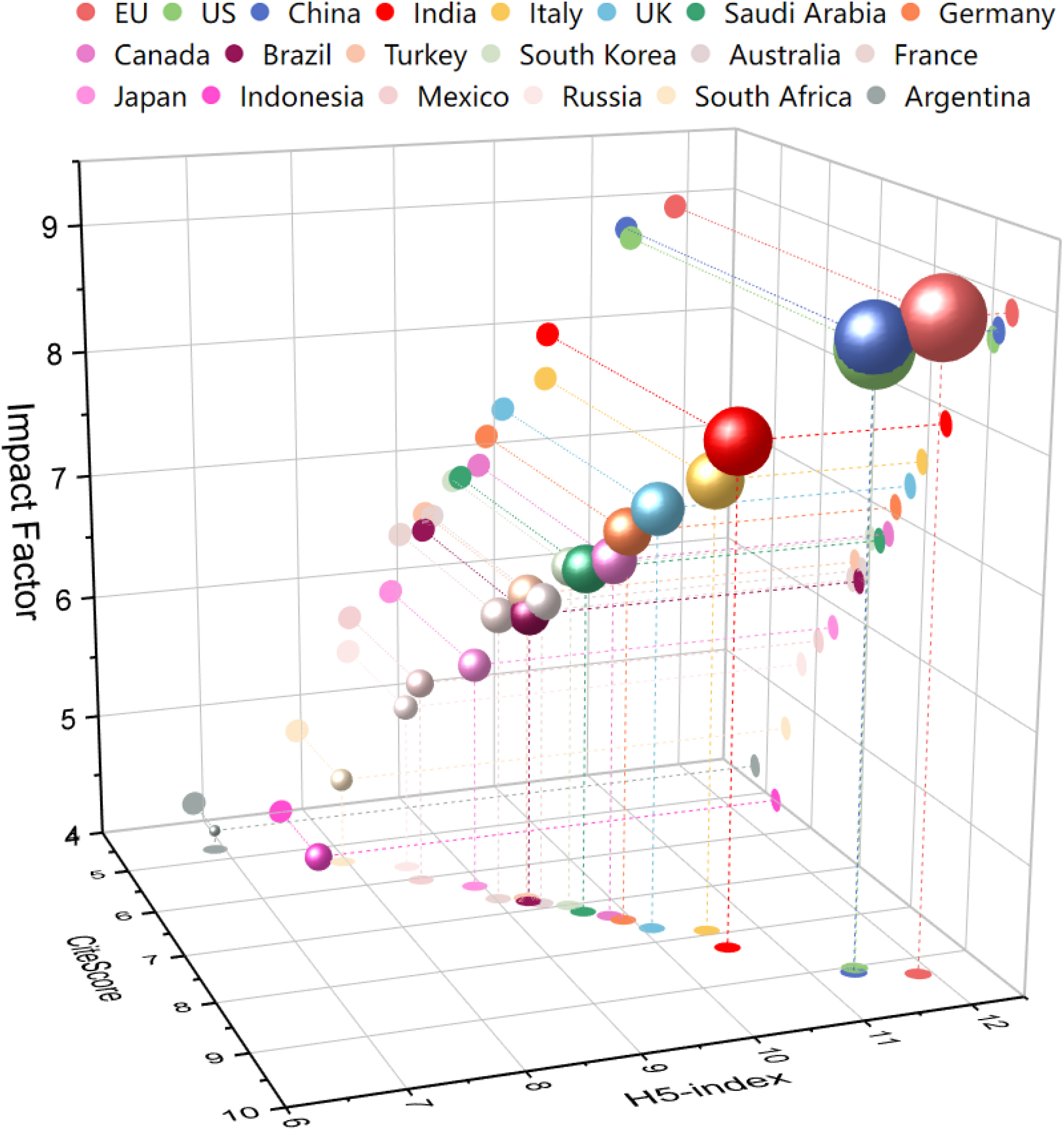
G20 country’s research impact of computer science publications

Table 9 further lists the details of the data shown in Figure 16. *Num_pub* refers to the number of publications in each country.

**Table 9:** G20 country’s research impact of computer science publications

| **Country** | **Ln**<br>**(sum(CiteScore)**<br>**+1)** | **Ln**<br>**(sum(H5-index)**<br>**+1)** | **Ln**<br>**(sum(Impact factor)**<br>**+1)** | **Num_**<br>**pub** |
| --- | --- | --- | --- | --- |
| US | 8.99 | 11.39 | 8.62 | 1629 |
| China | 9.14 | 11.42 | 8.76 | 1504 |
| India | 8.27 | 10.53 | 7.85 | 860 |
| Italy | 7.87 | 10.49 | 7.49 | 504 |
| UK | 7.65 | 10.05 | 7.24 | 387 |
| Saudi Arabia | 7.16 | 9.64 | 6.71 | 314 |
| Germany | 7.41 | 9.88 | 7.02 | 301 |
| Canada | 7.29 | 9.79 | 6.79 | 286 |
| Brazil | 6.76 | 9.23 | 6.28 | 196 |
| Turkey | 6.7 | 9.25 | 6.42 | 179 |
| South Korea | 6.95 | 9.54 | 6.68 | 175 |
| Australia | 6.83 | 9.32 | 6.39 | 164 |
| France | 6.66 | 9 | 6.28 | 143 |
| Japan | 6.28 | 8.9 | 5.78 | 121 |
| Mexico | 6.02 | 8.49 | 5.62 | 89 |
| Indonesia | 5.06 | 7.73 | 3.91 | 84 |
| Russia | 5.68 | 8.48 | 5.31 | 70 |
| South Africa | 5.36 | 7.92 | 4.69 | 57 |
| Argentina | 4.65 | 6.81 | 4.09 | 20 |
| European Union | 9.26 | 11.83 | 8.85 | 2104 |

In computer science, the top-5 countries in terms of H5-index are EU, the US, China, India and Italy. The top-5 countries in terms of Impact Factor are EU, China, the US, India and Italy. EU, China, the US, India and Italy rank the top-5 countries in terms of CiteScore.

### 6.3 G20 country’s publication impact in medical science

Similar to the analysis undertaken in Section 6.2, here, we analyze the research impact of each G20 country’s publications in medical science.

Figure 17 shows the statistics of the G20 country’s COVID-19 research quality in terms of CiteScore, Impact Factor, and H5-index of their publications in the category of medical science. The meaning of each axis is the same as in Figure 15. Table 10 further lists the data details of Figure 17. The data can be found in “G20 country’s research impact in medical science.xlsx.”

**Figure 17:**
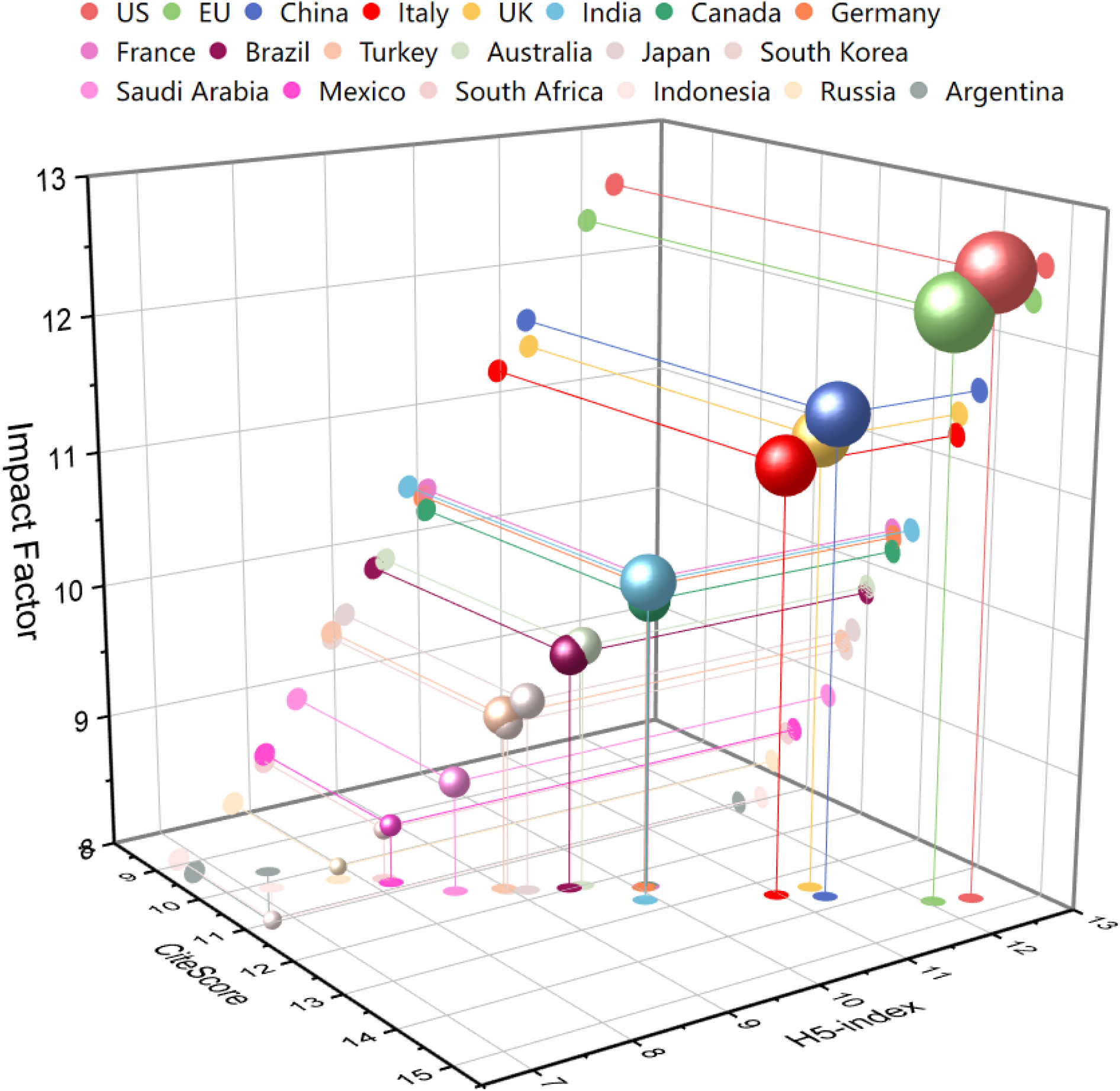
G20 country’s research impact of medical science publications

**Table 10:**
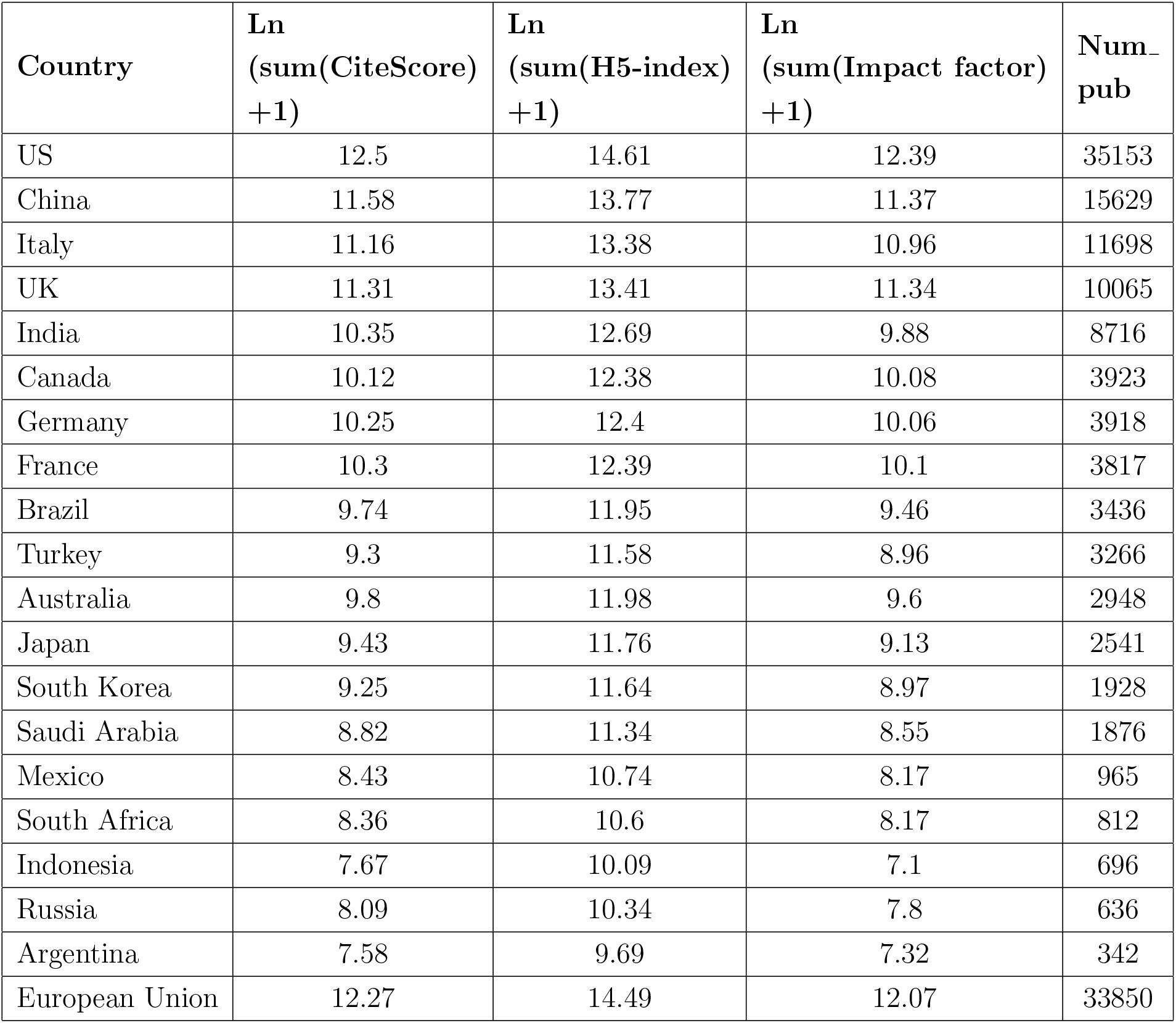
G20 country’s research impact of medical science publications

| **Country** | **Ln**<br>**(sum(CiteScore)**<br>**+1)** | **Ln**<br>**(sum(H5-index)**<br>**+1)** | **Ln**<br>**(sum(Impact factor)**<br>**+1)** | **Num_**<br>**pub** |
| --- | --- | --- | --- | --- |
| US | 12.5 | 14.61 | 12.39 | 35153 |
| China | 11.58 | 13.77 | 11.37 | 15629 |
| Italy | 11.16 | 13.38 | 10.96 | 11698 |
| UK | 11.31 | 13.41 | 11.34 | 10065 |
| India | 10.35 | 12.69 | 9.88 | 8716 |
| Canada | 10.12 | 12.38 | 10.08 | 3923 |
| Germany | 10.25 | 12.4 | 10.06 | 3918 |
| France | 10.3 | 12.39 | 10.1 | 3817 |
| Brazil | 9.74 | 11.95 | 9.46 | 3436 |
| Turkey | 9.3 | 11.58 | 8.96 | 3266 |
| Australia | 9.8 | 11.98 | 9.6 | 2948 |
| Japan | 9.43 | 11.76 | 9.13 | 2541 |
| South Korea | 9.25 | 11.64 | 8.97 | 1928 |
| Saudi Arabia | 8.82 | 11.34 | 8.55 | 1876 |
| Mexico | 8.43 | 10.74 | 8.17 | 965 |
| South Africa | 8.36 | 10.6 | 8.17 | 812 |
| Indonesia | 7.67 | 10.09 | 7.1 | 696 |
| Russia | 8.09 | 10.34 | 7.8 | 636 |
| Argentina | 7.58 | 9.69 | 7.32 | 342 |
| European Union | 12.27 | 14.49 | 12.07 | 33850 |

In medical science, the top-5 countries in terms of H5-index are the US, the EU, China, the UK and Italy. The top-5 countries in terms of Impact Factor are the US, the EU, the UK, China and Italy. The US, the EU, China, the UK and Italy rank the top-5 countries in terms of CiteScore.

### 6.4 G20 country’s publication impact in social science

Similar to the analysis undertaken in Section 6.2, here, we analyze the research impact of each G20 country’s publications in social science.

Figure 18 shows the statistics of the G20 country’s COVID-19 research quality in terms of CiteScore, Impact Factor, and H5-index of their publications in the category of social science. The meaning of each axis is the same as in Figure 15. Table 11 further lists the data details of Figure 18. The data can be found in “G20 country’s research impact of social science.xlsx.”

**Figure 18:**
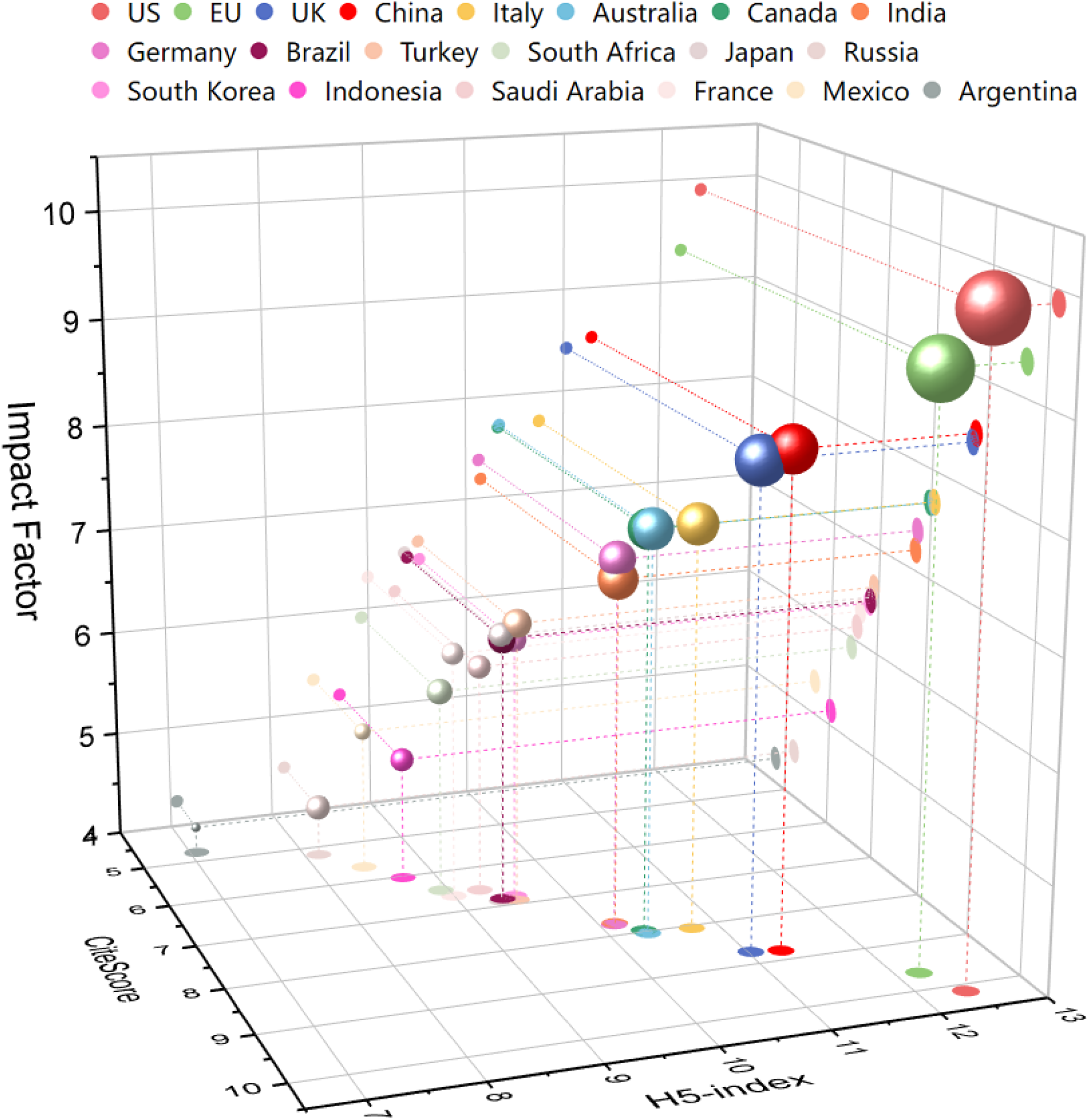
G20 country’s research impact of social science publications

**Table 11:** G20 country’s research impact of social science publications

| **Country** | **Ln**<br>**(sum(CiteScore)**<br>**+1)** | **Ln**<br>**(sum(H5-index)**<br>**+1)** | **Ln**<br>**(sum(Impact factor)**<br>**+1)** | **Num_**<br>**pub** |
| --- | --- | --- | --- | --- |
| US | 10.11 | 12.38 | 9.87 | 6217 |
| UK | 8.68 | 10.98 | 8.38 | 1639 |
| China | 8.82 | 11.33 | 8.53 | 1574 |
| Italy | 8.01 | 10.7 | 7.68 | 878 |
| Australia | 7.98 | 10.28 | 7.66 | 859 |
| Canada | 7.92 | 10.29 | 7.65 | 824 |
| India | 7.67 | 10.11 | 7.15 | 756 |
| Germany | 7.71 | 10.09 | 7.35 | 616 |
| Turkey | 6.89 | 9.51 | 6.62 | 354 |
| Brazil | 6.76 | 9.36 | 6.43 | 343 |
| Russia | 5.11 | 8.06 | 4.43 | 253 |
| Japan | 6.82 | 9.29 | 6.46 | 249 |
| South Africa | 6.38 | 8.86 | 5.88 | 241 |
| South Korea | 6.74 | 9.47 | 6.39 | 221 |
| Saudi Arabia | 6.52 | 9.22 | 6.1 | 208 |
| Indonesia | 5.92 | 8.63 | 5.14 | 196 |
| France | 6.56 | 8.93 | 6.24 | 192 |
| Mexico | 5.58 | 8.38 | 5.27 | 113 |
| Argentina | 4.75 | 7.04 | 4.25 | 49 |
| European Union | 9.58 | 12.15 | 9.25 | 4241 |

In social science, the top-5 countries in terms of H5-index are the US, the EU, China, the UK and Italy. The top-5 countries in terms of Impact Factor are the US, the EU, China, the UK and Italy. The US, the EU, China, the UK and Italy rank the top-5 countries in terms of CiteScore.

### 6.5 G20 country’s mean publication impact

Here, we further analyze the publication-averaged mean impact of each G20 country’s total publications in terms of the five major impact metrics: H5-index, Impact Factor, CiteScore, SNIP, and SJR. The mean publication impact of each impact metric is calculated per the method introduced in Section 4.1.

Figure 19 shows the statistics of the G20 countries’ research impact per publication in terms of the five impact metrics. We here apply the original impact metrics values of each publication to estimate the mean publication impact. The impact metrics value were crawled from different websites, as introduced in Section 2.3 on the research impact metrics collection. The data can be found in “G20 paper-averaged research impact metrics.xlsx.”

**Figure 19:**
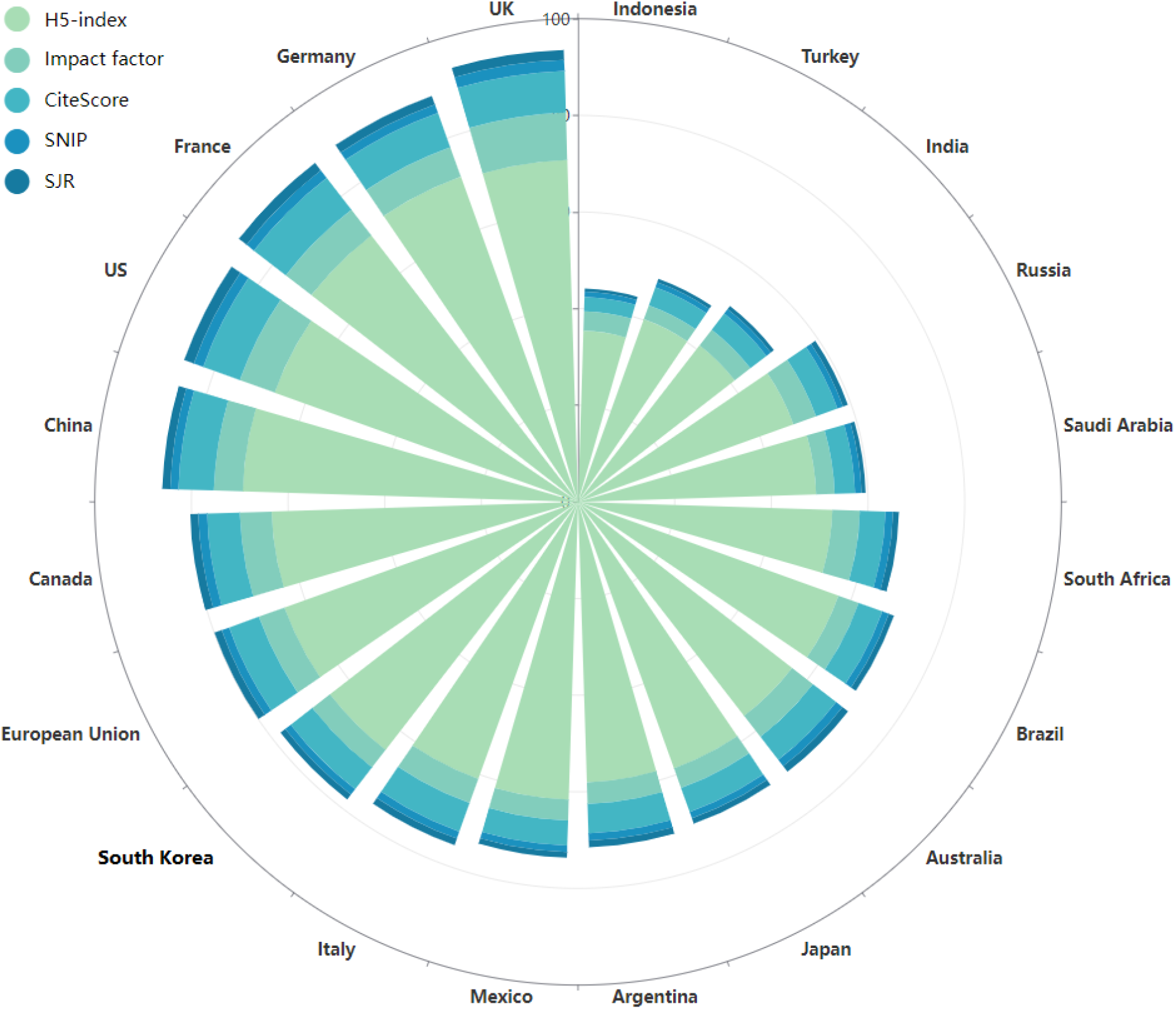
G20 publication-averaged mean publication impact

The results in Figure 19 show both the individual mean impact metrics and their accumulation for each country. Table 12 further shows the details of Figure 19 in terms of the mean impact metrics of the G20 countries globally.

**Table 12:** G20 publication-averaged mean publication impact

| **Country** | **Num_**<br>**pub** | **Mean**<br>**H5-index** | **Mean**<br>**Impact Factor** | **Mean**<br>**CiteScore** | **Mean**<br>**SNIP** | **Mean**<br>**SJR** |
| --- | --- | --- | --- | --- | --- | --- |
| US | 49,203 | 66.809 | 7.665 | 8.043 | 2.048 | 2.138 |
| China | 21,693 | 69.241 | 5.879 | 7.204 | 1.688 | 1.628 |
| Italy | 15,029 | 61.102 | 5.318 | 6.291 | 1.55 | 1.436 |
| UK | 14,475 | 71.561 | 9.842 | 8.64 | 2.318 | 2.059 |
| India | 13,044 | 41.635 | 4.099 | 4.23 | 1.154 | 0.828 |
| Canada | 6,093 | 63.64 | 6.553 | 6.734 | 1.839 | 1.654 |
| Germany | 5,857 | 71.669 | 6.49 | 7.614 | 1.824 | 1.859 |
| France | 5,196 | 70.012 | 6.715 | 8.36 | 1.935 | 1.99 |
| Brazil | 5,024 | 57.759 | 4.423 | 5.235 | 1.415 | 1.148 |
| Australia | 4,719 | 56.525 | 5.485 | 6.118 | 1.658 | 1.487 |
| Turkey | 4,600 | 40.328 | 3.086 | 3.989 | 1.072 | 0.778 |
| Japan | 3,678 | 58.556 | 4.338 | 5.286 | 1.372 | 1.227 |
| Saudi Arabia | 2,792 | 49.524 | 3.814 | 4.24 | 1.334 | 0.87 |
| South Korea | 2,696 | 65.022 | 4.582 | 5.666 | 1.444 | 1.242 |
| Mexico | 1,436 | 61.314 | 4.288 | 5.14 | 1.343 | 1.157 |
| South Africa | 1,339 | 54.233 | 5.761 | 5.528 | 1.567 | 1.324 |
| Russia | 1,323 | 47.833 | 4.905 | 4.928 | 1.26 | 1.103 |
| Indonesia | 1,252 | 37.072 | 3.822 | 3.236 | 1.108 | 0.696 |
| Argentina | 590 | 57.626 | 4.385 | 6.068 | 1.504 | 1.387 |
| EU | 48,140 | 64.922 | 5.618 | 6.598 | 1.639 | 1.549 |

As shown in Figure 19, overall, UK, Germany, France, US and China make the top-5 cumulative mean publication impact of the COVID-19 research. Specifically, as shown in Table 12, US achieves the highest number of publications and the highest mean SJR, UK makes the highest mean Impact Factor, CiteScore and SNIP, Germany makes the highest mean H5-index.

### 6.6 OECD country’s mean publication impact

Similar to the analysis of G20 country’s mean publication impact in Section 6.5, here, we analyze the mean publication impact of OECD countries and regions.

Table 13 shows the publication-averaged mean values of different impact metrics in the 38 OECD countries and regions. Here, the US achieves the highest number of publications, then Iceland makes the highest mean H5-index, impact factor, CiteScore, SNIP and SJR uniformly. Italy, Switzerland, the UK, the UK, the UK and the US make the second in terms of publication number, H5-index, impact factor, CiteScore, SNIP and SJR, respectively.

**Table 13:** OECD country’s publication-averaged mean publication impact

| **Country** | **Num_**<br>**pub** | **Mean**<br>**H5-index** | **Mean**<br>**Impact Factor** | **Mean**<br>**CiteScore** | **Mean**<br>**SNIP** | **Mean**<br>**SJR** |
| --- | --- | --- | --- | --- | --- | --- |
| US | 49,203 | 66.809 | 7.665 | 8.043 | 2.048 | 2.138 |
| Italy | 15,029 | 61.102 | 5.318 | 6.291 | 1.55 | 1.436 |
| UK | 14,475 | 71.561 | 9.842 | 8.64 | 2.318 | 2.059 |
| Spain | 6,372 | 69.03 | 5.264 | 6.121 | 1.574 | 1.4 |
| Canada | 6,093 | 63.64 | 6.553 | 6.734 | 1.839 | 1.654 |
| Germany | 5,857 | 71.669 | 6.49 | 7.614 | 1.824 | 1.859 |
| France | 5,196 | 70.012 | 6.715 | 8.36 | 1.935 | 1.99 |
| Australia | 4,719 | 56.525 | 5.485 | 6.118 | 1.658 | 1.487 |
| Turkey | 4,600 | 40.328 | 3.086 | 3.989 | 1.072 | 0.778 |
| Japan | 3,678 | 58.556 | 4.338 | 5.286 | 1.372 | 1.227 |
| South Korea | 2,696 | 65.022 | 4.582 | 5.666 | 1.444 | 1.242 |
| Netherlands | 2,123 | 72.92 | 7.328 | 8.385 | 2.004 | 2.128 |
| Poland | 2,012 | 65.6 | 4.127 | 4.415 | 1.302 | 1.009 |
| Switzerland | 1,880 | 73.086 | 7.394 | 7.802 | 1.995 | 1.946 |
| Israel | 1,766 | 71.008 | 6.562 | 7.737 | 1.901 | 1.903 |
| Greece | 1,491 | 54.706 | 4.895 | 5.572 | 1.397 | 1.297 |
| Mexico | 1,436 | 61.314 | 4.288 | 5.14 | 1.343 | 1.157 |
| Belgium | 1,424 | 62.561 | 6.036 | 7.291 | 1.787 | 1.731 |
| Portugal | 1,173 | 59.684 | 4.848 | 5.458 | 1.435 | 1.29 |
| Ireland | 1,131 | 50.754 | 5.1 | 5.404 | 1.473 | 1.277 |
| Austria | 1,096 | 65.489 | 5.476 | 6.489 | 1.612 | 1.557 |
| Sweden | 1,067 | 70.201 | 6.725 | 7.525 | 1.908 | 1.937 |
| Denmark | 809 | 71.498 | 6.269 | 7.677 | 1.925 | 1.787 |
| Norway | 639 | 67.307 | 6.092 | 6.479 | 1.738 | 1.574 |
| New Zealand | 599 | 51.561 | 5.93 | 5.414 | 1.622 | 1.237 |
| Colombia | 584 | 55.271 | 4.799 | 5.059 | 1.422 | 1.11 |
| Chile | 574 | 65.697 | 4.883 | 5.587 | 1.593 | 1.31 |
| Finland | 435 | 62.504 | 4.915 | 6.283 | 1.614 | 1.405 |
| Czech Republic | 390 | 67.179 | 4.409 | 4.989 | 1.342 | 1.122 |
| Hungary | 298 | 58.24 | 4.644 | 4.927 | 1.331 | 1.129 |
| Slovenia | 206 | 64.881 | 4.256 | 5.128 | 1.435 | 1.104 |
| Slovakia | 164 | 62.323 | 4.134 | 4.421 | 1.282 | 1.011 |
| Lithuania | 108 | 64.574 | 4.635 | 5.646 | 1.585 | 1.061 |
| Luxembourg | 77 | 66.54 | 4.108 | 5.124 | 1.538 | 1.121 |
| Latvia | 67 | 55.315 | 3.857 | 3.722 | 1.194 | 0.881 |
| Estonia | 51 | 60.865 | 3.753 | 4.639 | 1.278 | 1.076 |
| Costa Rica | 44 | 41.788 | 3.845 | 4.915 | 1.53 | 1.284 |
| Iceland | 26 | 112.4 | 15.504 | 14.732 | 3.262 | 3.526 |

## 7 Publication profile on modeling COVID-19

In this chapter, we analyze the publication profile of those publications on modeling COVID-19. We focus on two perspectives. On one hand, we explore the word cloud of keywords in all modeling publications collected in Section 3.7 with modeling keywords collected in Section 3.5 as well as in three major disciplines and the two mostly published countries the US and China. On the other hand, we extract the major trends of the modeling publications in terms of top-K concerned problems per the COVID-19 problem-related keywords collected in Section 3.4, top-K modeling keywords defined in Section 3.5, top-K published countries, and top-K monthly concerned problems, etc.

### 7.1 Keyword cloud of modeling publications

Here, we present the word cloud of all modeling publications. Modeling publications are those publications extracted in Section 3.7 with modeling keywords defined in Section 3.5. There are 43,921 publications on COVID-19 modeling. We count the frequency of each modeling keyword in these publications, and then extract the top-K keywords to draw the word cloud by the word cloud drawing tool.

Figure 20 shows the word cloud of the top-200 modeling words appeared in all modeling publications. The results show that the modeling methods are diversified, covering classic mathematical and statistical methods, epidemic modeling methods, simulation, and classic machine learning methods.

**Figure 20:**
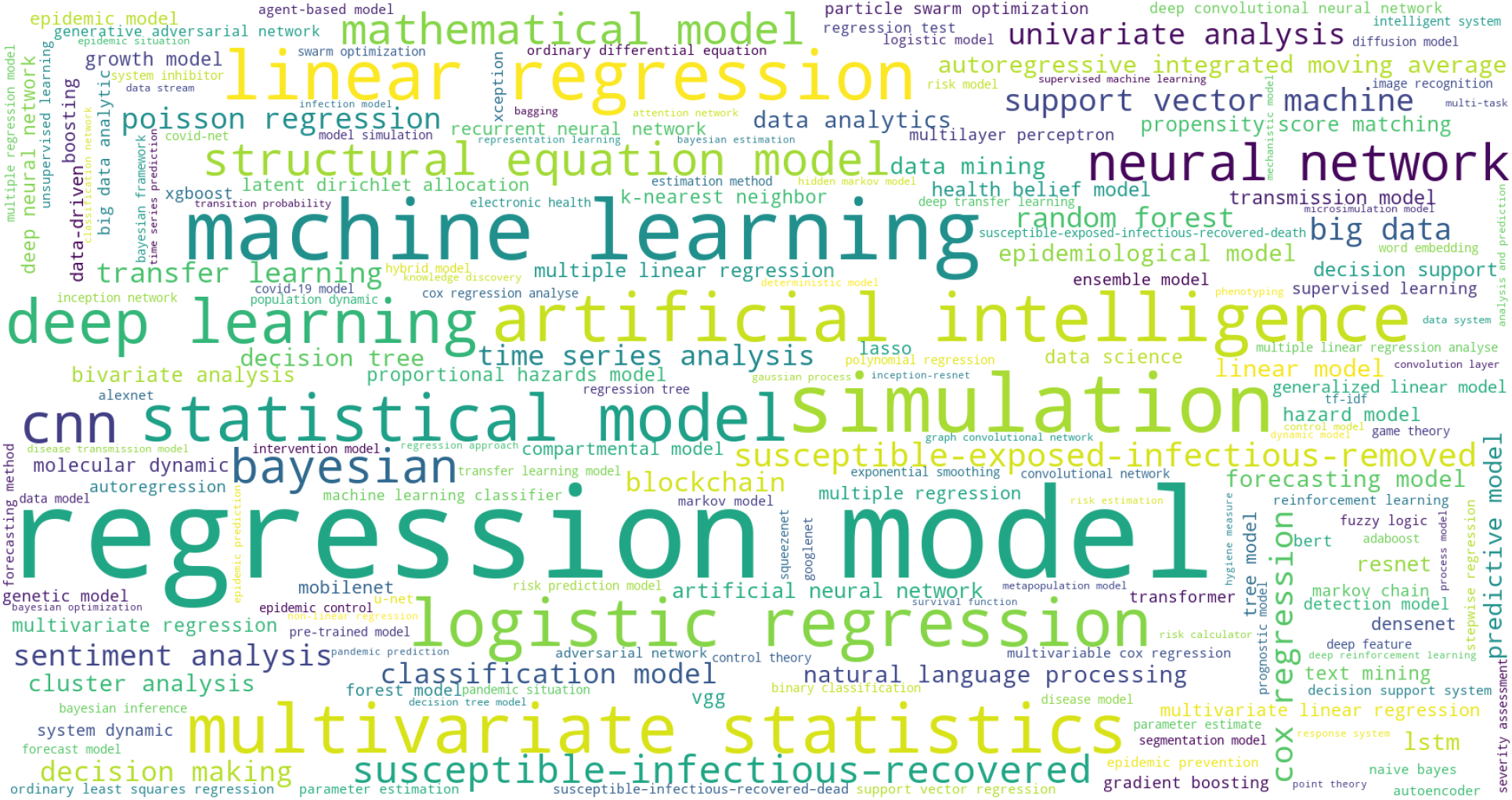
Modeling keyword cloud in modeling publications

Further, Table 14 extracts the top-10 modeling keywords mostly appearing in the modeling publications: regression model, machine learning, simulation, linear regression, multivariate statistics, artificial intelligence, logistic regression, statistical model, deep learning, and CNN. They show that classic regression methods dominate the publications in modeling COVID-19, statistical models and machine learning follow that. Deep learning, although emerged only in a rather shorter period than those classic methods, makes a strong presence in modeling COVID-19, in particular, by the most fundamental network CNN.

**Table 14:** Top-10 modeling keywords in modeling publications

| **Modeling keywords** | **Document frequency** |
| --- | --- |
| regression model | 9,048 |
| machine learning | 3,054 |
| simulation | 2,881 |
| linear regression | 1,942 |
| multivariate statistics | 1,918 |
| artificial intelligence | 1,844 |
| logistic regression | 1,799 |
| statistical model | 1,719 |
| deep learning | 1,527 |
| CNN | 1,205 |

The source code for generating the word cloud can be found in the file:

data_processing/word_cloud.py. The data can be found in “Modeling keyword cloud in modeling publications.xlsx”

### 7.2 Keyword cloud of modeling publications in computer science

Here, we further analyze the word cloud of the keywords in computer science publications on COVID-19. We take the similar method to that in Section 7.1.

Figure 21 shows the word cloud of the top-200 modeling words appeared in modeling publications which belong to the computer science category, categorized per the method discussed in Section 3.6 on the disciplinary categorization, with the list of disciplines presented in Appendix 10.1.

**Figure 21:**
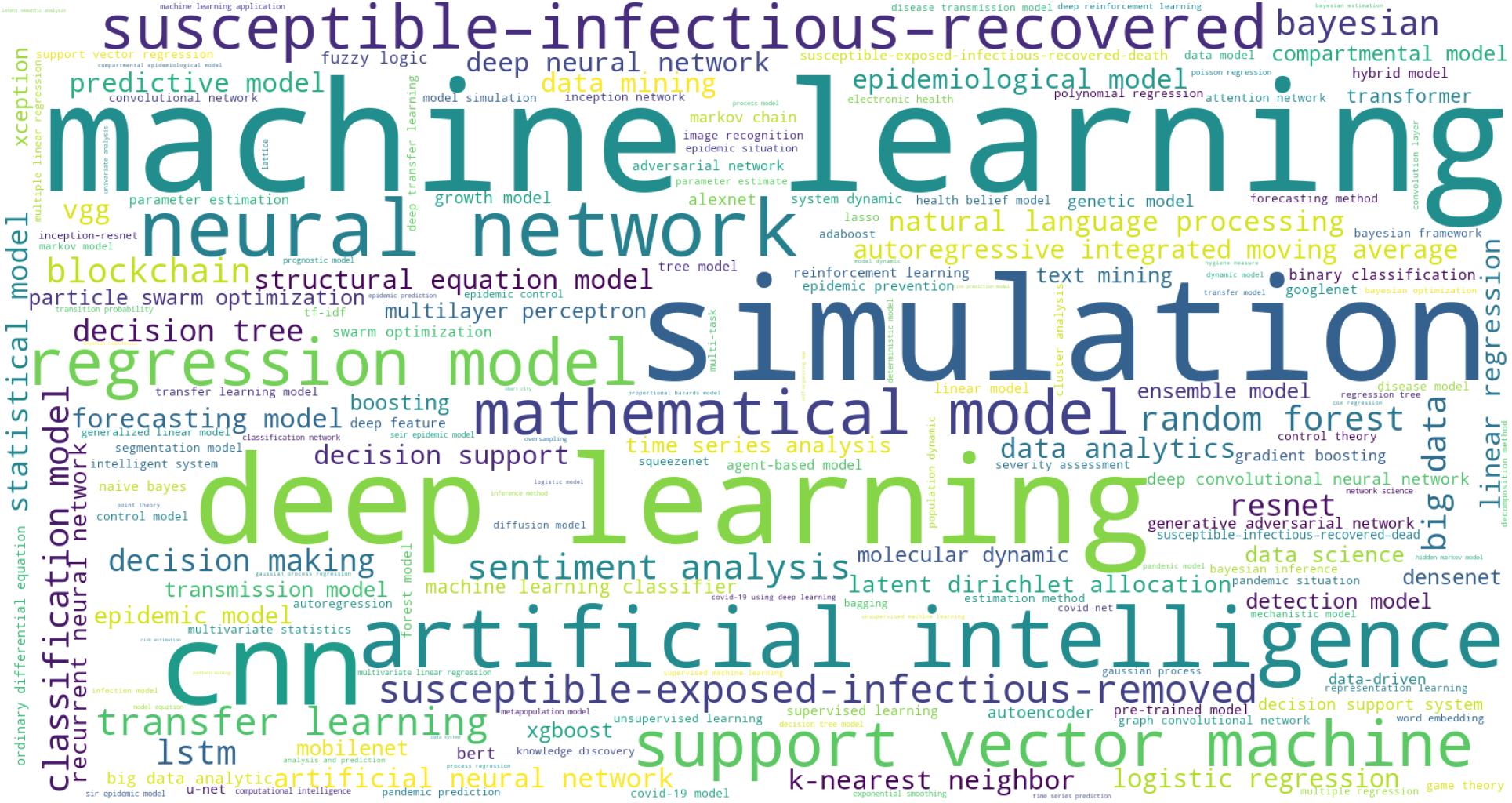
Modeling keyword cloud of modeling publications in computer science

The results show that, in contrast to that of all modeling publications in Figure 20, the modeling publications from the computer science discipline mainly applied machine learning, deep learning and AI methods, although mathematical models also appeared highly frequently in the publications. Contrast to the top-10 keywords of all modeling publications in Table 14, here, as shown in Table 15, classic machine learning methods including neural networks and SVM, simulation, and deep learning networks including CNN dominate the publications, although their frequencies are much lower than that in the overall modeling publications.

**Table 15:** Top-10 keywords in modeling publications in computer science

| Keyword | Document frequency |
| --- | --- |
| machine learning | 883 |
| simulation | 773 |
| deep learning | 559 |
| CNN | 489 |
| artificial intelligence | 452 |
| neural network | 396 |
| regression model | 254 |
| support vector machine | 244 |
| mathematical model | 230 |
| susceptible–infectious–recovered | 225 |

The source code for generating the word cloud can be found in the file:

data_processing/word_cloud.py. The data can be found in “Modeling word cloud of modeling publications in computer science.xlsx.”

### 7.3 Keyword cloud of modeling publications in medical science

Here, we take the similar method as in Section 7.1 to further analyze the word cloud of the keywords in medical science publications on COVID-19.

Figure 22 shows the word cloud of the top-200 modeling keywords appeared in modeling publications which belong to the medical science category. These publications are categorized into medical science per the method introduced in Section 3.6 and the list of disciplinary categorization in Appendix 10.1. Table 16 further lists the top-10 keywords in medical science publications.

**Table 16:** Top-10 keywords in modeling publications in medical science

| **Keyword** | **Count** |
| --- | --- |
| regression model | 5,921 |
| multivariate statistics | 1,461 |
| logistic regression | 1,235 |
| simulation | 1,173 |
| linear regression | 1,158 |
| statistical model | 1,006 |
| machine learning | 977 |
| artificial intelligence | 652 |
| cox regression | 519 |
| Bayesian | 476 |

**Figure 22:**
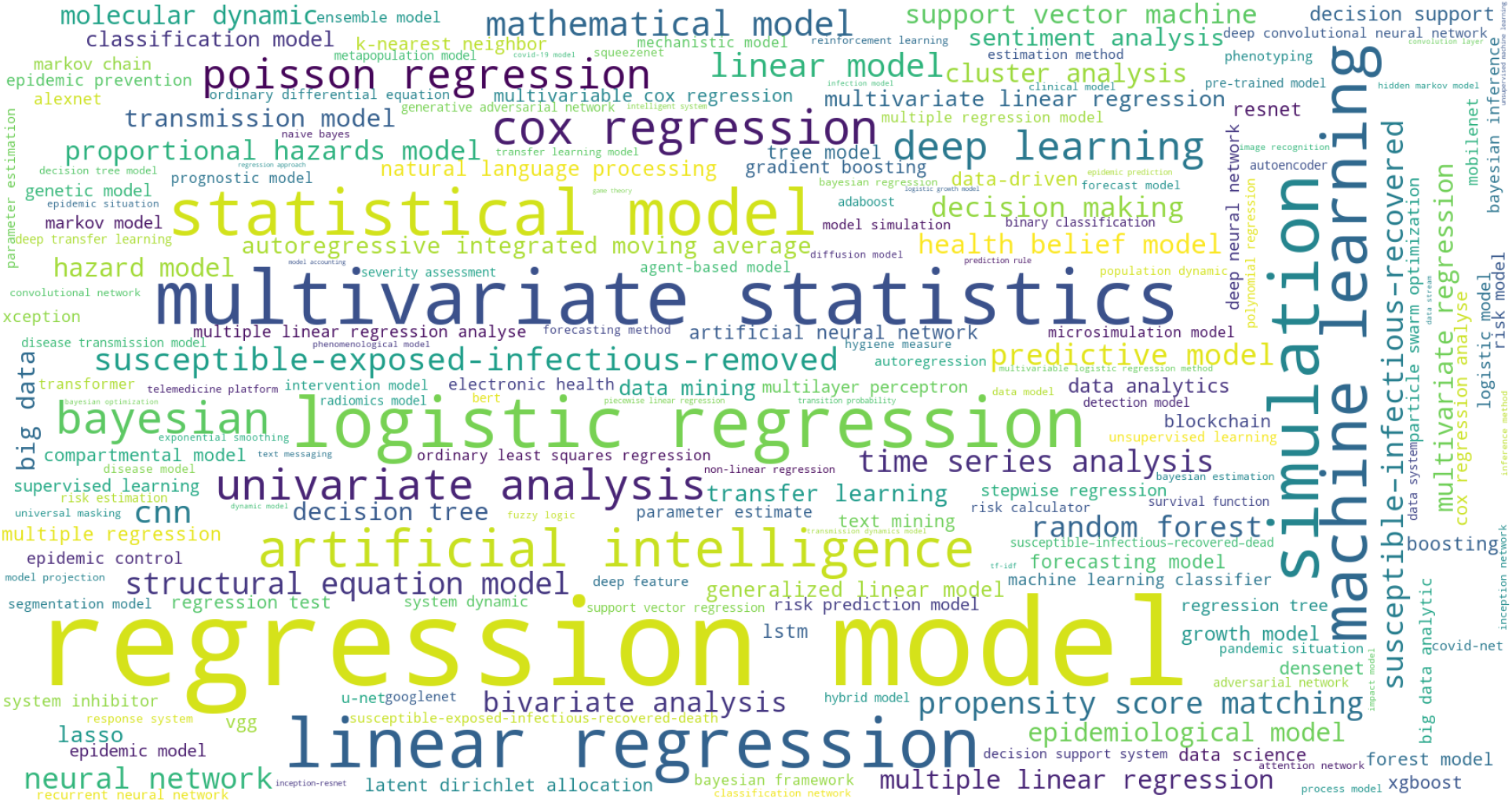
Modeling word cloud of modeling publications in medical science

In contrast to the results presented in Figure 20 and Table 14 for all modeling publications and Figure 21 and Table 15 for modeling publications in computer science, the top-200 keywords shown in Figure 22 and the top-10 keywords in Table 16 largely overlap those in the overall and computer science-based publications. However, publications related to medical science seem to favor classic mathematical and statistical methods than those modern methods such as deep learning and machine learning methods, and their frequencies are also much higher than those in computer science.

The source code for generating the word cloud can be found in the file:

data_processing/word_cloud.py. The data can be found in “Modeling word cloud of modeling publications in medical science.xlsx”.

### 7.4 Keyword cloud of modeling publications in social science

Similarly, here we further analyze the word cloud of the keywords in social science publications on COVID-19.

Figure 23 shows the keyword cloud of the top-200 modeling words appeared in modeling publications which were published in SSRN, as collected in the method introduced in Section 3.6 and the list of disciplinary categorization in Appendix 10.1. Table 17 further lists their top-10 keywords.

**Figure 23:**
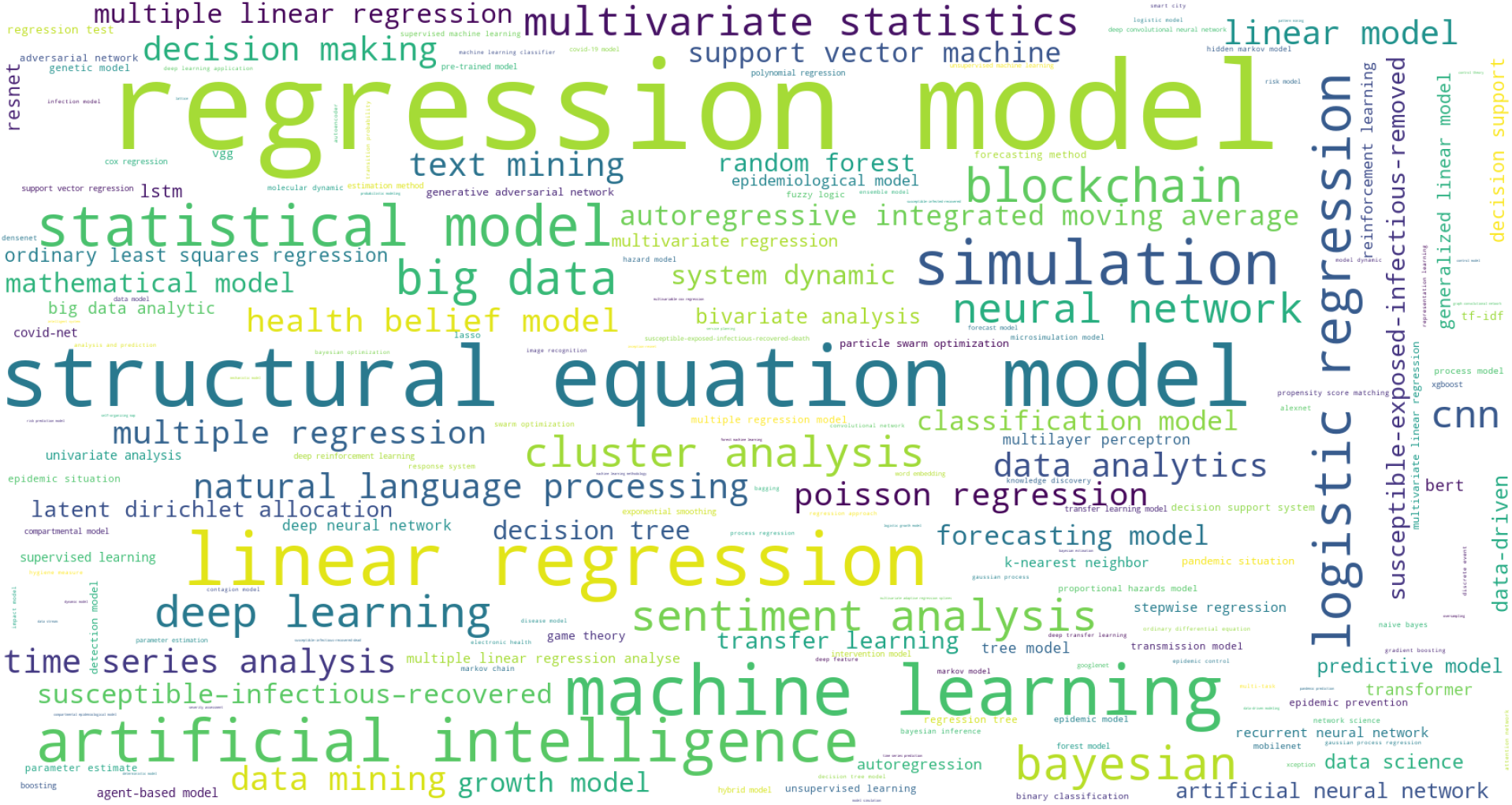
Keyword cloud of modeling publications in social science

**Table 17:** Top-10 keywords of modeling publications in social science

| Keyword | Count |
| --- | --- |
| regression model | 812 |
| structural equation model | 299 |
| linear regression | 219 |
| machine learning | 197 |
| simulation | 151 |
| artificial intelligence | 142 |
| statistical model | 129 |
| logistic regression | 104 |
| big data | 84 |
| blockchain | 84 |

In comparison to the results presented in Figure 20 and Table 14 for all modeling publications, Figure 21 and Table 15 for modeling publications in computer science, Figure 22 and Table 16 for medical science, here, social science publications on COVID-19 favor classic mathematical tools such as regression models and structural equation models, although machine learning and AI methods also play a significant role. It is interesting to see big data and blockchain emerge as top concerns in social science publications.

The source code for generating the word cloud can be found in the file:

data_processing/word_cloud.py. The data can be found in “Modeling word cloud of modeling publications in in social science.xlsx.”

### 7.5 Keyword cloud of modeling publications from the US

Since the US and China are the top-2 countries with most of the publications on COVID-19, we analyze their mostly favored modeling methods in this section and Section 7.6. We extracted those modeling publications with the first authors from the US for this analysis.

Figure 24 shows the word cloud of the top-200 modeling keywords appeared in the modeling publications whose first authors are from the US. Table 18 lists their top-10 modeling keywords.

**Table 18:**
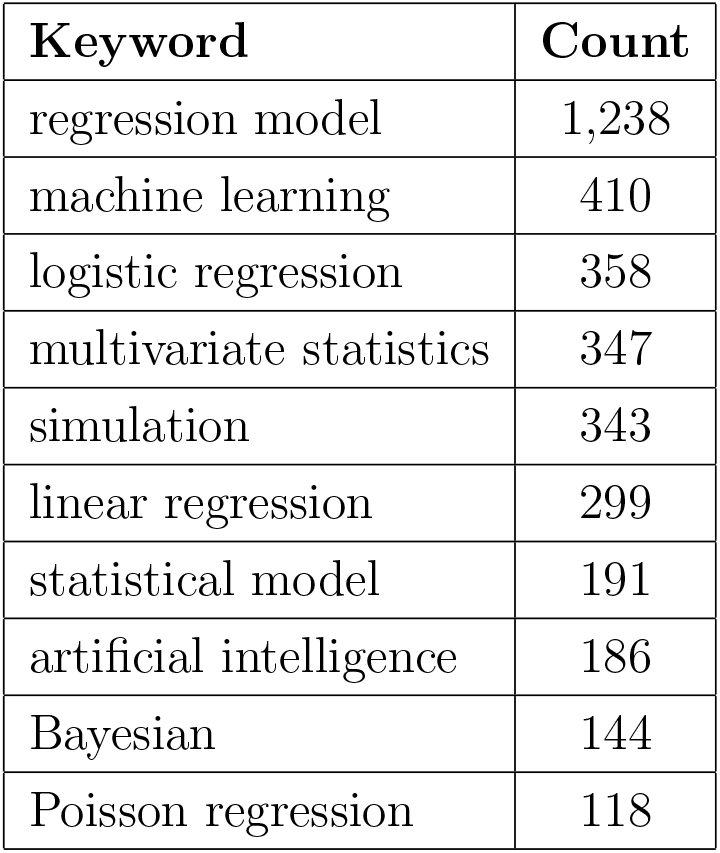
Top-10 keywords in modeling publications from US

| **Keyword** | **Count** |
| --- | --- |
| regression model | 1,238 |
| machine learning | 410 |
| logistic regression | 358 |
| multivariate statistics | 347 |
| simulation | 343 |
| linear regression | 299 |
| statistical model | 191 |
| artificial intelligence | 186 |
| Bayesian | 144 |
| Poisson regression | 118 |

**Figure 24:**
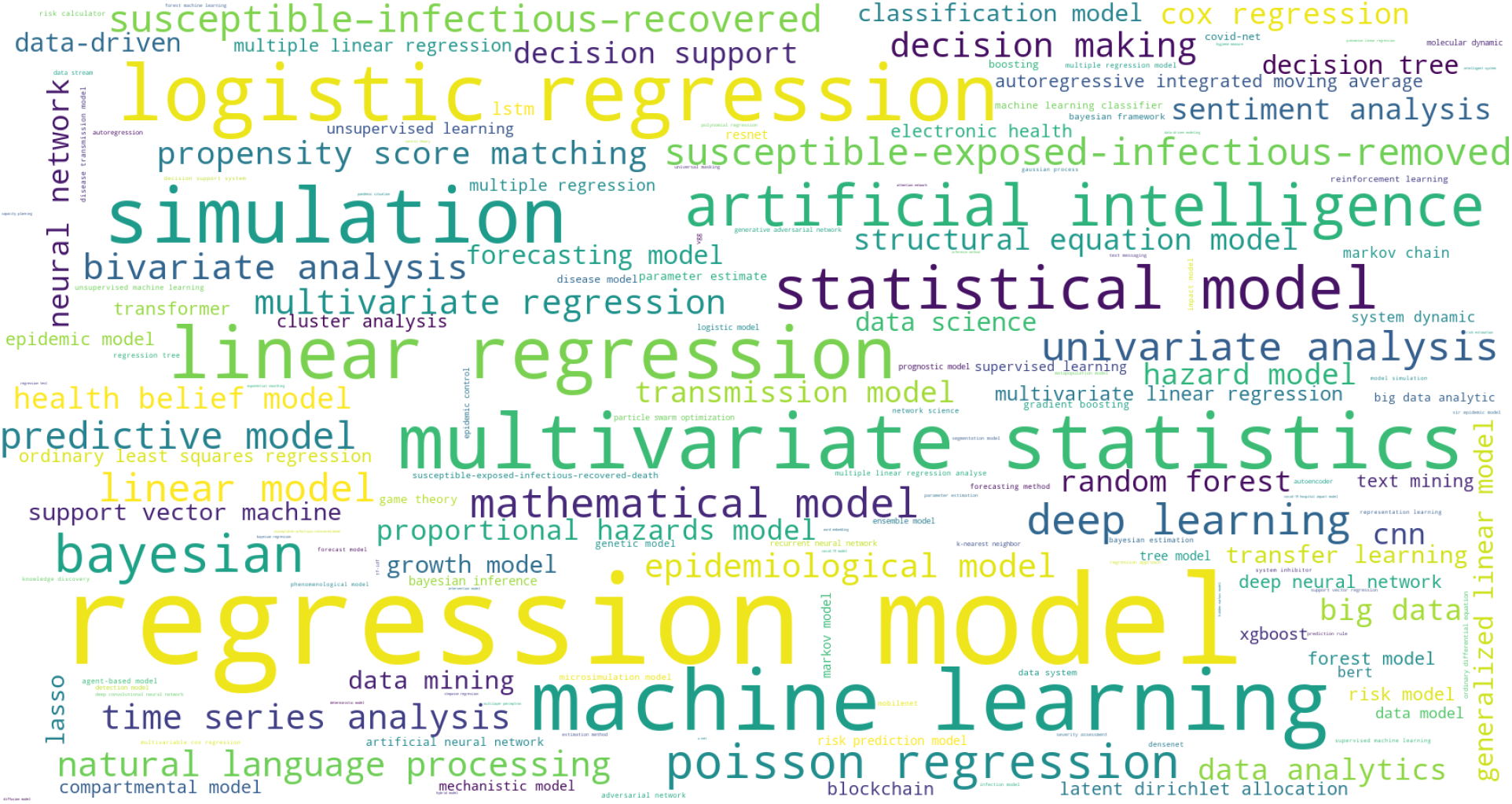
Keyword cloud of modeling publications from the US

In comparison to the results presented in Figure 20 and Table 14 for all modeling publications, publications from the US seem to overwhelmingly favor classic modeling methods from statistical and mathematical families, including regression models, multivariate and Bayesian methods. It is realized that deep learning and CNN, favored in the global modeling communities, did not mark their top-10 roles here in the US publications.

The collection and categorization of first-author countries are introduced in Section 3.2. The source code for generating the word cloud can be found in the data_processing/word_cloud.py. The data can be found in “Modeling word cloud of modeling publications from the US.xlsx.”

### 7.6 Keyword cloud of modeling publications from China

Similar to the analysis in Section 7.5 on the keywords of modeling publications in the US, here, we analyze those mostly applied by the first authors from China.

Figure 25 shows the word cloud of the top-200 modeling keywords appeared in modeling publications whose first authors are from China mainland. Table 19 shows the top-10 keywords mostly concerned by Chinese researchers.

**Figure 25:**
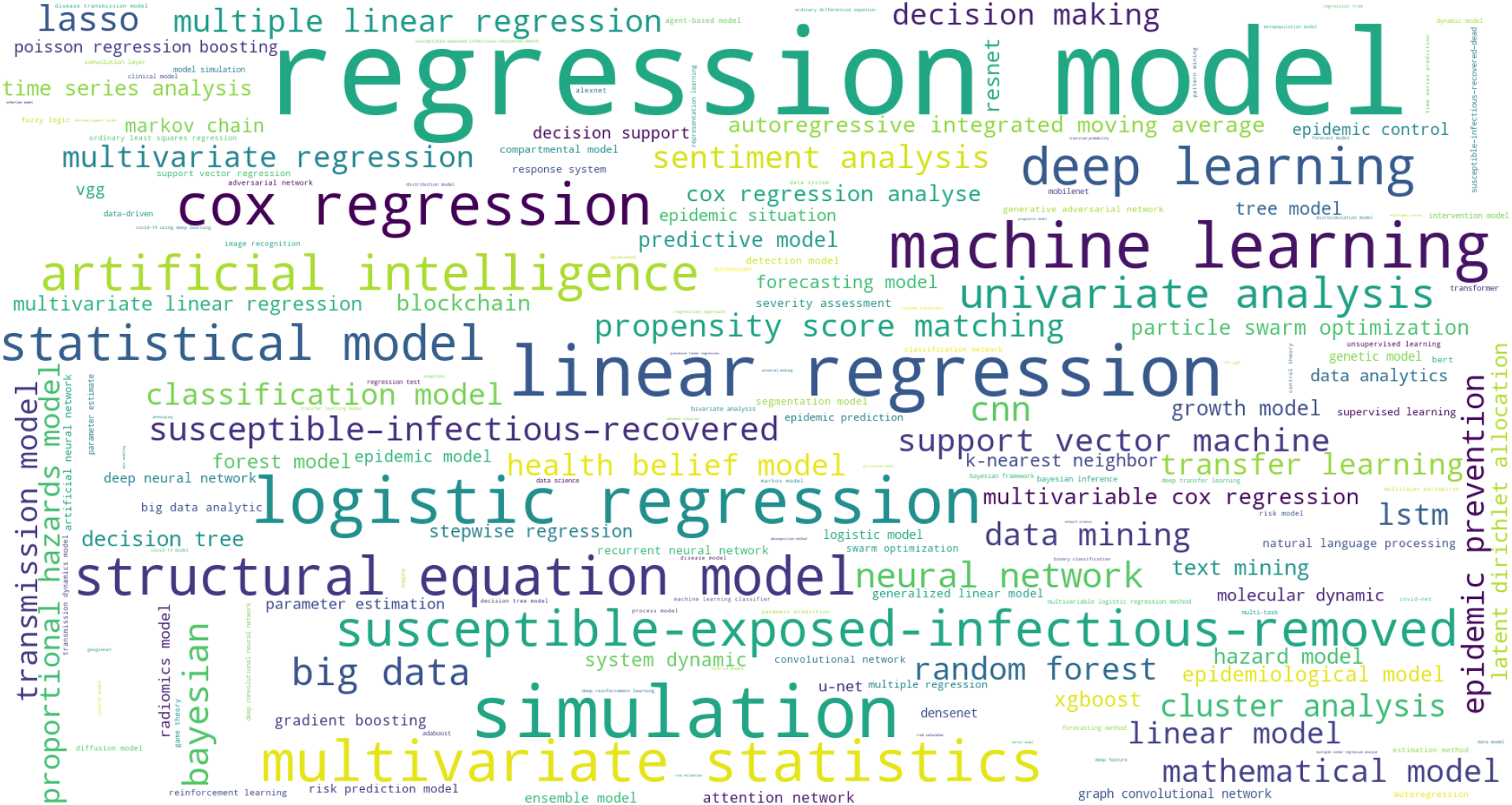
Keyword cloud of modeling publications from China

**Table 19:** Top-10 keywords in modeling publications from China

| Keyword | Count |
| --- | --- |
| regression model | 1,191 |
| simulation | 249 |
| linear regression | 236 |
| logistic regression | 194 |
| machine learning | 188 |
| cox regression | 155 |
| multivariate statistics | 152 |
| structural equation model | 134 |
| deep learning | 123 |
| susceptible-exposed-infectious-removed | 116 |

In comparison to the results presented in Figure 24 and Table 18 for the US, the results in Figure 25 and Table 19 show major similarity in favoring classic regression and statistical models but also significant priority difference, where Chinese researchers favored structural equation models, deep learning, and epidemic models like the susceptible-exposed-infectious-removed (SEIR) model. It is interesting to see, although both countries appreciated the importance of machine learning and simulation, their priorities (i.e., frequencies appearing in respective modeling publications) are exactly opposite, Chinese researchers like simulation, while the American ones prefer machine learning as their no. 2 favorite tool. It is also interesting to see Chinese drops ‘artificial intelligence’ and ‘Bayesian’ in their publications in the top-10 list, which were favored by Americans instead.

The collection and categorization of author countries are introduced in Section 3.2. The source code for generating the word cloud can be found in the data_processing/word_cloud.py. The data can be found in “Modeling word cloud of modeling publications from China.xlsx.”

### 7.7 Keyword cloud of modeling publications from EU

Similar to the analysis in Section 7.5 on the keywords of modeling publications in the US, here, we analyze those mostly applied by the first authors from the EU.

Figure 26 shows the word cloud of the top-200 modeling keywords appeared in modeling publications whose first authors are from China mainland. Table 20 lists their top-10 modeling keywords.

**Figure 26:**
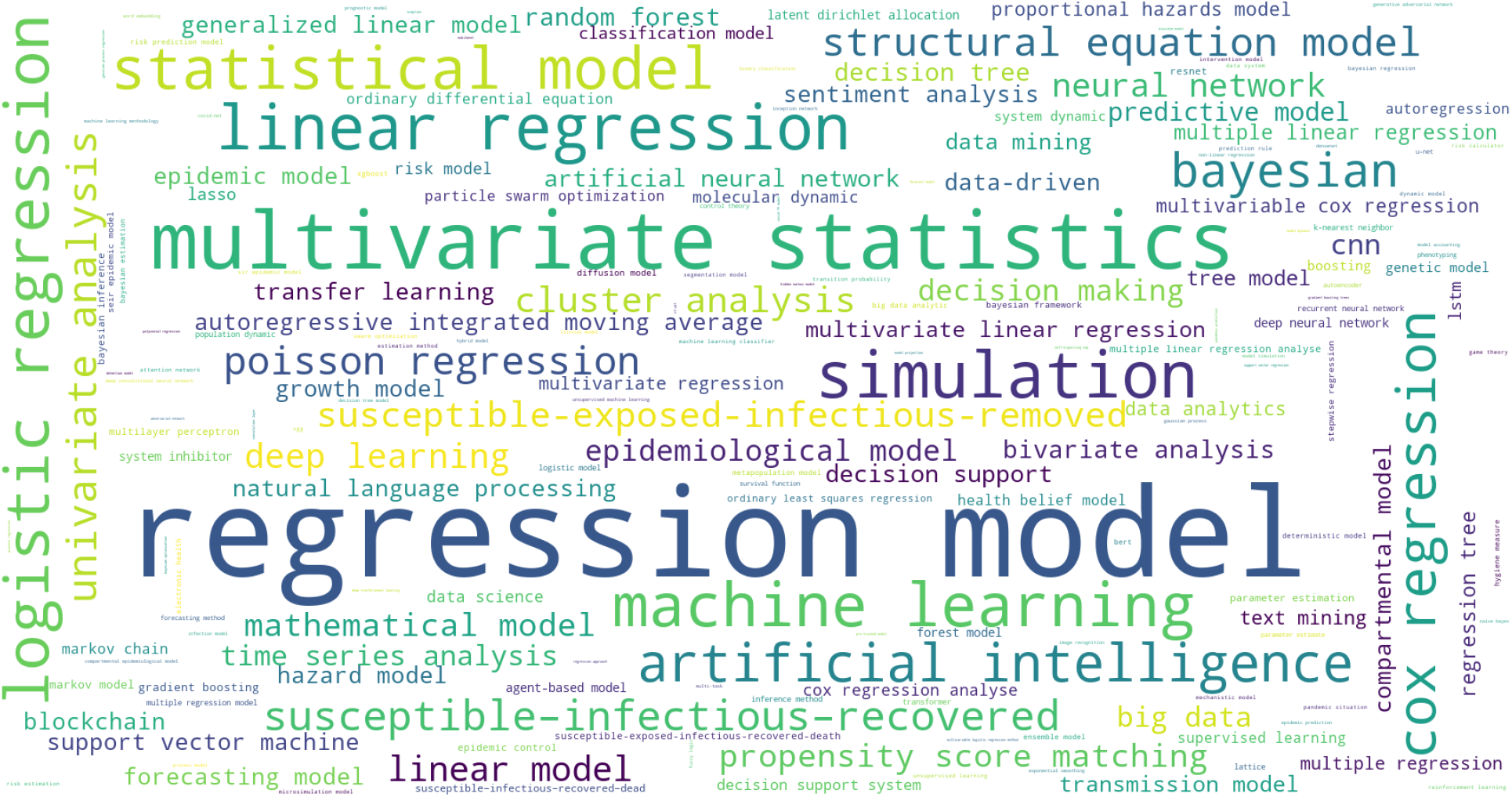
Keyword cloud of modeling publications from the EU

**Table 20:** Top-10 keywords in modeling publications from the EU

| Keyword | Count |
| --- | --- |
| regression model | 1503 |
| multivariate statistics | 452 |
| simulation | 279 |
| linear regression | 264 |
| logistic regression | 252 |
| machine learning | 251 |
| statistical model | 239 |
| cox regression | 192 |
| artificial intelligence | 188 |
| bayesian | 149 |

### 7.8 Top-50 modeling problems and their publication impact

Here, we analyze the mostly concerned COVID-19 problems in modeling publications and their publication impact. The problem-related keywords are selected per the method introduced in Section 3.4 from three resources. We then select the top-50 problem-related keywords, and extract their associated modeling publications. We further calculate their mean publication impact metrics in terms of H5-index, Impact Factor, CiteScore, SNIP, and SJR. The mean publication impact is calculated per the method introduced in Section 4.1.

Table 21 shows the mean of publication research impact in terms of metrics H5-index, Impact Factor, CiteScore, SNIP and SJR for the publications associated with the top-50 problem keywords.

**Table 21:**
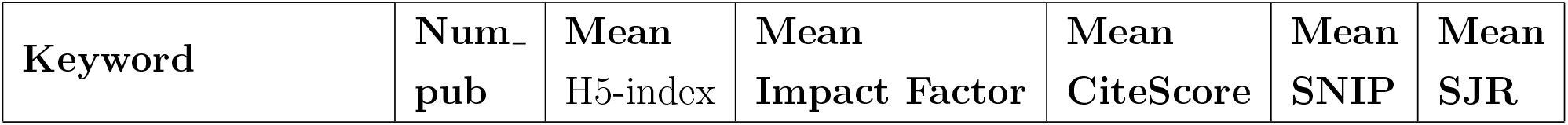
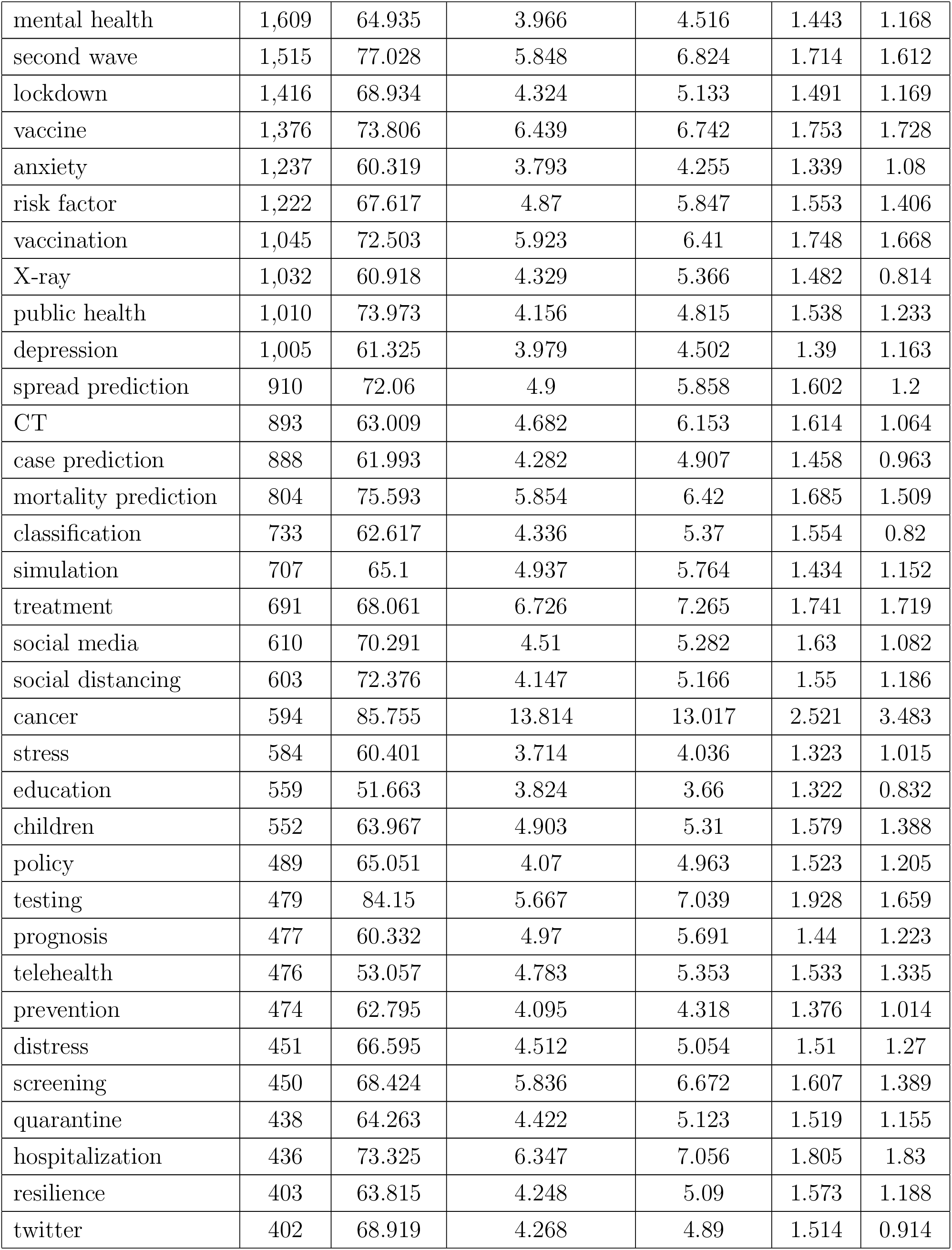
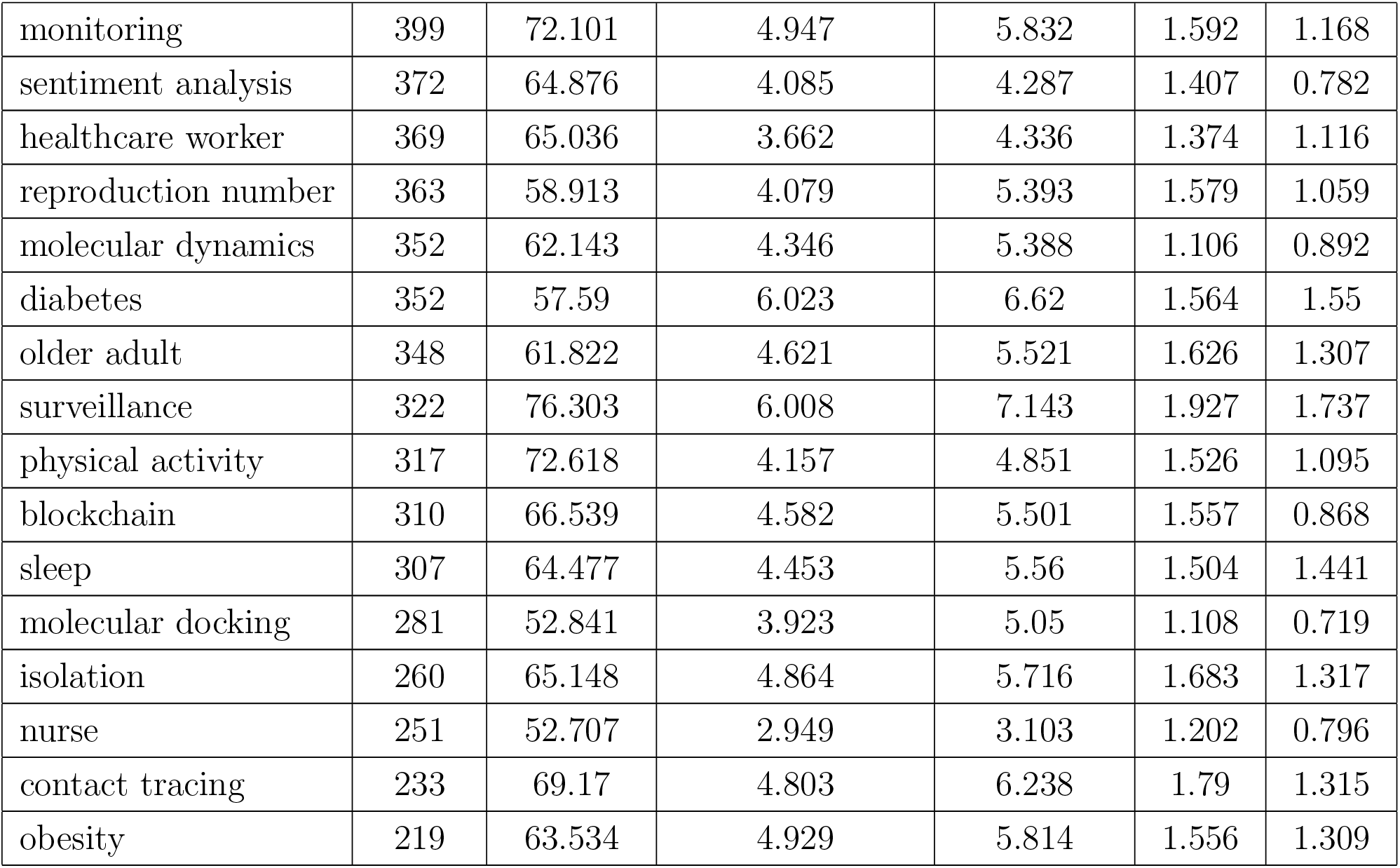
Mean publication impact of top-50 modeling problems

| Keyword | Num.<br>pub | Mean<br>H5-index | Mean<br>Impact Factor | Mean<br>CiteScore | Mean<br>SNIP | Mean<br>SJR |
| --- | --- | --- | --- | --- | --- | --- |
| mental health | 1,609 | 64.935 | 3.966 | 4.516 | 1.443 | 1.168 |
| second wave | 1,515 | 77.028 | 5.848 | 6.824 | 1.714 | 1.612 |
| lockdown | 1,416 | 68.934 | 4.324 | 5.133 | 1.491 | 1.169 |
| vaccine | 1,376 | 73.806 | 6.439 | 6.742 | 1.753 | 1.728 |
| anxiety | 1,237 | 60.319 | 3.793 | 4.255 | 1.339 | 1.08 |
| risk factor | 1,222 | 67.617 | 4.87 | 5.847 | 1.553 | 1.406 |
| vaccination | 1,045 | 72.503 | 5.923 | 6.41 | 1.748 | 1.668 |
| X-ray | 1,032 | 60.918 | 4.329 | 5.366 | 1.482 | 0.814 |
| public health | 1,010 | 73.973 | 4.156 | 4.815 | 1.538 | 1.233 |
| depression | 1,005 | 61.325 | 3.979 | 4.502 | 1.39 | 1.163 |
| spread prediction | 910 | 72.06 | 4.9 | 5.858 | 1.602 | 1.2 |
| CT | 893 | 63.009 | 4.682 | 6.153 | 1.614 | 1.064 |
| case prediction | 888 | 61.993 | 4.282 | 4.907 | 1.458 | 0.963 |
| mortality prediction | 804 | 75.593 | 5.854 | 6.42 | 1.685 | 1.509 |
| classification | 733 | 62.617 | 4.336 | 5.37 | 1.554 | 0.82 |
| simulation | 707 | 65.1 | 4.937 | 5.764 | 1.434 | 1.152 |
| treatment | 691 | 68.061 | 6.726 | 7.265 | 1.741 | 1.719 |
| social media | 610 | 70.291 | 4.51 | 5.282 | 1.63 | 1.082 |
| social distancing | 603 | 72.376 | 4.147 | 5.166 | 1.55 | 1.186 |
| cancer | 594 | 85.755 | 13.814 | 13.017 | 2.521 | 3.483 |
| stress | 584 | 60.401 | 3.714 | 4.036 | 1.323 | 1.015 |
| education | 559 | 51.663 | 3.824 | 3.66 | 1.322 | 0.832 |
| children | 552 | 63.967 | 4.903 | 5.31 | 1.579 | 1.388 |
| policy | 489 | 65.051 | 4.07 | 4.963 | 1.523 | 1.205 |
| testing | 479 | 84.15 | 5.667 | 7.039 | 1.928 | 1.659 |
| prognosis | 477 | 60.332 | 4.97 | 5.691 | 1.44 | 1.223 |
| telehealth | 476 | 53.057 | 4.783 | 5.353 | 1.533 | 1.335 |
| prevention | 474 | 62.795 | 4.095 | 4.318 | 1.376 | 1.014 |
| distress | 451 | 66.595 | 4.512 | 5.054 | 1.51 | 1.27 |
| screening | 450 | 68.424 | 5.836 | 6.672 | 1.607 | 1.389 |
| quarantine | 438 | 64.263 | 4.422 | 5.123 | 1.519 | 1.155 |
| hospitalization | 436 | 73.325 | 6.347 | 7.056 | 1.805 | 1.83 |
| resilience | 403 | 63.815 | 4.248 | 5.09 | 1.573 | 1.188 |
| twitter | 402 | 68.919 | 4.268 | 4.89 | 1.514 | 0.914 |
| monitoring | 399 | 72.101 | 4.947 | 5.832 | 1.592 | 1.168 |
| sentiment analysis | 372 | 64.876 | 4.085 | 4.287 | 1.407 | 0.782 |
| healthcare worker | 369 | 65.036 | 3.662 | 4.336 | 1.374 | 1.116 |
| reproduction number | 363 | 58.913 | 4.079 | 5.393 | 1.579 | 1.059 |
| molecular dynamics | 352 | 62.143 | 4.346 | 5.388 | 1.106 | 0.892 |
| diabetes | 352 | 57.59 | 6.023 | 6.62 | 1.564 | 1.55 |
| older adult | 348 | 61.822 | 4.621 | 5.521 | 1.626 | 1.307 |
| surveillance | 322 | 76.303 | 6.008 | 7.143 | 1.927 | 1.737 |
| physical activity | 317 | 72.618 | 4.157 | 4.851 | 1.526 | 1.095 |
| blockchain | 310 | 66.539 | 4.582 | 5.501 | 1.557 | 0.868 |
| sleep | 307 | 64.477 | 4.453 | 5.56 | 1.504 | 1.441 |
| molecular docking | 281 | 52.841 | 3.923 | 5.05 | 1.108 | 0.719 |
| isolation | 260 | 65.148 | 4.864 | 5.716 | 1.683 | 1.317 |
| nurse | 251 | 52.707 | 2.949 | 3.103 | 1.202 | 0.796 |
| contact tracing | 233 | 69.17 | 4.803 | 6.238 | 1.79 | 1.315 |
| obesity | 219 | 63.534 | 4.929 | 5.814 | 1.556 | 1.309 |

The top-10 concerned problem keywords in the literature are mental health, second wave, lockdown, vaccine, anxiety, risk factor, vaccination, X-ray, public health, and depression. Further, of the top-50 problem keywords, the keywords can be grouped into the following major aspects and categories:

- various issues related to mental health are mostly concerned, including anxiety, depression, stress, distress, sentiment analysis, and sleep, forming the mostly concern issues in the literature (5193).
- non-pharmaceutical interventions and policies are widely concerned, including lockdown, social distancing, policy, quarantine, surveillance, physical activity, and isolation.
- issues related to public health, vaccine, vaccination, testing, prevention, telehealth, screening, monitoring, and contact tracing, and concerns about children, cancer, diabetes, older adult, sleep, and obesity are widely concerned in the literature.
- many literature concerns about risk factor, X-ray, CT, classification, treatment, hospitalization, and prognosis.
- many references are on the prediction of spread, cases, mortality, and reproduction number.

### 7.9 Top-50 problems and their modeling methods

Here, we analyze the popularly concerned problems in COVID-19 modeling and their modeling methods. We extract the problem keywords per the method introduced in Section 3.4. Those modeling publications associated with the top-50 selected problem keywords are extracted per the method introduced in Section 3.7. The modeling keywords are then extracted from these problems-related publications by the method introduced in Section 3.5.

Figure 27 shows the mapping relationship between the top-50 problem keywords and their modeling methods. Each cell in the diagram refers to the interaction between a COVID-19 problem and its related modeling methods. The vertical columns on the top and at the bottom respectively refer to the categories of problems and specific problems of each category. The top-50 specific problems are grouped into seven major categories: epidemic, detection, impact, mental health, NIP, vaccine, and other. The horizontal rows correspond to the families of modeling methods and their specific methods. The families of modeling methods are machine learning, deep learning, statistical modeling, epidemic modeling, and others, labelled on the left of the diagram. Their specific methods are labelled on the left of the diagram respectively.

**Figure 27:**
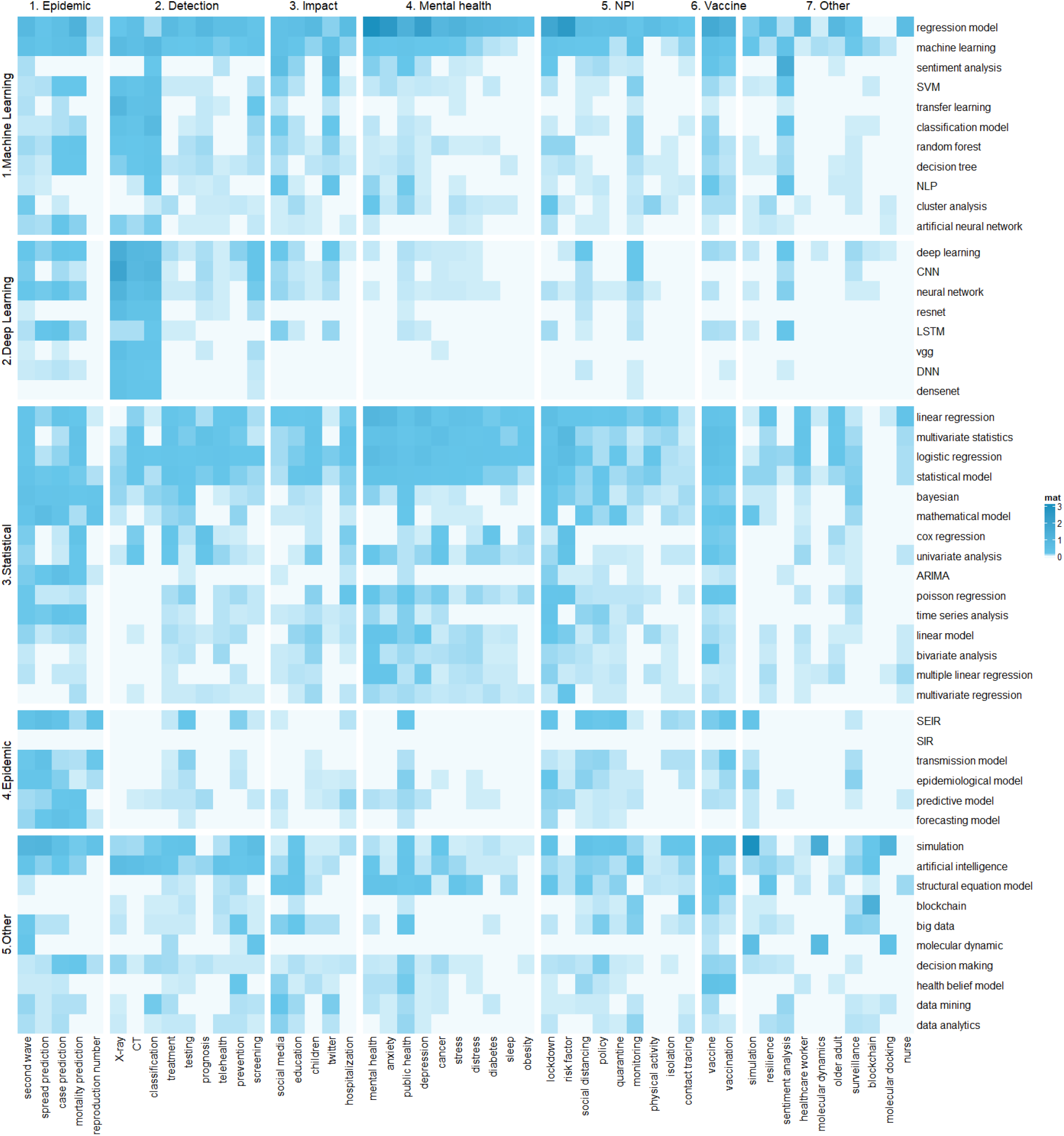
Mapping between top-50 problems and their modeling methods

The color of each cell measures the percentage is equal to the publications of the business problems and their modeling methods/top-50 business problems-related publications. The color of cell stands for the percentage, dark-to-light colors refer to high-to-low percentages correspondingly.

The data can be found in “Mapping between top-50 business problems and their modelling methods.xlsx.”

### 7.10 Top-50 modeling methods and their publication impact

Here, we analyze the popular modeling methods applied in modeling COVID-19 and then analyze their publication impact. We extract the modeling keywords per the method introduced in Section 3.5. Then, the publications association with top-K modeling keywords are selected. We further calculate the publication impact of these publications.

Table 23 shows the top-50 modeling keywords concerned in modeling publications on COVID-19 and their mean publication impact metrics in terms of H5-index, Impact Factor, CiteScore, SNIP, and SJR. The mean publication impact is calculated per the method introduced in Section 4.1.

**Table 22:**
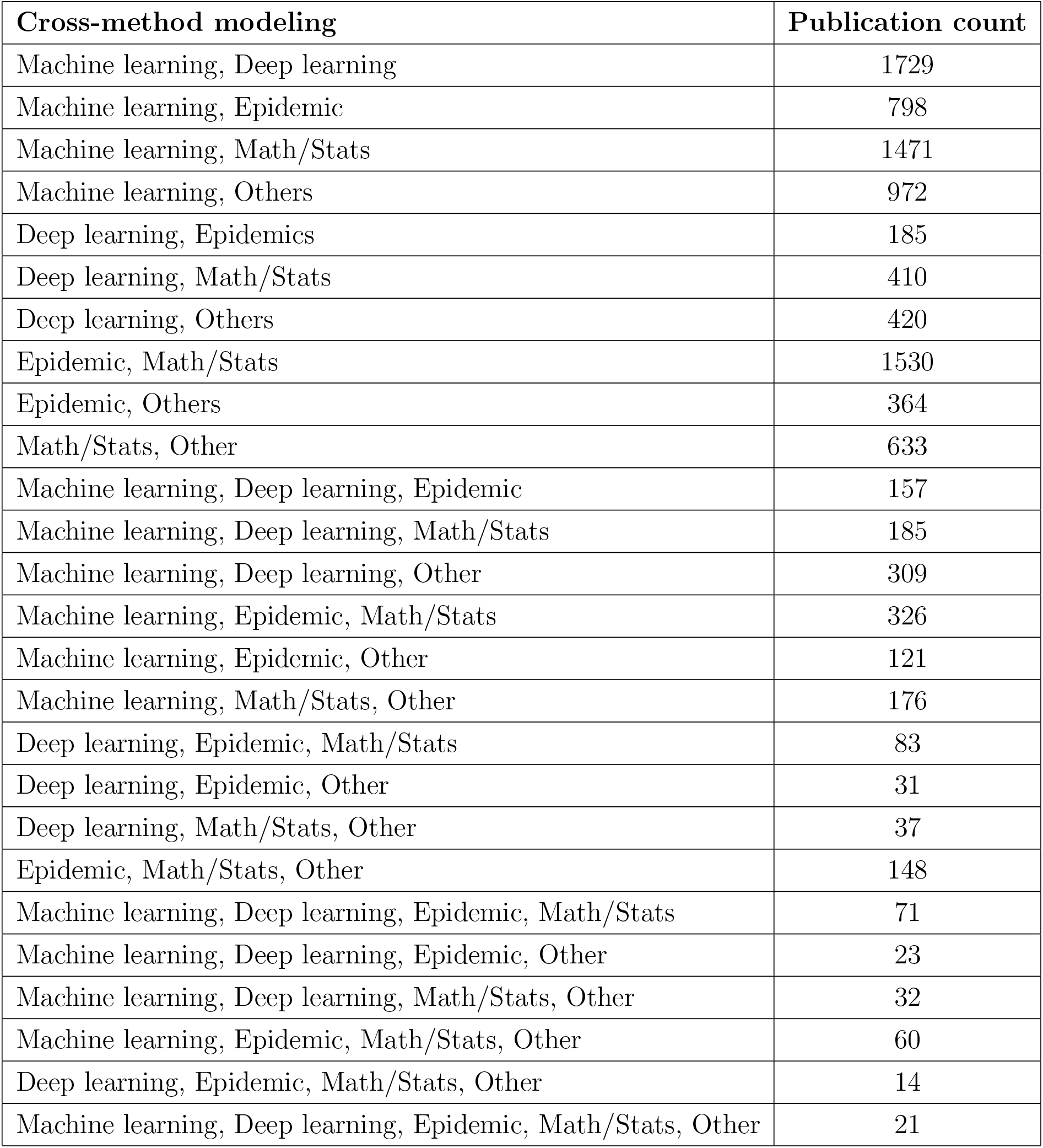
Cross-disciplinary/method modeling of COVID-19

**Table 23:**
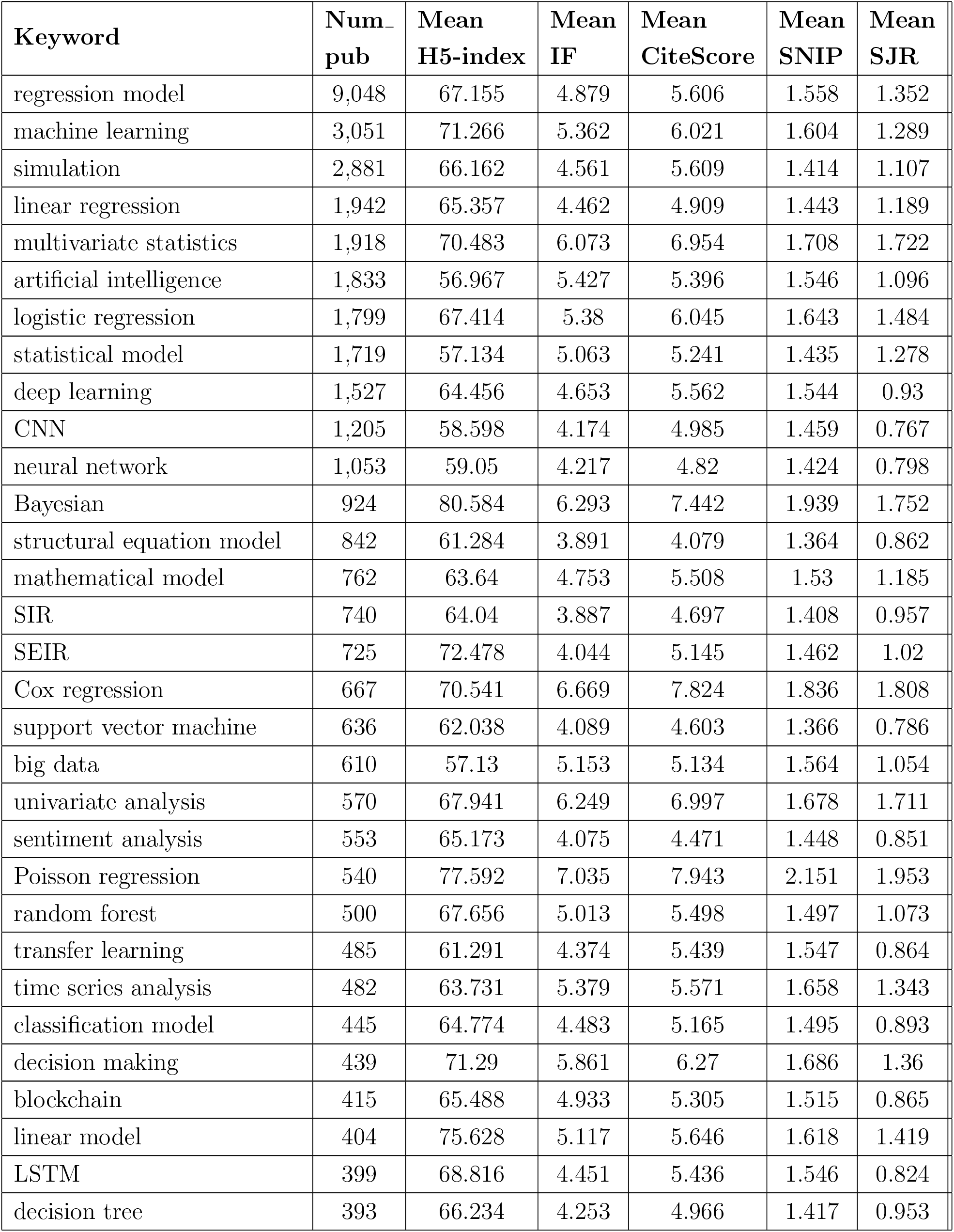
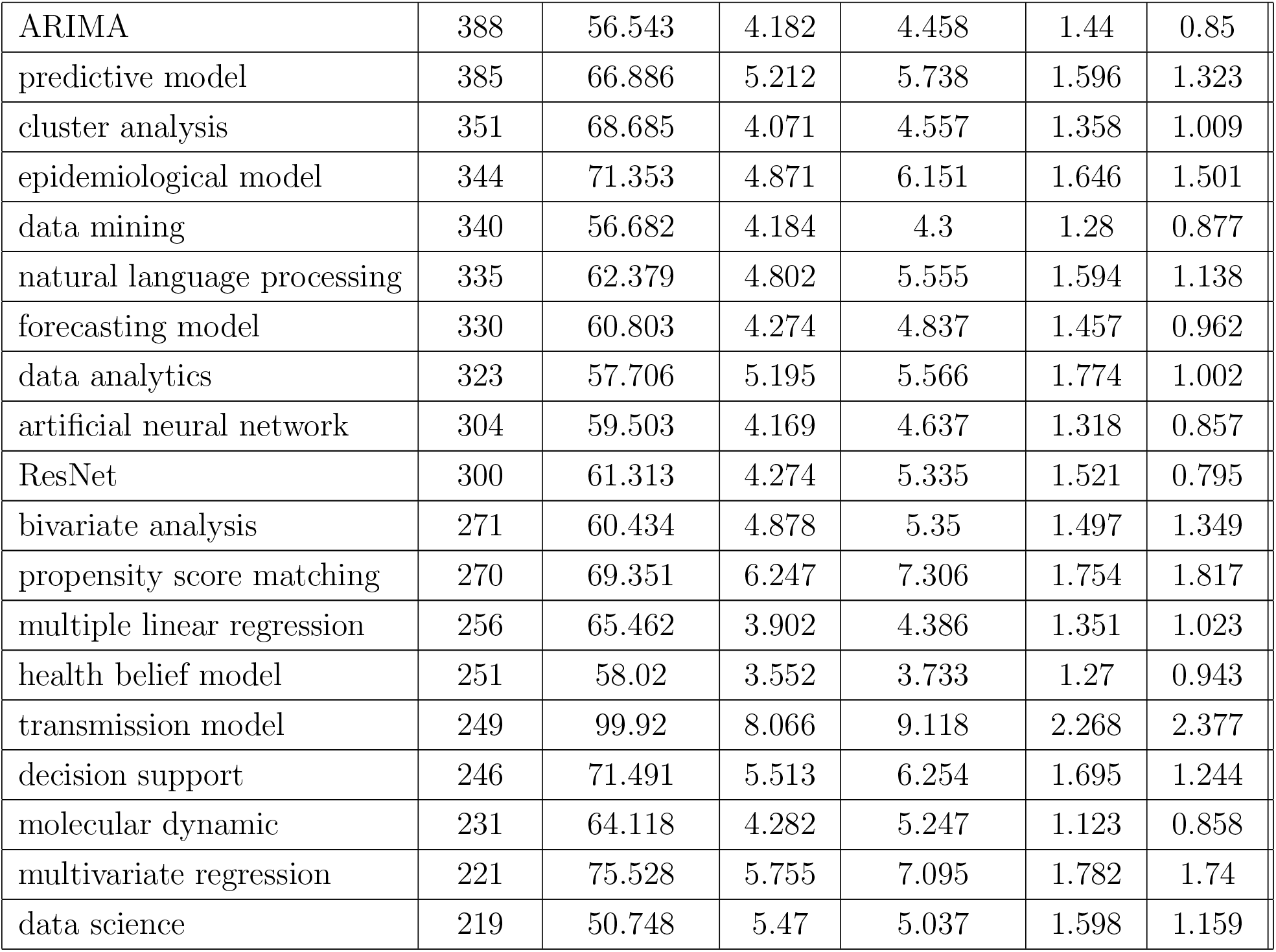
Top-50 modeling methods in modeling publications and their publication impact

The top-50 modeling keywords can be grouped into a few major specific modeling techniques: time-series analysis, statistical modeling, shallow machine learning, deep learning, epidemic modeling, and medical analytics. Typical modeling tasks include estimation and forecasting, classification, prediction, sentiment analysis.

- time-series analysis, including regression, linear regression, logistic regression, Cox regression, Poisson regression, linear model, ARIMA, multiple linear regression, and multivariate regression.
- statistical modeling, including multivariate statistics, statistical model, Bayesian, univariate statistics, and mathematical models such as structural equation model, forecasting model, and bivariate analysis.
- shallow machine learning, including neural networks and ANN, support vector machine, random forest, transfer learning, decision tree, classification model, predictive model, cluster analysis, sentiment analysis, and natural language processing.
- deep learning, including deep learning, CNN, LSTM, and ResNet.
- epidemic modeling, including epidemiological model, SIR, SEIR, and transmission model.
- medical and biomedical modeling, health belief model, and molecular dynamic.

Table 22 shows the detailed data of the distributions of cross-disciplinary/method modeling on COVID-19.

### 7.11 Top-10 monthly concerned problems in modeling COVID-19

Here, we analyze the temporal trend of mostly concerned problems in modeling COVID-19. We first extract the modeling keywords per the method introduced in Section 3.4, where we choose the top-10 mostly concerned problems. Then, per the publication date of each publication with the date available, we categorize their publications into each month from Jan 2020 to Mar 2022, respectively. This forms the monthly publications associated with top-10 modeling problems.

Figure 28 shows the monthly top-10 problems concerned in modeling publications on COVID-19 over months between Jan 2020 and Mar 2022. The top-10 problems are mental health, second wave, lockdown, vaccine, anxiety, risk factor, vaccination, X-ray, public health, and depression, where mental and public health dominate the publications with significant concern increase over the whole period, the same trend holds for second wave, vaccine and vaccination. In contrast, concerns on X-ray seem to decline over the recent year. Table 24 further shows the number of publications in modeling publications of monthly top-10 problems over the 27 months.

**Figure 28:**
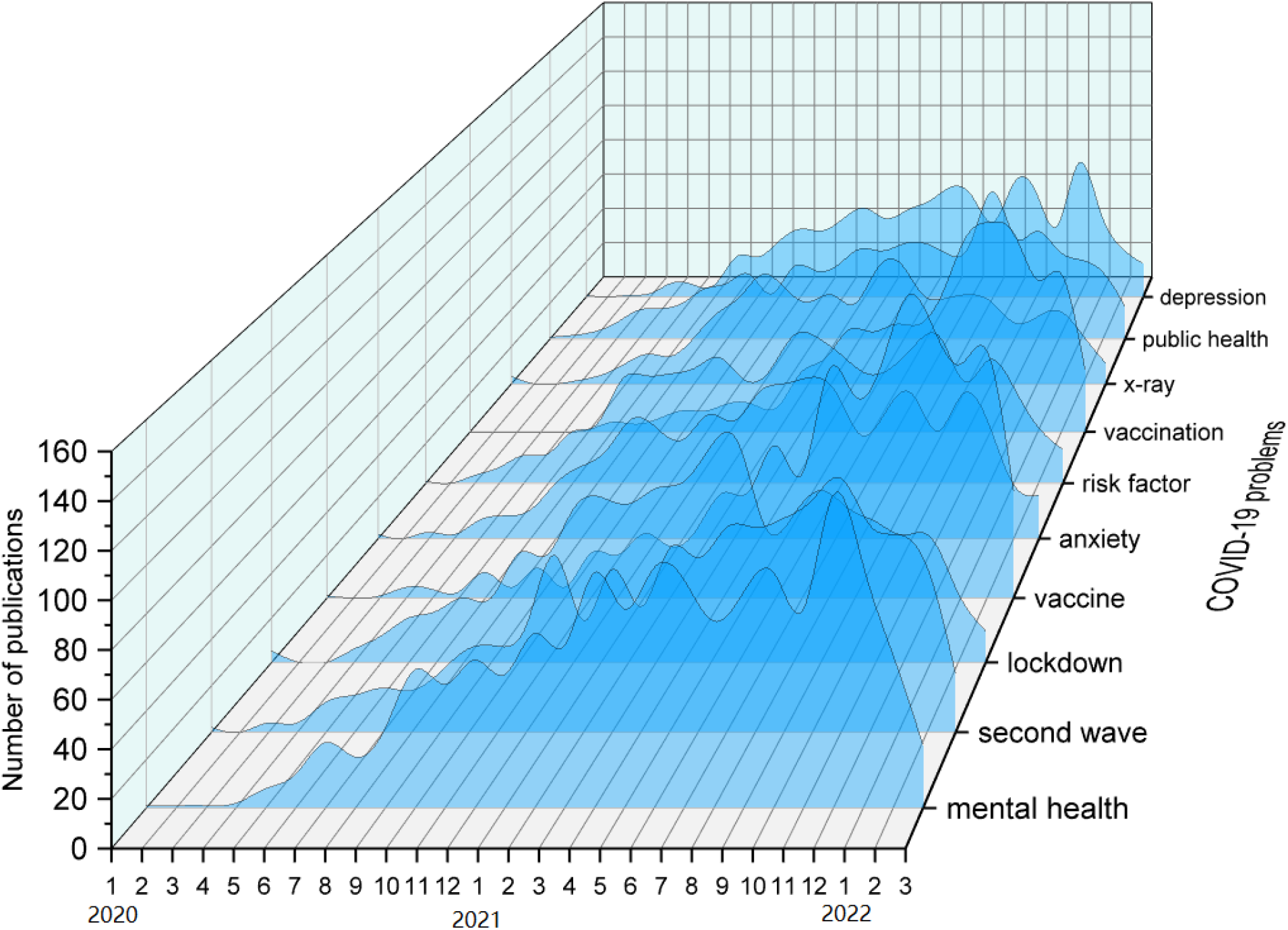
Top-10 monthly concerned problems in modeling publications

**Table 24:** Top-10 monthly concerned problems in modeling publications

|  | Mental health | Second wave | Lockdown | Vaccine | Anxiety | Risk factor | Vaccination | X-ray | Public health | Depression |
| --- | --- | --- | --- | --- | --- | --- | --- | --- | --- | --- |
| 2020-01 | 1 | 2 | 5 | 1 | 2 | 1 | 0 | 4 | 1 | 1 |
| 2020-02 | 1 | 0 | 0 | 0 | 0 | 0 | 0 | 0 | 2 | 0 |
| 2020-03 | 1 | 4 | 0 | 1 | 3 | 4 | 0 | 0 | 4 | 1 |
| 2020-04 | 2 | 4 | 6 | 5 | 2 | 8 | 0 | 2 | 9 | 2 |
| 2020-05 | 8 | 13 | 12 | 4 | 9 | 14 | 0 | 5 | 16 | 8 |
| 2020-06 | 14 | 16 | 20 | 3 | 11 | 15 | 1 | 11 | 17 | 7 |
| 2020-07 | 27 | 19 | 23 | 12 | 14 | 26 | 5 | 16 | 20 | 7 |
| 2020-08 | 21 | 18 | 29 | 5 | 27 | 30 | 4 | 15 | 28 | 23 |
| 2020-09 | 33 | 26 | 29 | 14 | 35 | 52 | 7 | 25 | 31 | 23 |
| 2020-10 | 57 | 36 | 49 | 5 | 43 | 54 | 11 | 38 | 36 | 32 |
| 2020-11 | 51 | 37 | 48 | 21 | 58 | 55 | 11 | 47 | 25 | 39 |
| 2020-12 | 61 | 46 | 69 | 20 | 53 | 58 | 15 | 59 | 40 | 38 |
| 2021-01 | 55 | 76 | 74 | 30 | 48 | 63 | 14 | 51 | 38 | 45 |
| 2021-02 | 72 | 48 | 71 | 24 | 60 | 52 | 13 | 44 | 41 | 51 |
| 2021-03 | 67 | 70 | 78 | 32 | 57 | 56 | 31 | 48 | 50 | 45 |
| 2021-04 | 97 | 54 | 81 | 49 | 71 | 74 | 38 | 43 | 49 | 50 |
| 2021-05 | 82 | 79 | 97 | 50 | 75 | 73 | 53 | 62 | 53 | 55 |
| 2021-06 | 99 | 75 | 98 | 71 | 78 | 64 | 52 | 65 | 53 | 63 |
| 2021-07 | 94 | 88 | 60 | 58 | 71 | 55 | 56 | 48 | 47 | 58 |
| 2021-08 | 77 | 88 | 66 | 106 | 50 | 58 | 56 | 46 | 53 | 43 |
| 2021-09 | 89 | 92 | 77 | 94 | 62 | 70 | 73 | 48 | 82 | 66 |
| 2021-10 | 97 | 100 | 69 | 109 | 70 | 73 | 102 | 44 | 57 | 60 |
| 2021-11 | 82 | 109 | 63 | 141 | 52 | 51 | 109 | 33 | 60 | 41 |
| 2021-12 | 129 | 89 | 57 | 125 | 70 | 62 | 103 | 36 | 49 | 77 |
| 2022-01 | 100 | 85 | 56 | 109 | 56 | 48 | 79 | 39 | 44 | 49 |
| 2022-02 | 63 | 71 | 31 | 115 | 26 | 28 | 82 | 26 | 37 | 28 |
| 2022-03 | 26 | 25 | 14 | 50 | 21 | 17 | 32 | 11 | 18 | 19 |

The visualization tool for generating this result is 3D Waterfall^43^. The data can be found in “Monthly top-10 business problems in modeling publications.xlsx”

### 7.12 Top-10 problems in top-10 published countries

In this section, we analyze the top-10 problems concerned by top-10 mostly published countries on modeling COVID-19. Similar to the approach taken in Section 7.11, we firstly extract the top-10 problem keywords from all extracted problem keywords in modeling publications following the problem keyword extraction method in Section 3.4. On the other hand, we count the number of total publications of each first-authored country per the author country extraction method in Section 3.2 and then extract the top-10 mostly published countries in all modeling publications. We then count the number of publications on each of the top-10 problems by each country.

Figure 29 shows the number of publications associated with top-10 problems concerned by each of the top-10 published countries. Table 25 further lists their publication numbers in detail. The mostly concerned top-10 problems are mental health, second wave, lockdown, vaccine, anxiety, risk factor, vaccination, X-ray, public health, and depression. The top-10 mostly published first-authored countries are the US, China, Italy, the UK, India, Spain, Canada, Germany, France, and Brazil.

**Figure 29:**
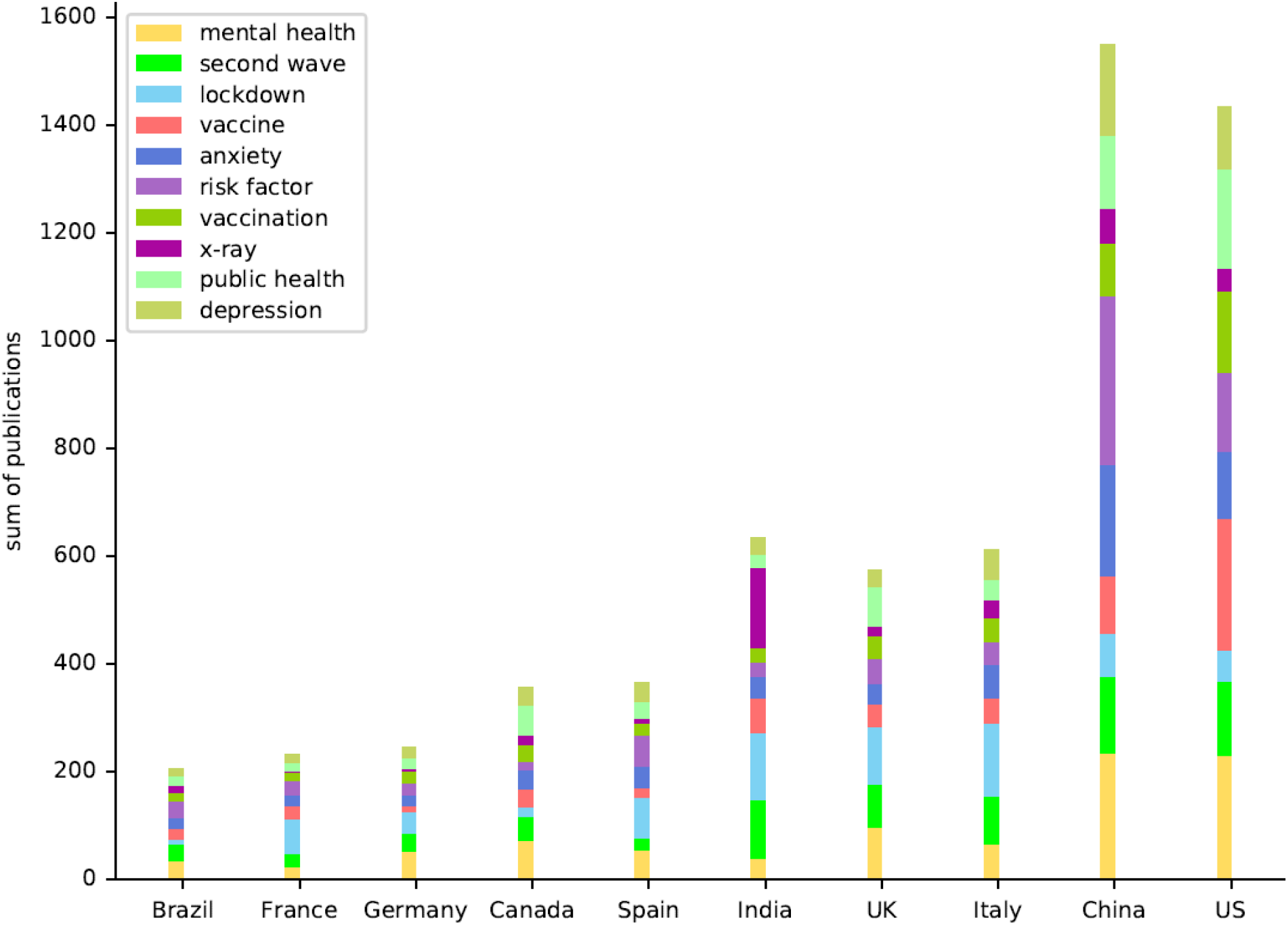
Top-10 problems in top-10 published countries in modeling publications

**Table 25:** Top-10 problems in top-10 published countries

| Country | Mental health | Second wave | Lockdown | Vaccine | Anxiety | Risk factor | Vaccination | X-ray | Public health | Depression |
| --- | --- | --- | --- | --- | --- | --- | --- | --- | --- | --- |
| US | 230 | 138 | 56 | 246 | 125 | 146 | 151 | 42 | 184 | 115 |
| China | 234 | 141 | 82 | 106 | 207 | 313 | 98 | 64 | 135 | 169 |
| Italy | 66 | 87 | 136 | 48 | 61 | 43 | 43 | 35 | 38 | 54 |
| UK | 96 | 79 | 108 | 42 | 37 | 47 | 43 | 18 | 72 | 33 |
| India | 38 | 110 | 123 | 65 | 40 | 27 | 26 | 150 | 23 | 31 |
| Spain | 53 | 23 | 76 | 18 | 40 | 57 | 22 | 10 | 30 | 36 |
| Canada | 71 | 46 | 17 | 33 | 35 | 17 | 30 | 19 | 54 | 35 |
| Germany | 52 | 34 | 39 | 12 | 20 | 22 | 21 | 6 | 18 | 20 |
| France | 23 | 24 | 65 | 23 | 20 | 27 | 17 | 2 | 15 | 15 |
| Brazil | 33 | 32 | 9 | 19 | 21 | 31 | 16 | 12 | 18 | 14 |

This result shows the difference and similarity of prioritized problems between countries. For example, vaccine/vaccination, mental health/anxiety/depression were on the list of top concerned problems by the American scientists. In China, more priority was placed on understanding the risk factors of COVID-19, with much less intensity on vaccine, although similar interest to the US is on mental health/depression. In contrast, Italy, UK, Spain, and France paid highest attention to lockdown. Interestingly, Indian researchers favored X-ray and then lockdown in their COVID-19 research.

Specifically, the US produced 1,433 publications to address the top-10 problems, in contrast to 1,549 by China.

The data can be found in “Top-10 business problems in top-10 countries in modeling publications.xlsx.”

### 7.13 Top-10 modeling methods in top-10 published countries

Similar to Section 7.12, here, we are concerned about the top-10 modeling methods mostly applied by the top-10 mostly published countries. We firstly extract the modeling publications as in Section 3.7 per the modeling keyword extraction method introduced in Section 3.5. We then extract the top-10 modeling keywords from the extracted modeling keyword list. Further, we extract the top-10 mostly published countries on modeling COVID-19, as shown in Sections 2.2 and 3.2.

Figure 30 shows the number of publications associated with the top-10 modeling methods mostly concerned by the top-10 countries with the highest number of modeling publications. Table 26 further lists their publication numbers in detail. The top-10 modeling methods are regression model, machine learning, simulation, linear regression, multivariate statistics, artificial intelligence, logistic regression, statistical model, deep learning, and CNN. The top-10 mostly published first-authored countries are the US, China, Italy, the UK, India, Spain, Canada, Germany, France, and Brazil.

**Figure 30:**
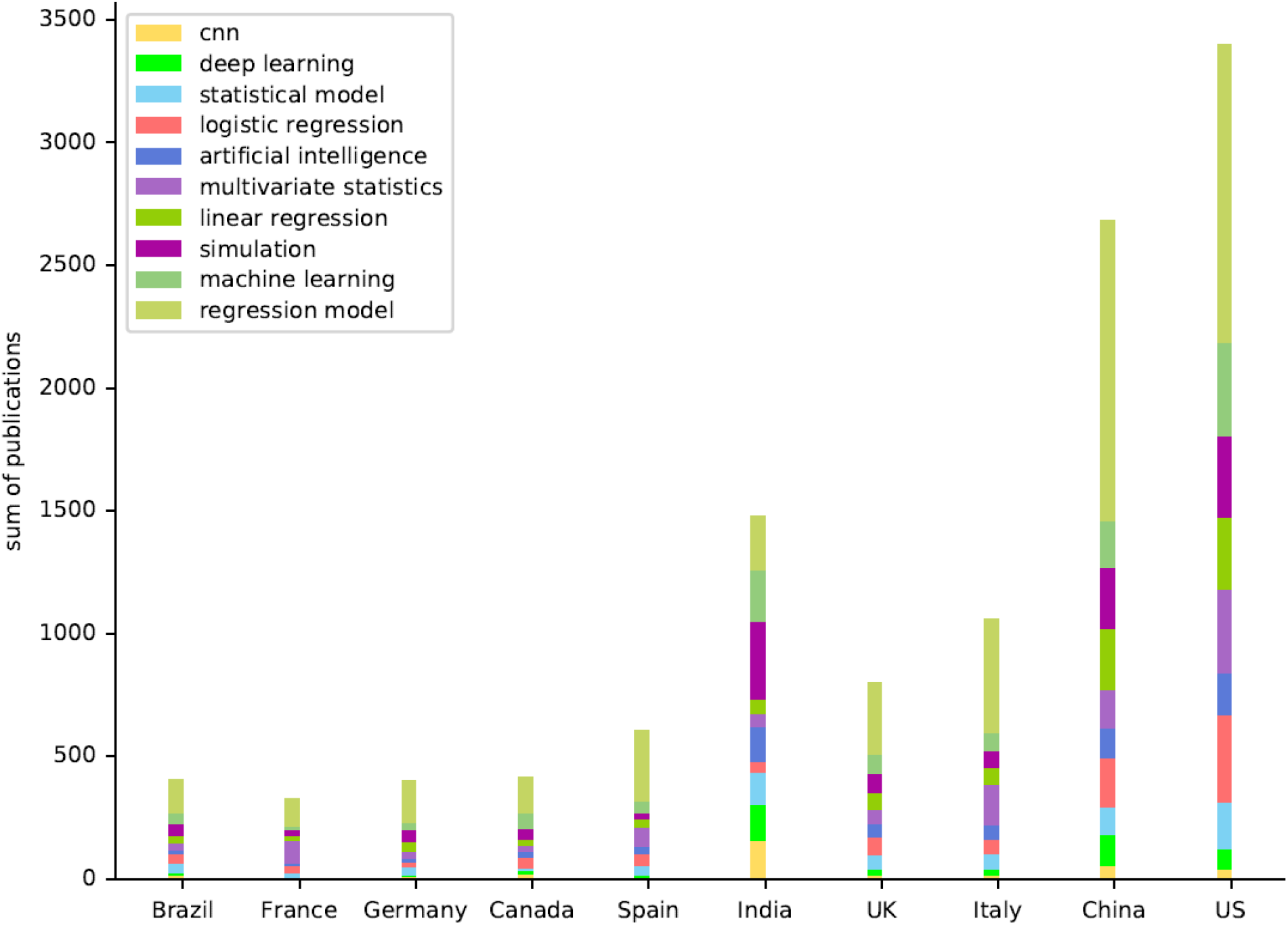
Top-10 modeling methods in top-10 published countries in modeling publications

**Table 26:** Top-10 modeling methods in top-10 published countries in modeling publications

| Country | Regression model | Machine learning | Simulation | Linear regression | Multivariate statistics | Artificial intelligence | Logistic regression | Statistical model | Deep learning | CNN |
| --- | --- | --- | --- | --- | --- | --- | --- | --- | --- | --- |
| US | 1211 | 384 | 330 | 290 | 343 | 172 | 355 | 188 | 84 | 41 |
| China | 1223 | 193 | 250 | 248 | 157 | 121 | 199 | 114 | 124 | 55 |
| Italy | 463 | 70 | 72 | 68 | 167 | 57 | 57 | 63 | 25 | 16 |
| UK | 293 | 77 | 78 | 68 | 58 | 54 | 72 | 61 | 23 | 15 |
| India | 219 | 213 | 317 | 57 | 51 | 145 | 42 | 134 | 144 | 157 |
| Spain | 285 | 51 | 23 | 36 | 74 | 30 | 49 | 39 | 11 | 6 |
| Canada | 149 | 60 | 44 | 24 | 26 | 23 | 43 | 12 | 16 | 19 |
| Germany | 173 | 27 | 50 | 40 | 29 | 14 | 20 | 34 | 6 | 9 |
| France | 109 | 14 | 26 | 19 | 95 | 10 | 30 | 17 | 4 | 2 |
| Brazil | 136 | 42 | 50 | 28 | 32 | 15 | 40 | 39 | 9 | 14 |

The results show some interesting preference difference and similarity across the top-10 countries. Almost all countries favor and heavily rely on regression models in studying COVID-19. Not all of the top-10 modeling methods selected from all of the modeling publications appear in the top-10 of all countries, for example, deep learning including CNNs were mainly applied in India, China, and the US. Both American and Chinese researchers heavily applied regression models in comparison with other techniques. Indians seem to like simulation much more than other countries, although they all made simulation in their mostly applied technique. Classic machine learning and AI methods were widely applied by all countries as a major tool of studying COVID-19, which seem to be particularly favored by Indians and Americans. The full details can be found in “Top-10 modeling methods globally in modeling publications.xlsx.”

### 7.14 Top-10 problems and top-10 modeling methods by top-10 published countries

Here, we study the interactions between top-10 modeling methods, top-10 modeled problems, and the top-10 published countries, and measure their publication impact. Taken the modeling keyword extraction method in Section 3.5, problem keyword extraction method in Section 3.4, modeling publication extraction method in Section 3.7, and first-authored country extraction in Section 3.2, we generate the top-10 modeling keywords, top-10 problem keywords, and top-10 published countries respectively from the modeling publications.

Figure 31 shows the results of the top-10 modeling methods interacted with top-10 problems concerned by top-10 published countries with the highest number of their modeling publications. The mean of the collective impact metrics CI measures the mean publication impact of each country in terms of integrating H5-index, Impact Factor, CiteScore, SNIP, and SJR. CI is calculated per the method in Section 4.2. There are two pairwise bars for each country, where the left bars refer to the number of publications associated with each of the top-10 problems concerned by each country, the right bars refer to the number of publications corresponding to each of the top-10 modeling methods applied by each country. There are two Y axes in the diagram. The left Y axis refers to two pairwise bars, showing the number of publications corresponding to each of top-10 problems and each of the top-10 modeling methods concerned by each country, we stack the publications over all 10 problems and 10 modeling methods respectively for each country to obtain their total number of publications across all top-10 problems and top-10 modeling methods for each country. The right Y axis refers to the mean CI, showing the publication-averaged composite indicator value of each country on those top-10 problems and top-10 modeling methods jointly.

**Figure 31:**
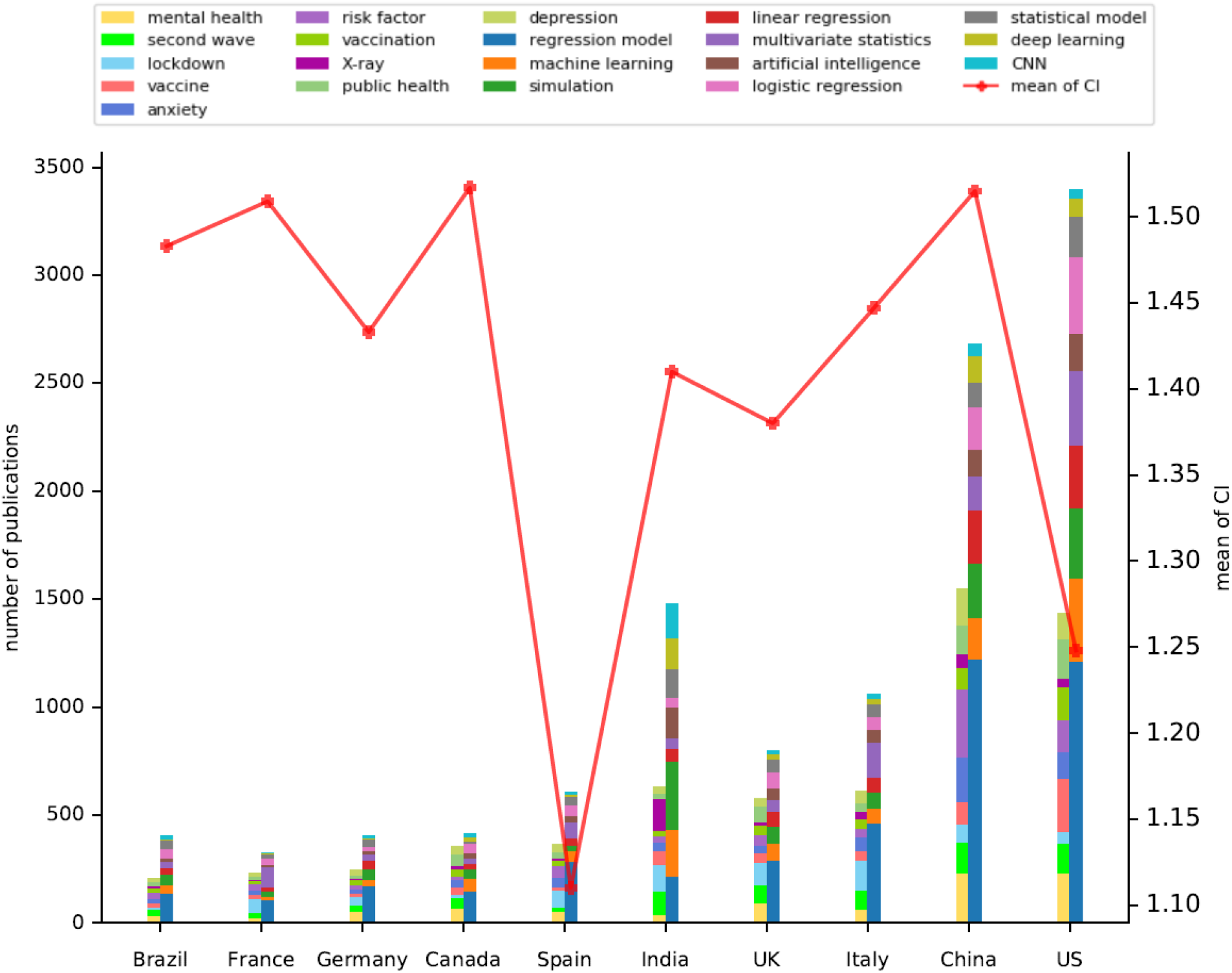
Top-10 problems and top-10 modeling methods by top-10 published countries with mean composite indicator

The mostly concerned top-10 problems are mental health, second wave, lockdown, vaccine, anxiety, risk factor, vaccination, X-ray, public health, and depression. The top-10 modeling methods are regression model, machine learning, simulation, linear regression, multivariate statistics, artificial intelligence, logistic regression, statistical model, deep learning, and CNN. The top-10 mostly published first-authored countries are the US, China, Italy, the UK, India, Spain, Canada, Germany, France, and Brazil.

In addition to the findings discussed in Section 7.12 (e.g., Figure 29) on top-10 problems by top-10 countries, and that in Section 7.13 (e.g., Figure 30) on top-10 modeling methods by top-10 countries, here, we can further find out the significant inconsistency between publication numbers and their mean publication impact in terms of mean CI across countries. The UK did not make the most contributions in terms of the number of modeling publications, while they mean CI is the highest across the top-10 published countries. India unfortunately marks the lowest impact although their overall number of publications ranks the third. France made the second lowest number of publications among the top-10 countries, however, their mean CI ranks the second. There seems to be a strong correlation between the quality and quantity of COVID-19 modeling publications and their economic status, as further investigated in Section 4.4 on the correlation between the COVID-19 publications and GDP per capita.

The results for the top-10 problems concerned by all countries is in the file “Top-10 business problems globally in modeling publications.xlsx;” and the results for the top-10 modeling methods applied by all countries is in the file “Top-10 modeling methods globally in modeling publications.xlsx.”

## 8 Correlation between publications, economy and infection

This chapter fulfils the fourth aims and objectives discussed in Section 1.1 on understanding the relations between COVID-19 research and a country’s COVID-19 development and their economic status. We thus explore a few questions here, including the correlations between publications and GDP, the correlations between coronavirus infections and resultant deaths and publications globally and in G20 and OECD countries and regions, respectively.

### 8.1 Correlation between global publications and GDP per capita

Here, we analyze the correlation between the number of publications and GDP per capita, and between the cumulative composite indicator and GDP per capita for each first-authored country. First, we extract all publications for each first-authored country per the method introduced in Section 3.2. Then, we calculate the cumulative composite indicator value of all publications from each first-authored country per the method in Section 4.2. In addition, we obtain the GDP and population data for each country per the method introduced in Section 3.2 and then calculate the GDP per capita (GDPPC) per the measurement in Section 4.3.

Further, we calculate the correlation between the number of publications and the GDPPC for each country per the absolute and relative publication-GDP correlation coefficients, and the absolute and relative publication composite impact-GDP coefficients between the cumulative composite indicator and the GDPPC of a country, respectively, as introduced in Section 4.4.

As a result, Figure 32 shows the results of the absolute publication composite impact-GDP coefficient, Figure 33 shows the results of the relative publication composite impact-GDP coefficient, Figure 34 shows the results of the dispersion publication composite impact-GDP coefficient of the global COVID-19 research, visualized by world map. Table 27 further lists the top-10 mostly correlated countries in terms of absolute, relative and dispersion publication composite impact-GDP correlation coefficients, respectively. The results can be found in file ‘Global countries’ publications and GDP per capita.xlsx.’

**Figure 32:**
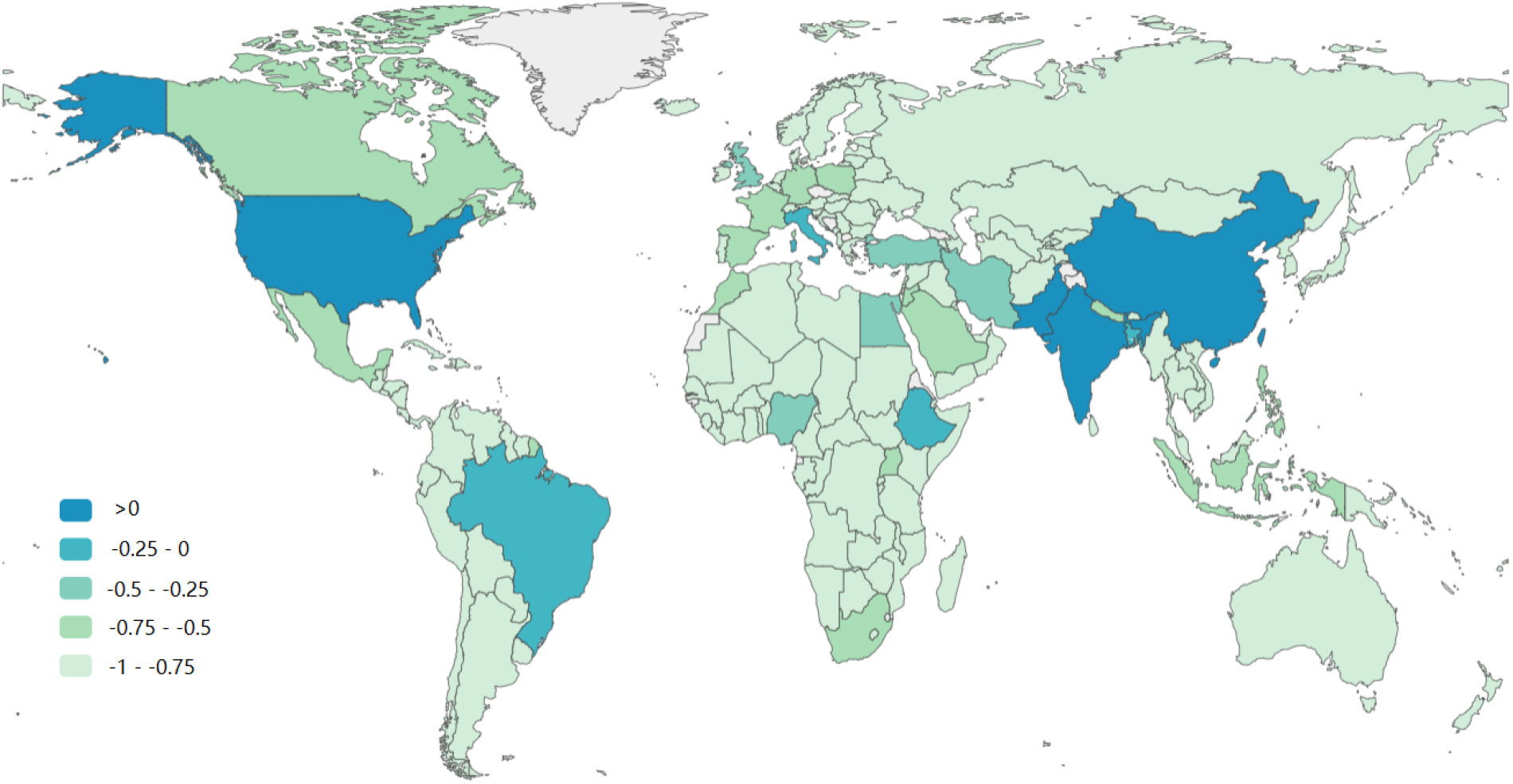
Global absolute publication composite impact-GDP coefficient

**Figure 33:**
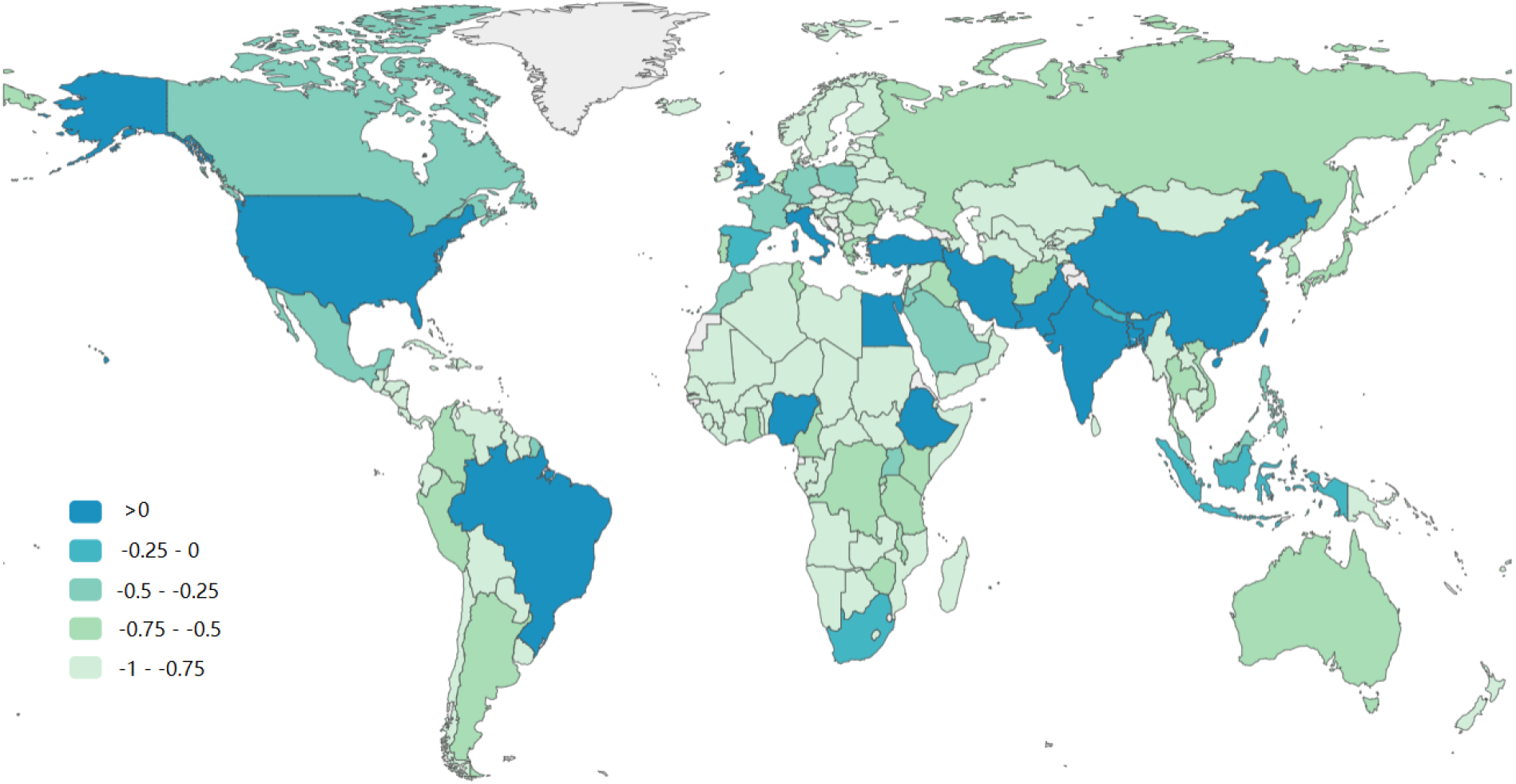
Global relative publication composite impact-GDP coefficient

**Figure 34:**
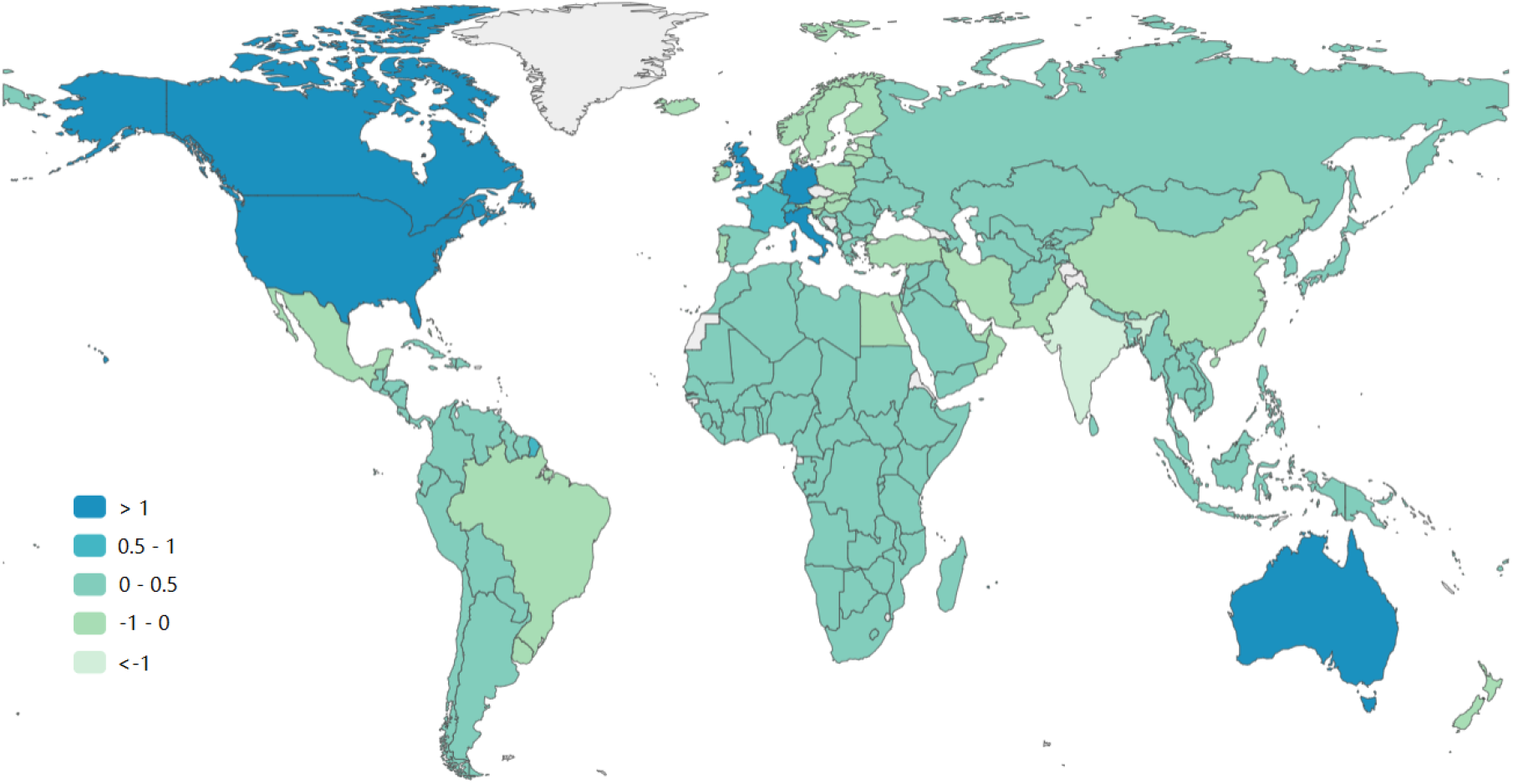
Global dispersion publication composite impact-GDP coefficient

**Table 27:** Correlation between top 10 countries’ publications and GDP per capita

| Country | Num_pub<br>$N$ | GDPPC<br>$\bar{G}_i$ | CI<br>$\mathcal{C}_i$ | $\rho_n$ | $\tilde{\rho}_n$ | $\hat{\rho}_n$ | $\rho_c$ | $\tilde{\rho}_c$ | $\hat{\rho}_c$ |
| --- | --- | --- | --- | --- | --- | --- | --- | --- | --- |
| US | 49,203 | 61763 | 67854 | -0.113 | 0.483 | 19.786 | 0.047 | 0.483 | 20.061 |
| China | 21,693 | 10423 | 30735 | 0.351 | 0.765 | -0.858 | 0.494 | 0.77 | -0.897 |
| Italy | 15,029 | 32031 | 20165 | -0.361 | 0.257 | 2.063 | -0.227 | 0.244 | 2.04 |
| UK | 14,475 | 40867 | 20422 | -0.477 | 0.122 | 3.017 | -0.334 | 0.133 | 3.152 |
| India | 13,044 | 1882 | 13215 | 0.748 | 0.923 | -1.39 | 0.751 | 0.897 | -1.006 |
| Spain | 6,372 | 26924 | 7908 | -0.617 | -0.079 | 0.538 | -0.546 | -0.131 | 0.487 |
| Canada | 6,093 | 42789 | 7702 | -0.751 | -0.322 | 1.194 | -0.695 | -0.36 | 1.106 |
| Germany | 5,857 | 46100 | 7797 | -0.775 | -0.372 | 1.273 | -0.711 | -0.387 | 1.258 |
| France | 5,196 | 40571 | 7108 | -0.773 | -0.368 | 0.897 | -0.702 | -0.372 | 0.919 |
| Brazil | 5,024 | 6728 | 5580 | -0.145 | 0.458 | -0.285 | -0.093 | 0.369 | -0.221 |

These results show that

- Germany, France and Canada show the lowest correlation coefficient between their publication numbers and their GDP per capita and their publication impact in terms of cumulative composite indicator (CI). This shows that these countries have the lowest productivity in terms of COVID-19 publication number and impact in the context of their much greater GDP per capita.
- In contrast, India shows the highest correlation coefficient between their publication number and GDP per capita and publication impact, while China ranks the second highest among the top-10 mostly published countries. This means India and China show the highest productivity of producing COVID-19 research outcomes in the context of their lower GDP per capita.

### 8.2 Correlation between G20 publications and GDP per capita

Specifically, here, we analyze the correlation between the COVID-19 publications and their GDP per capita for those G20 countries and regions. We take the same approach as in Section 8.1 to obtain the total publications, the cumulative composite indicator of all publications, and the GDP per capita for each G20 country.

Table 28 shows the absolute and relative publication-GDP coefficients, and the absolute and relative publication composite impact-GDP coefficients over the G20 countries. The measures and their calculations in the table are introduced in Section 4.4. The full data can be found in “G20 publications and GDP per capita.xlsx.”

**Table 28:** Correlation between G20 publications and GDP per capita

| Country | Num_pub<br>$N$ | GDPPC<br>$\overline{G}_i$ | CI<br>$C_i$ | $\rho_n$ | $\tilde{\rho}_n$ | $\hat{\rho}_n$ | $\rho_c$ | $\tilde{\rho}_c$ | $\hat{\rho}_c$ |
| --- | --- | --- | --- | --- | --- | --- | --- | --- | --- |
| US | 49,203 | 61763 | 67854 | -0.113 | 0 | 7.242 | 0.047 | 0 | 7.244 |
| China | 21,693 | 10423 | 30735 | 0.351 | 0.446 | -0.919 | 0.494 | 0.457 | -0.983 |
| Italy | 15,029 | 32031 | 20165 | -0.361 | -0.259 | 0.229 | -0.227 | -0.271 | 0.229 |
| UK | 14,475 | 40867 | 20422 | -0.477 | -0.384 | 0.467 | -0.334 | -0.375 | 0.524 |
| India | 13,044 | 1882 | 13215 | 0.748 | 0.794 | -0.51 | 0.751 | 0.729 | -0.179 |
| Canada | 6,093 | 42789 | 7702 | -0.751 | -0.697 | -0.201 | -0.695 | -0.718 | -0.201 |
| Germany | 5,857 | 46100 | 7797 | -0.775 | -0.725 | -0.262 | -0.711 | -0.733 | -0.231 |
| France | 5,196 | 40571 | 7108 | -0.773 | -0.723 | -0.244 | -0.702 | -0.725 | -0.208 |
| Brazil | 5,024 | 6728 | 5580 | -0.145 | -0.032 | 0.296 | -0.093 | -0.14 | 0.335 |
| Australia | 4,719 | 51851 | 6152 | -0.833 | -0.795 | -0.481 | -0.788 | -0.805 | -0.447 |
| Turkey | 4,600 | 8436 | 4421 | -0.294 | -0.187 | 0.301 | -0.312 | -0.354 | 0.369 |
| Japan | 3,678 | 40662 | 4305 | -0.834 | -0.796 | -0.361 | -0.809 | -0.824 | -0.363 |
| Saudi Arabia | 2,792 | 19319 | 3156 | -0.747 | -0.693 | 0.149 | -0.719 | -0.741 | 0.148 |
| South Korea | 2,696 | 31617 | 3643 | -0.843 | -0.807 | -0.187 | -0.793 | -0.81 | -0.171 |
| Mexico | 1,436 | 8526 | 1594 | -0.712 | -0.651 | 0.548 | -0.685 | -0.709 | 0.526 |
| South Africa | 1,339 | 5599 | 1464 | -0.614 | -0.538 | 0.655 | -0.585 | -0.616 | 0.629 |
| Russia | 1,323 | 10250 | 1045 | -0.771 | -0.721 | 0.498 | -0.815 | -0.83 | 0.498 |
| Indonesia | 1,252 | 3847 | 1087 | -0.509 | -0.42 | 0.724 | -0.559 | -0.591 | 0.713 |
| Argentina | 590 | 8549 | 586 | -0.871 | -0.841 | 0.613 | -0.872 | -0.883 | 0.582 |

The results show that

- Argentina, South Korea and Japan rank the top-3 with the lowest correlation between their publication number and GDP per capita. This indicates that these countries have the lowest productivity of producing COVID-19 publication number in the context of their GDP per capita.
- Argentina, Russia and Japan rank the top-3 with the lowest correlation between their overall publication impact (i.e., CI) and GDP per capita (GDPPC). This shows that these countries have the lowest COVID-19 research impact in terms of their GDP per capita.
- In contrast, India, China and the US rank top-3 in terms of their correlation coefficient between their publication number and GDP per capita, and their overall publication impact (i.e., CI) and their GDP per capita (GDPPC). This shows that these countries have the highest productivity and impact of COVID-19 research in terms of their GDP per capita.

### 8.3 Correlation between OECD publications and GDP per capita

Similar to the analysis in Section 8.2 for G20 countries and regions, here, we further analyze the correlation between OECD country’s publications and their GDP per capita for each country and region.

Table 29 shows the absolute and relative publication-GDP coefficients, and the absolute and relative publication composite impact-GDP coefficients over the OECD countries. The symbols and their calculations in the table are introduced in Section 4.4. The details can be found in “OECD publications and GDP per capita.xlsx.”

**Table 29:** Correlation between OECD publications and GDP per capita

| Country | Num_pub<br>$N$ | GDPPC<br>$\bar{G}_i$ | CI<br>$C_i$ | $\rho_n$ | $\tilde{\rho}_n$ | $\hat{\rho}_n$ | $\rho_c$ | $\tilde{\rho}_c$ | $\hat{\rho}_c$ |
| --- | --- | --- | --- | --- | --- | --- | --- | --- | --- |
| US | 49,203 | 61763 | 67854 | -0.113 | 0.294 | 5.462 | 0.047 | 0.294 | 5.48 |
| Turkey | 4,600 | 8436 | 4421 | -0.294 | 0.113 | -0.144 | -0.312 | -0.067 | 0.051 |
| Italy | 15,029 | 32031 | 20165 | -0.361 | 0.038 | -0.378 | -0.227 | 0.024 | -0.369 |
| UK | 14,475 | 40867 | 20422 | -0.477 | -0.102 | 0.132 | -0.334 | -0.091 | 0.138 |
| Spain | 6,372 | 26924 | 7908 | -0.617 | -0.295 | -0.161 | -0.546 | -0.342 | -0.131 |
| Mexico | 1,436 | 8526 | 1594 | -0.712 | -0.442 | 0.346 | -0.685 | -0.525 | 0.368 |
| Canada | 6,093 | 42789 | 7702 | -0.751 | -0.507 | 0.054 | -0.695 | -0.538 | 0.045 |
| Poland | 2,012 | 14969 | 2335 | -0.763 | -0.528 | 0.201 | -0.73 | -0.587 | 0.224 |
| France | 5,196 | 40571 | 7108 | -0.773 | -0.545 | 0.016 | -0.702 | -0.548 | 0.017 |
| Germany | 5,857 | 46100 | 7797 | -0.775 | -0.548 | 0.086 | -0.711 | -0.56 | 0.083 |
| Colombia | 584 | 5213 | 642 | -0.799 | -0.59 | 0.53 | -0.781 | -0.659 | 0.527 |
| Australia | 4,719 | 51851 | 6152 | -0.833 | -0.654 | 0.072 | -0.788 | -0.67 | 0.064 |
| Japan | 3,678 | 40662 | 4305 | -0.834 | -0.656 | 0 | -0.809 | -0.7 | -0.005 |
| South Korea | 2,696 | 31617 | 3643 | -0.843 | -0.672 | 0.035 | -0.793 | -0.678 | 0.032 |
| Greece | 1,491 | 18169 | 1738 | -0.848 | -0.682 | 0.229 | -0.825 | -0.725 | 0.238 |
| Portugal | 1,173 | 22233 | 1391 | -0.9 | -0.784 | 0.209 | -0.882 | -0.811 | 0.212 |
| Chile | 574 | 12880 | 654 | -0.915 | -0.814 | 0.409 | -0.903 | -0.844 | 0.404 |
| Netherlands | 2,123 | 51989 | 3161 | -0.922 | -0.828 | -0.108 | -0.885 | -0.816 | -0.087 |
| Israel | 1,766 | 45042 | 2453 | -0.925 | -0.835 | -0.064 | -0.897 | -0.833 | -0.059 |
| Belgium | 1,424 | 44736 | 2001 | -0.938 | -0.864 | -0.072 | -0.914 | -0.861 | -0.067 |
| Austria | 1,096 | 48426 | 1476 | -0.956 | -0.901 | -0.132 | -0.941 | -0.903 | -0.126 |
| Switzerland | 1,880 | 86007 | 2632 | -0.957 | -0.904 | -0.439 | -0.941 | -0.903 | -0.399 |
| Sweden | 1,067 | 51329 | 1488 | -0.959 | -0.909 | -0.173 | -0.944 | -0.908 | -0.163 |
| Hungary | 298 | 15632 | 355 | -0.963 | -0.916 | 0.398 | -0.956 | -0.927 | 0.386 |
| Czech Republic | 390 | 23379 | 466 | -0.967 | -0.926 | 0.256 | -0.961 | -0.936 | 0.249 |
| New Zealand | 599 | 40634 | 711 | -0.971 | -0.934 | -0.034 | -0.966 | -0.943 | -0.033 |
| Denmark | 809 | 60535 | 1157 | -0.974 | -0.94 | -0.326 | -0.962 | -0.938 | -0.307 |
| Ireland | 1,131 | 84718 | 1341 | -0.974 | -0.94 | -0.606 | -0.969 | -0.949 | -0.611 |
| Norway | 639 | 66650 | 835 | -0.981 | -0.957 | -0.44 | -0.975 | -0.959 | -0.426 |
| Finland | 435 | 49022 | 561 | -0.982 | -0.96 | -0.176 | -0.977 | -0.963 | -0.17 |
| Slovakia | 164 | 18621 | 169 | -0.983 | -0.96 | 0.359 | -0.982 | -0.97 | 0.35 |
| Slovenia | 206 | 25260 | 257 | -0.984 | -0.963 | 0.236 | -0.98 | -0.967 | 0.228 |
| Lithuania | 108 | 20546 | 134 | -0.99 | -0.976 | 0.33 | -0.987 | -0.978 | 0.318 |
| Latvia | 67 | 18166 | 70 | -0.993 | -0.983 | 0.378 | -0.992 | -0.987 | 0.365 |
| Costa Rica | 44 | 11995 | 36 | -0.993 | -0.983 | 0.496 | -0.994 | -0.99 | 0.479 |
| Estonia | 51 | 23095 | 53 | -0.996 | -0.99 | 0.287 | -0.995 | -0.992 | 0.278 |
| Luxembourg | 77 | 113178 | 89 | -0.999 | -0.997 | -1.385 | -0.998 | -0.997 | -1.337 |
| Iceland | 26 | 57992 | 50 | -0.999 | -0.998 | -0.367 | -0.998 | -0.997 | -0.352 |

The results show that

- Luxembourg, Iceland and Estonia rank the lowest correlation between their publication number and GDP per capita. These countries show the lowest productivity of COVID-19 publications in terms of their GDP per capita.
- In contrast, the US, Turkey and Italy rank top 3 in terms of of their correlation coefficient between their publication number and GDP per capita; showing their highest productivity in terms of their economic status.
- With regard to the correlation between cumulative composite indicator (CI) and their GDP per capita (GDPPC), Luxembourg, Iceland and Estonia rank the lowest cor relation, showing the lowest productivity in COVID-19 publications; while the US, Turkey and Italy rank the highest three, showing their highest productivity in terms of COVID-19 research impact in terms of their economic conditions.

### 8.4 Global correlation between publications and infections

Here, we analyze the correlation between the COVID-19 publications and the number of infected cases in each country. The publications of each first-authored country are collected per the method introduced in Section 3.2, the collection of COVID-19 infected cases is done per the method introduced in Section 3.9. The correlation between publications and infections for each country is calculated per the correlation measures introduced in Section 4.5, i.e., Equation (14).

Figure 35 shows the correlation coefficient between publication number and case number in all the countries. Table 30 shows the detailed data of the correlation coefficient in top-20 countries. The results can be found in file ‘Global correlation between publications, infections and deaths.xlsx.’

**Figure 35:**
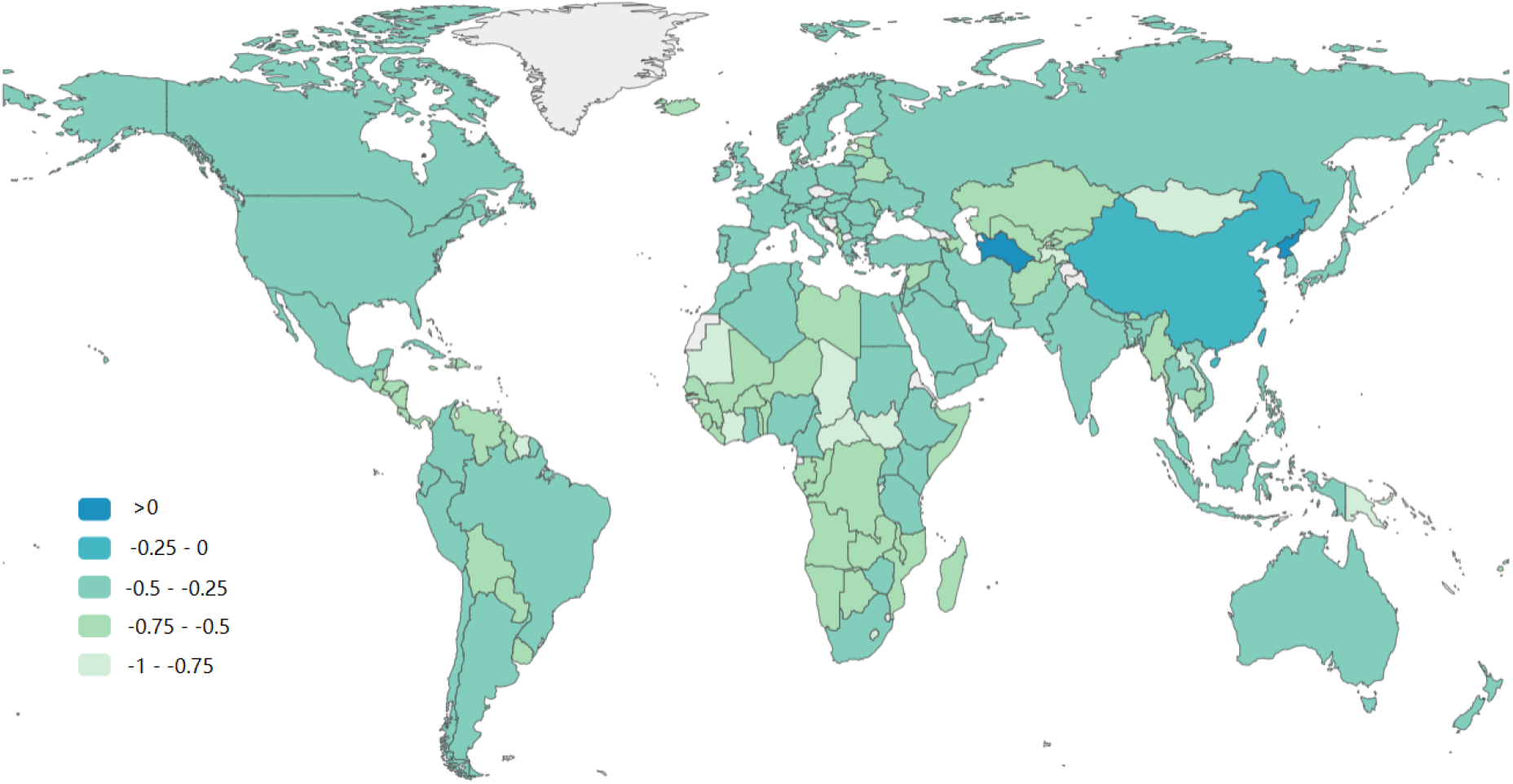
Correlation coefficient between publication number and new case number in all the countries

**Table 30:**
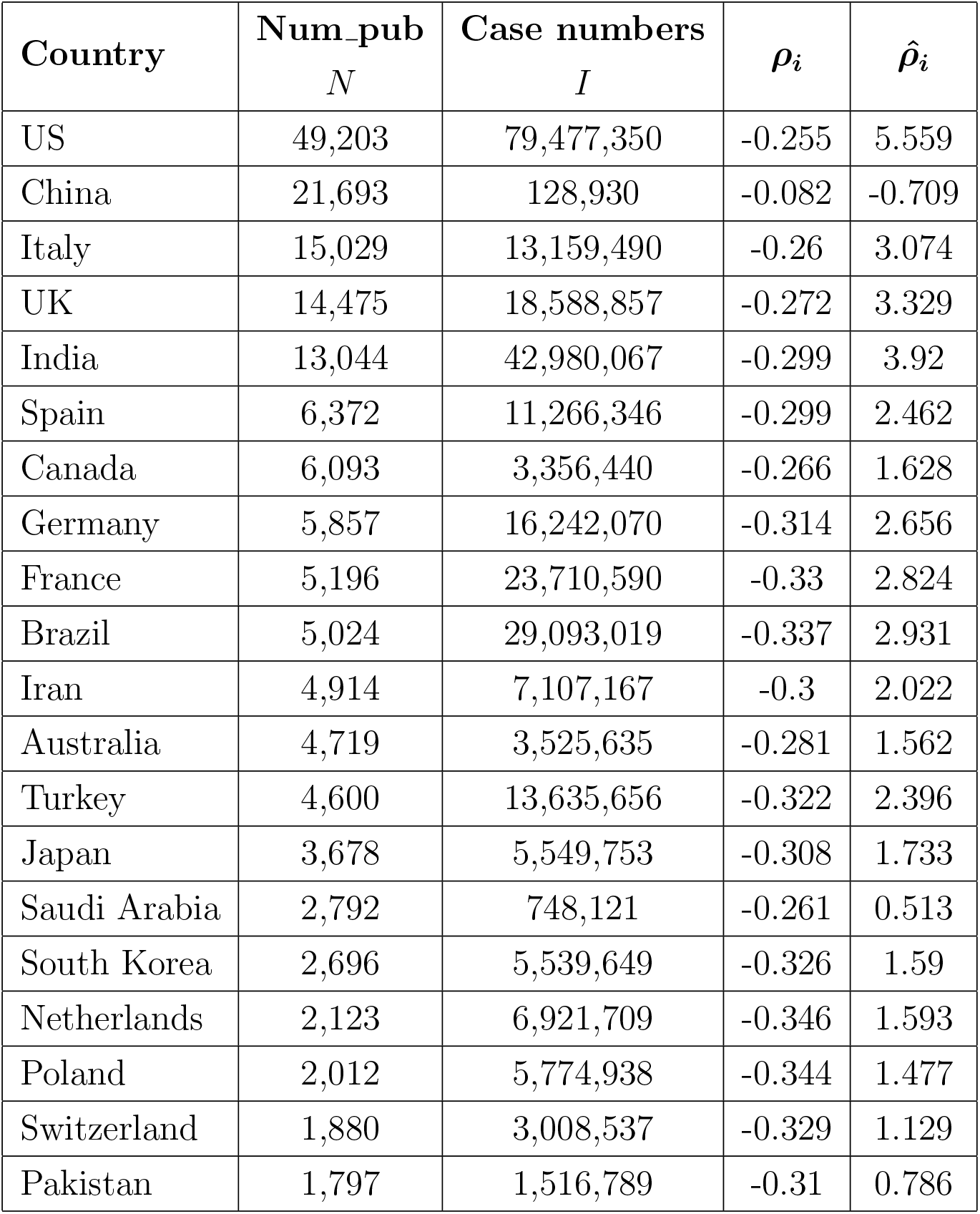
Correlation between publications and infections in top-20 countries

| Country | Num_pub<br>$N$ | Case numbers<br>$I$ | $\rho_i$ | $\hat{\rho}_i$ |
| --- | --- | --- | --- | --- |
| US | 49,203 | 79,477,350 | -0.255 | 5.559 |
| China | 21,693 | 128,930 | -0.082 | -0.709 |
| Italy | 15,029 | 13,159,490 | -0.26 | 3.074 |
| UK | 14,475 | 18,588,857 | -0.272 | 3.329 |
| India | 13,044 | 42,980,067 | -0.299 | 3.92 |
| Spain | 6,372 | 11,266,346 | -0.299 | 2.462 |
| Canada | 6,093 | 3,356,440 | -0.266 | 1.628 |
| Germany | 5,857 | 16,242,070 | -0.314 | 2.656 |
| France | 5,196 | 23,710,590 | -0.33 | 2.824 |
| Brazil | 5,024 | 29,093,019 | -0.337 | 2.931 |
| Iran | 4,914 | 7,107,167 | -0.3 | 2.022 |
| Australia | 4,719 | 3,525,635 | -0.281 | 1.562 |
| Turkey | 4,600 | 13,635,656 | -0.322 | 2.396 |
| Japan | 3,678 | 5,549,753 | -0.308 | 1.733 |
| Saudi Arabia | 2,792 | 748,121 | -0.261 | 0.513 |
| South Korea | 2,696 | 5,539,649 | -0.326 | 1.59 |
| Netherlands | 2,123 | 6,921,709 | -0.346 | 1.593 |
| Poland | 2,012 | 5,774,938 | -0.344 | 1.477 |
| Switzerland | 1,880 | 3,008,537 | -0.329 | 1.129 |
| Pakistan | 1,797 | 1,516,789 | -0.31 | 0.786 |

The correlation between COVID-19 publications and infections show that, of the top-20 publishing countries:

- China shows the highest log-correlation between their COVID-19 publication number and infections, showing their highest productivity in the context of their infections;
- Netherlands shows the lowest log-correlation between their COVID-19 publication number and infections, showing their lowest productivity in the context of their infections.

### 8.5 Global correlation between publications and deaths

Here, we analyze the correlation between the COVID-19 publications and the number of death cases in each country. The publications of each first-authored country are collected per the method introduced in Section 3.2, the collection of COVID-19 death cases is done per the method introduced in Section 3.9. The correlation between publications and deaths for each country is calculated per the correlation measures introduced in Section 4.5, i.e., Equation (14).

Figure 36 shows the correlation coefficient between publication number and death number in all the countries. Table 31 shows the detailed data of the correlation coefficient in top-20 countries. The results can be found in file ‘Global correlation between publications, infections and deaths.xlsx.’

**Figure 36:**
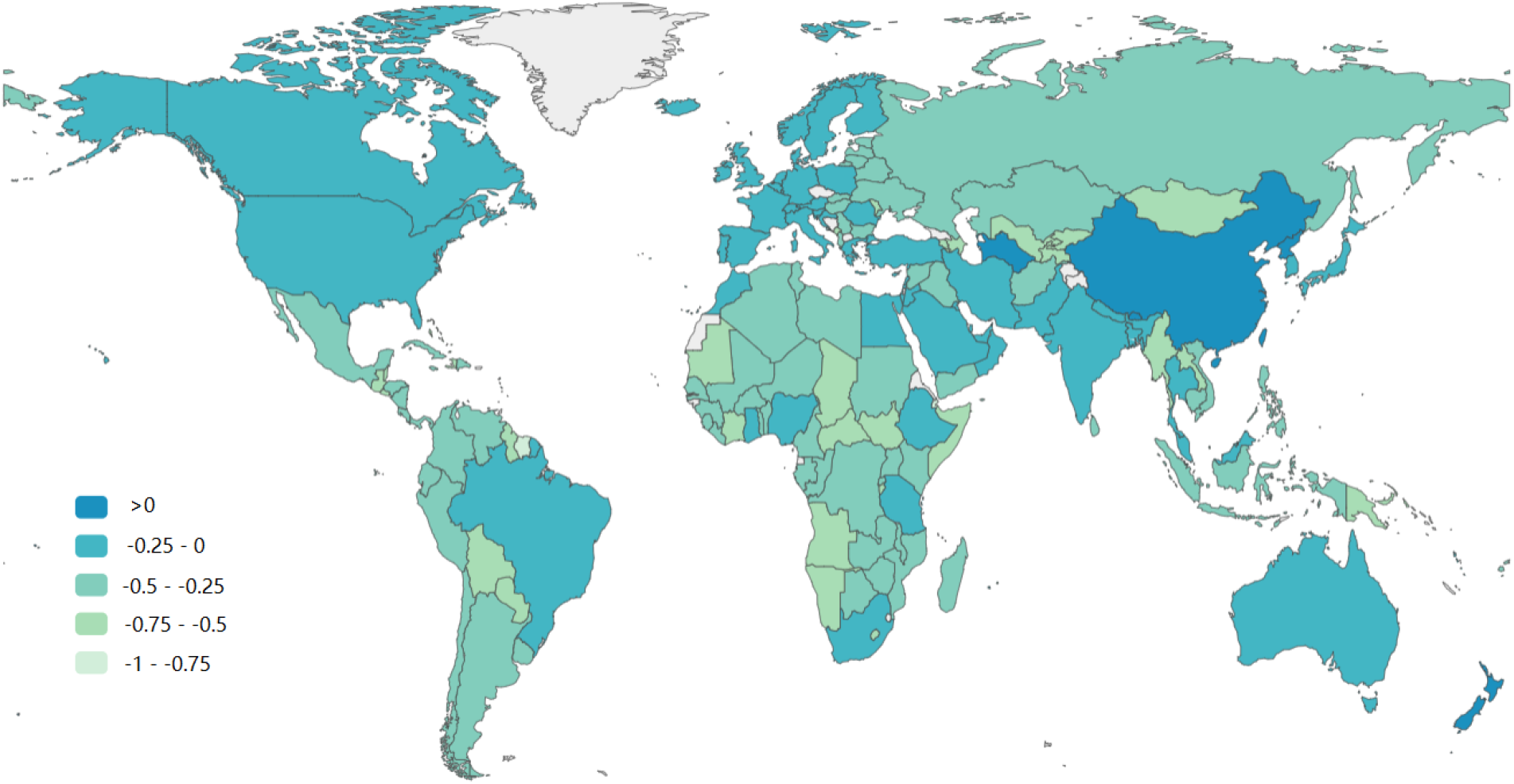
Correlation coefficient between publication number and death number in all the countries

**Table 31:** Correlation between publications and deaths in top-20 countries

| **Country** | **Num_pub**<br>$N$ | **Death numbers**<br>$I$ | $\rho_d$ | $\hat{\rho}_d$ |
| --- | --- | --- | --- | --- |
| US | 49,203 | 962,818 | -0.121 | 5.916 |
| China | 21,693 | 3,332 | 0.104 | -0.037 |
| Italy | 15,029 | 156,388 | -0.109 | 3.261 |
| UK | 14,475 | 161,507 | -0.112 | 3.264 |
| India | 13,044 | 508,086 | -0.162 | 4.153 |
| Spain | 6,372 | 101,733 | -0.137 | 2.415 |
| Canada | 6,093 | 37,207 | -0.094 | 1.678 |
| Germany | 5,857 | 124,772 | -0.15 | 2.511 |
| France | 5,196 | 140,229 | -0.161 | 2.52 |
| Brazil | 5,024 | 653,767 | -0.222 | 3.54 |
| Iran | 4,914 | 138,433 | -0.164 | 2.477 |
| Australia | 4,719 | 5,523 | -0.009 | 0.309 |
| Turkey | 4,600 | 95,954 | -0.153 | 2.195 |
| Japan | 3,678 | 25,530 | -0.106 | 1.246 |
| Saudi Arabia | 2,792 | 9,013 | -0.069 | 0.553 |
| South Korea | 2,696 | 9,646 | -0.075 | 0.586 |
| Netherlands | 2,123 | 21,742 | -0.132 | 0.98 |
| Poland | 2,012 | 113,002 | -0.209 | 1.829 |
| Switzerland | 1,880 | 13,364 | -0.115 | 0.693 |
| Pakistan | 1,797 | 30,291 | -0.159 | 1.097 |

The correlation between COVID-19 publications and deaths show that, of the top-20 publishing countries:

- China shows the highest log-correlation between their COVID-19 publication number and deaths, showing their highest productivity in the context of their deceased cases;
- Brazil shows the lowest log-correlation between their COVID-19 publication number and deaths, showing their lowest productivity in the context of their COVID-19 deaths.

### 8.6 Correlation between monthly disciplinary publications and infections

Here, we further analyze the correlation between the global publications and global infections for every month between Jan 2020 and Mar 2022 and the global publication-infection correlation in the three major disciplines: computer science, medical science, and social science, respectively.

The monthly publications are collected per the method introduced in Section 3.3 to group the publications to each month per their publication dates. We only count those publications with publication dates available. The categorization of monthly publications to the three major disciplines is done by the method introduced in Section 3.6. The monthly infections and deaths are collected per the method introduced in Section 3.9 globally.

Table 32 shows the number of the monthly global publications in computer science, medical science and social science and the number of global monthly new infection cases and deaths. It further shows the correlation coefficient between monthly publication number and infections and deaths, respectively.

**Table 32:**
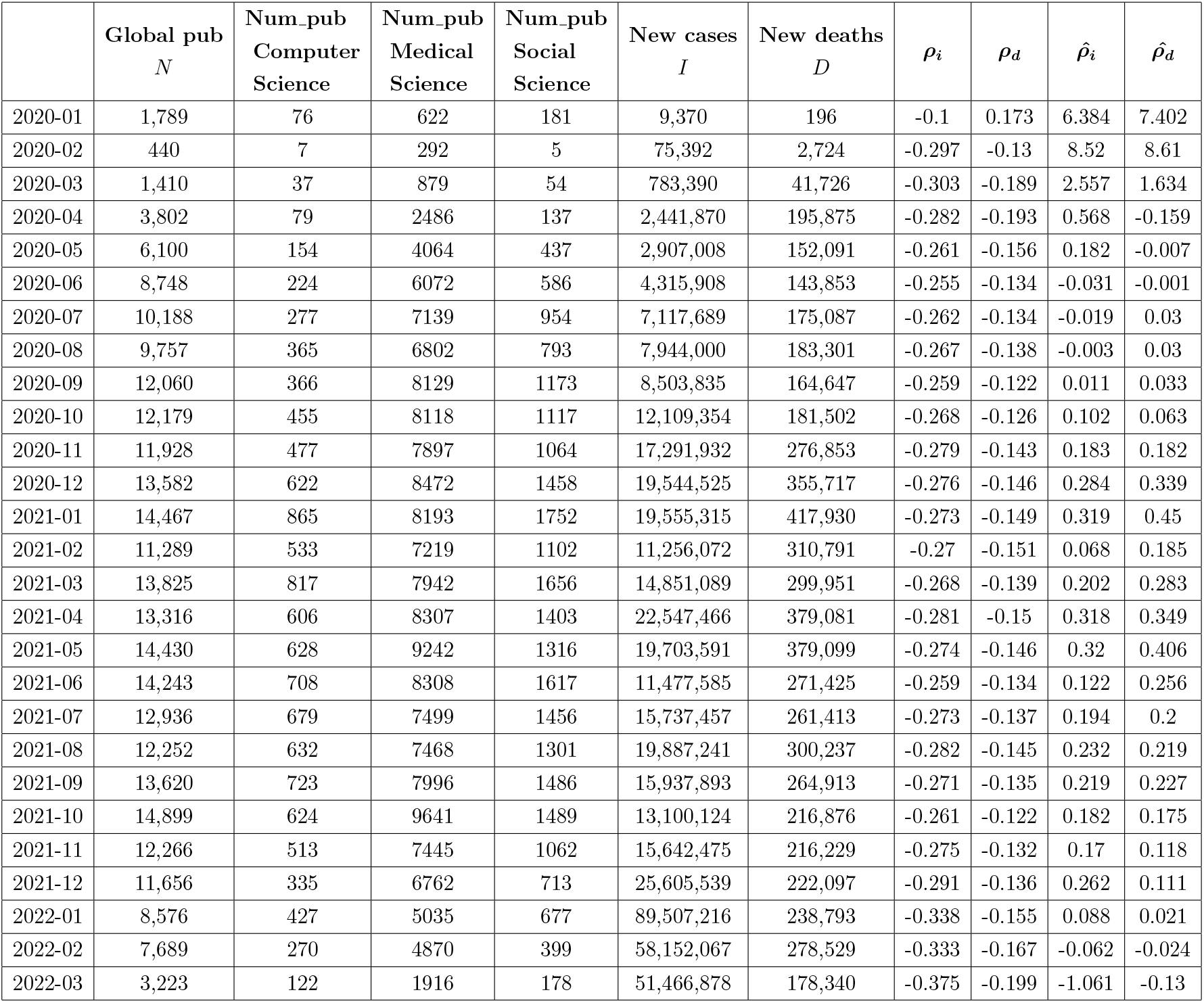
Monthly disciplinary publications and infection

| | Global pub<br>$N$ | Num_pub<br>Computer<br>Science | Num_pub<br>Medical<br>Science | Num_pub<br>Social<br>Science | New cases<br>$I$ | New deaths<br>$D$ | $\rho_i$ | $\rho_d$ | $\hat{\rho}_i$ | $\hat{\rho}_d$ |
| --- | --- | --- | --- | --- | --- | --- | --- | --- | --- | --- |
| 2020-01 | 1,789 | 76 | 622 | 181 | 9,370 | 196 | -0.1 | 0.173 | 6.384 | 7.402 |
| 2020-02 | 440 | 7 | 292 | 5 | 75,392 | 2,724 | -0.297 | -0.13 | 8.52 | 8.61 |
| 2020-03 | 1,410 | 37 | 879 | 54 | 783,390 | 41,726 | -0.303 | -0.189 | 2.557 | 1.634 |
| 2020-04 | 3,802 | 79 | 2486 | 137 | 2,441,870 | 195,875 | -0.282 | -0.193 | 0.568 | -0.159 |
| 2020-05 | 6,100 | 154 | 4064 | 437 | 2,907,008 | 152,091 | -0.261 | -0.156 | 0.182 | -0.007 |
| 2020-06 | 8,748 | 224 | 6072 | 586 | 4,315,908 | 143,853 | -0.255 | -0.134 | -0.031 | -0.001 |
| 2020-07 | 10,188 | 277 | 7139 | 954 | 7,117,689 | 175,087 | -0.262 | -0.134 | -0.019 | 0.03 |
| 2020-08 | 9,757 | 365 | 6802 | 793 | 7,944,000 | 183,301 | -0.267 | -0.138 | -0.003 | 0.03 |
| 2020-09 | 12,060 | 366 | 8129 | 1173 | 8,503,835 | 164,647 | -0.259 | -0.122 | 0.011 | 0.033 |
| 2020-10 | 12,179 | 455 | 8118 | 1117 | 12,109,354 | 181,502 | -0.268 | -0.126 | 0.102 | 0.063 |
| 2020-11 | 11,928 | 477 | 7897 | 1064 | 17,291,932 | 276,853 | -0.279 | -0.143 | 0.183 | 0.182 |
| 2020-12 | 13,582 | 622 | 8472 | 1458 | 19,544,525 | 355,717 | -0.276 | -0.146 | 0.284 | 0.339 |
| 2021-01 | 14,467 | 865 | 8193 | 1752 | 19,555,315 | 417,930 | -0.273 | -0.149 | 0.319 | 0.45 |
| 2021-02 | 11,289 | 533 | 7219 | 1102 | 11,256,072 | 310,791 | -0.27 | -0.151 | 0.068 | 0.185 |
| 2021-03 | 13,825 | 817 | 7942 | 1656 | 14,851,089 | 299,951 | -0.268 | -0.139 | 0.202 | 0.283 |
| 2021-04 | 13,316 | 606 | 8307 | 1403 | 22,547,466 | 379,081 | -0.281 | -0.15 | 0.318 | 0.349 |
| 2021-05 | 14,430 | 628 | 9242 | 1316 | 19,703,591 | 379,099 | -0.274 | -0.146 | 0.32 | 0.406 |
| 2021-06 | 14,243 | 708 | 8308 | 1617 | 11,477,585 | 271,425 | -0.259 | -0.134 | 0.122 | 0.256 |
| 2021-07 | 12,936 | 679 | 7499 | 1456 | 15,737,457 | 261,413 | -0.273 | -0.137 | 0.194 | 0.2 |
| 2021-08 | 12,252 | 632 | 7468 | 1301 | 19,887,241 | 300,237 | -0.282 | -0.145 | 0.232 | 0.219 |
| 2021-09 | 13,620 | 723 | 7996 | 1486 | 15,937,893 | 264,913 | -0.271 | -0.135 | 0.219 | 0.227 |
| 2021-10 | 14,899 | 624 | 9641 | 1489 | 13,100,124 | 216,876 | -0.261 | -0.122 | 0.182 | 0.175 |
| 2021-11 | 12,266 | 513 | 7445 | 1062 | 15,642,475 | 216,229 | -0.275 | -0.132 | 0.17 | 0.118 |
| 2021-12 | 11,656 | 335 | 6762 | 713 | 25,605,539 | 222,097 | -0.291 | -0.136 | 0.262 | 0.111 |
| 2022-01 | 8,576 | 427 | 5035 | 677 | 89,507,216 | 238,793 | -0.338 | -0.155 | 0.088 | 0.021 |
| 2022-02 | 7,689 | 270 | 4870 | 399 | 58,152,067 | 278,529 | -0.333 | -0.167 | -0.062 | -0.024 |
| 2022-03 | 3,223 | 122 | 1916 | 178 | 51,466,878 | 178,340 | -0.375 | -0.199 | -1.061 | -0.13 |

Figure 37 shows the correlation between each of the disciplinary publications and the infections and deaths over each month. The correlations between publications and infected and death numbers are calculated per the method introduced in Section 4.5.

**Figure 37:**
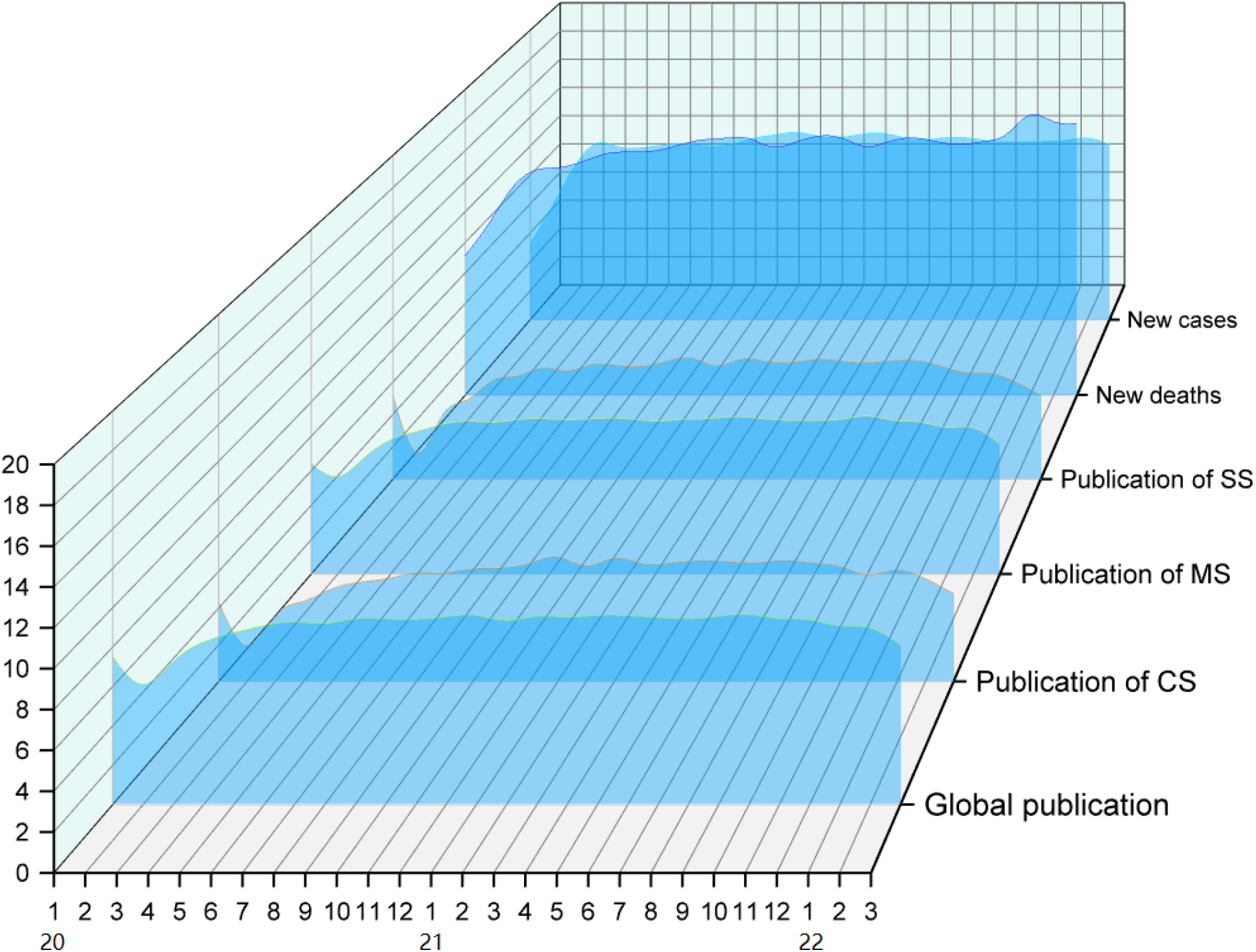
Correlation between monthly disciplinary publications and infections

The full details of the global monthly publications and infections are in the file “Correlation between monthly disciplinary publications and infections.xlsx.”

### 8.7 Correlation between G20 publications and infections

Table 33 shows the publications and case numbers in G20 countries and regions and their correlations with the publication number in each of their countries, respectively.

**Table 33:** Correlation between publications and infections in G20 countries

| Country | Num. pub<br>$N$ | Infection number<br>$I$ | Coefficient<br>$\rho_i$ | Dispersion Coefficient<br>$\hat{\rho}_i$ |
| --- | --- | --- | --- | --- |
| US | 49,203 | 79,477,350 | -0.255 | 3.547 |
| China | 21,693 | 128,930 | -0.082 | -4.098 |
| Italy | 15,029 | 13,159,490 | -0.26 | 0.404 |
| UK | 14,475 | 18,588,857 | -0.272 | 0.647 |
| India | 13,044 | 42,980,067 | -0.299 | 1.154 |
| Canada | 6,093 | 3,356,440 | -0.266 | -0.157 |
| Germany | 5,857 | 16,242,070 | -0.314 | 0.119 |
| France | 5,196 | 23,710,590 | -0.33 | 0.095 |
| Brazil | 5,024 | 29,093,019 | -0.337 | 0.084 |
| Australia | 4,719 | 3,525,635 | -0.281 | -0.018 |
| Turkey | 4,600 | 13,635,656 | -0.322 | 0.004 |
| Japan | 3,678 | 5,549,753 | -0.308 | 0.046 |
| Saudi Arabia | 2,792 | 748,121 | -0.261 | 0.732 |
| South Korea | 2,696 | 5,539,649 | -0.326 | 0.113 |
| Mexico | 1,436 | 5,583,993 | -0.362 | 0.243 |
| South Africa | 1,339 | 3,669,837 | -0.355 | 0.587 |
| Russia | 1,323 | 16,893,631 | -0.397 | -0.615 |
| Indonesia | 1,252 | 5,826,589 | -0.372 | 0.237 |
| Argentina | 590 | 8,955,458 | -0.43 | -0.187 |

The data can be found in ‘G20 publications and infections.xlsx.’

### 8.8 Correlation between G20 publications and deaths

Table 34 shows the publications and death case numbers in G20 countries and regions and their correlations in each of their countries, respectively.

**Table 34:** Correlation between publications and deaths in G20 countries

| Country | Num_pub<br>$N$ | Death number<br>$D$ | Coefficient<br>$\rho_d$ | Dispersion Coefficient<br>$\hat{\rho}_d$ |
| --- | --- | --- | --- | --- |
| US | 49,203 | 962,818 | -0.121 | 3.297 |
| China | 21,693 | 3,332 | 0.104 | -2.88 |
| Italy | 15,029 | 156,388 | -0.109 | 0.417 |
| UK | 14,475 | 161,507 | -0.112 | 0.425 |
| India | 13,044 | 508,086 | -0.162 | 1.075 |
| Canada | 6,093 | 37,207 | -0.094 | -0.137 |
| Germany | 5,857 | 124,772 | -0.15 | 0.056 |
| France | 5,196 | 140,229 | -0.161 | 0.038 |
| Brazil | 5,024 | 653,767 | -0.222 | 0.115 |
| Australia | 4,719 | 5,523 | -0.009 | -0.056 |
| Turkey | 4,600 | 95,954 | -0.153 | 0.001 |
| Japan | 3,678 | 25,530 | -0.106 | 0.146 |
| Saudi Arabia | 2,792 | 9,013 | -0.069 | 0.624 |
| South Korea | 2,696 | 9,646 | -0.075 | 0.648 |
| Mexico | 1,436 | 313,347 | -0.27 | -0.859 |
| South Africa | 1,339 | 99,333 | -0.23 | -0.111 |
| Russia | 1,323 | 351,124 | -0.28 | -1 |
| Indonesia | 1,252 | 151,135 | -0.251 | -0.425 |
| Argentina | 590 | 126,955 | -0.296 | -0.47 |

The data can be found in ‘G20 publications and deaths.xlsx.’

### 8.9 Correlation between OECD publications and infections

Table 35 shows the publications and case numbers in OECD countries and regions and their correlations with the publication number in each of their countries, respectively.

**Table 35:** Correlation between publications and infections in OECD countries

| Country | Num_pub<br>$N$ | Infection number<br>$I$ | Coefficient<br>$\rho_i$ | Dispersion Coefficient<br>$\hat{\rho}_i$ |
| --- | --- | --- | --- | --- |
| US | 49,203 | 79,477,350 | -0.255 | 5.699 |
| Italy | 15,029 | 13,159,490 | -0.26 | 1.801 |
| UK | 14,475 | 18,588,857 | -0.272 | 2.185 |
| Spain | 6,372 | 11,266,346 | -0.299 | 1.102 |
| Canada | 6,093 | 3,356,440 | -0.266 | 0.106 |
| Germany | 5,857 | 16,242,070 | -0.314 | 1.338 |
| France | 5,196 | 23,710,590 | -0.33 | 1.524 |
| Australia | 4,719 | 3,525,635 | -0.281 | 0.125 |
| Turkey | 4,600 | 13,635,656 | -0.322 | 1.037 |
| Japan | 3,678 | 5,549,753 | -0.308 | 0.366 |
| South Korea | 2,696 | 5,539,649 | -0.326 | 0.278 |
| Netherlands | 2,123 | 6,921,709 | -0.346 | 0.285 |
| Poland | 2,012 | 5,774,938 | -0.344 | 0.208 |
| Switzerland | 1,880 | 3,008,537 | -0.329 | 0.007 |
| Israel | 1,766 | 3,692,204 | -0.338 | 0.057 |
| Greece | 1,491 | 2,561,503 | -0.338 | -0.024 |
| Mexico | 1,436 | 5,583,993 | -0.362 | 0.102 |
| Belgium | 1,424 | 3,609,122 | -0.35 | 0.032 |
| Portugal | 1,173 | 3,485,075 | -0.361 | 0.012 |
| Ireland | 1,131 | 1,337,082 | -0.335 | -0.041 |
| Austria | 1,096 | 2,945,418 | -0.361 | 0 |
| Sweden | 1,067 | 2,462,952 | -0.357 | -0.005 |
| Denmark | 809 | 2,910,722 | -0.379 | 0.001 |
| Norway | 639 | 1,333,793 | -0.372 | 0.158 |
| New Zealand | 599 | 286,755 | -0.325 | 0.534 |
| Colombia | 584 | 6,074,155 | -0.421 | -0.175 |
| Chile | 574 | 3,230,793 | -0.405 | -0.024 |
| Finland | 435 | 703,052 | -0.378 | 0.531 |
| Czech Republic | 390 | 3,656,351 | -0.434 | -0.092 |
| Hungary | 298 | 1,809,917 | -0.433 | 0.261 |
| Slovenia | 206 | 909,863 | -0.44 | 0.823 |
| Slovakia | 164 | 2,231,352 | -0.482 | 0.221 |
| Lithuania | 108 | 1,016,341 | -0.493 | 1.048 |
| Luxembourg | 77 | 191,178 | -0.472 | 3.103 |
| Latvia | 67 | 719,813 | -0.523 | 1.683 |
| Estonia | 51 | 523,279 | -0.538 | 2.27 |
| Costa Rica | 44 | 820,445 | -0.563 | 1.76 |
| Iceland | 26 | 153,245 | -0.567 | 4.746 |

The data can be found in ‘OECD publications and infections.xlsx.’

### 8.10 Correlation between OECD publications and deaths

Here, we take the similar approach as in Section 8.5 to analyze the correlation between COVID-19 publications and the number of deaths in each of the G20 and OECD countries and regions.

Table 36 shows the numbers of publications and deaths and their correlation in G20 and OECD countries.

**Table 36:** Correlation between publications and deaths in OECD countries

| Country | Publication number<br>$N$ | Death number<br>$D$ | Coefficient<br>$\rho_d$ | Dispersion Coefficient<br>$\hat{\rho}_d$ |
| --- | --- | --- | --- | --- |
| US | 49,203 | 962,818 | -0.121 | 4.599 |
| Italy | 15,029 | 156,388 | -0.109 | 1.753 |
| UK | 14,475 | 161,507 | -0.112 | 1.754 |
| Spain | 6,372 | 101,733 | -0.137 | 0.963 |
| Canada | 6,093 | 37,207 | -0.094 | 0.408 |
| Germany | 5,857 | 124,772 | -0.15 | 1.025 |
| France | 5,196 | 140,229 | -0.161 | 1.012 |
| Australia | 4,719 | 5,523 | -0.009 | -0.517 |
| Turkey | 4,600 | 95,954 | -0.153 | 0.767 |
| Japan | 3,678 | 25,530 | -0.106 | 0.15 |
| South Korea | 2,696 | 9,646 | -0.075 | -0.168 |
| Netherlands | 2,123 | 21,742 | -0.132 | 0.051 |
| Poland | 2,012 | 113,002 | -0.209 | 0.384 |
| Switzerland | 1,880 | 13,364 | -0.115 | -0.047 |
| Israel | 1,766 | 10,330 | -0.106 | -0.085 |
| Greece | 1,491 | 26,366 | -0.164 | 0.049 |
| Mexico | 1,436 | 313,347 | -0.27 | 0.304 |
| Belgium | 1,424 | 30,430 | -0.174 | 0.058 |
| Portugal | 1,173 | 21,267 | -0.17 | 0.01 |
| Ireland | 1,131 | 6,597 | -0.111 | -0.033 |
| Austria | 1,096 | 18,195 | -0.167 | 0.001 |
| Sweden | 1,067 | 17,949 | -0.168 | 0.001 |
| Denmark | 809 | 4,954 | -0.119 | 0.079 |
| Norway | 639 | 1,755 | -0.072 | 0.303 |
| New Zealand | 599 | 65 | 0.208 | 0.844 |
| Colombia | 584 | 139,189 | -0.3 | -0.332 |
| Chile | 574 | 42,439 | -0.253 | -0.148 |
| Finland | 435 | 2,389 | -0.123 | 0.485 |
| Czech Republic | 390 | 39,062 | -0.278 | -0.227 |
| Hungary | 298 | 44,549 | -0.305 | -0.338 |
| Slovenia | 206 | 6,382 | -0.243 | 0.461 |
| Slovakia | 164 | 18,786 | -0.317 | -0.046 |
| Lithuania | 108 | 8,591 | -0.318 | 0.454 |
| Luxembourg | 77 | 1,002 | -0.227 | 2.136 |
| Latvia | 67 | 5,384 | -0.341 | 0.92 |
| Estonia | 51 | 2,329 | -0.325 | 1.741 |
| Costa Rica | 44 | 8,142 | -0.406 | 0.684 |
| Iceland | 26 | 77 | -0.139 | 5.737 |

The data can be found in ‘OECD publications and deaths.xlsx.’

## 9 Concluding remarks and discussion

In this report, we summarize our literature analyses of the global scientist’s responses to tackling COVID-19. We have collected about 350k scientific references on COVID-19 in English between Jan 2020 and Mar 2022, produced by researchers from 189 countries, including 175 first-authored countries, and in 27 subject areas.

Various literature analyses of this paramount global scientific effort have been made, including

- the descriptive statistics of the global publications in terms of publication quality and impact over countries and regions, subject areas and major disciplines, and main impact metrics;
- the profile and trends of COVID-19 publication quality and impact in G20 and OECD countries and regions;
- the publication profile, patterns and trends of publications on COVID-19 modeling;
- the correlations between COVID-19 publications and economic status and COVID-19 infections globally and in major countries and regions;
- the imbalances of COVID-19 research across subject areas, countries and regions, and quantity and quality.

The above analyses have been conducted on the information of title, abstract, author affiliations, and keywords extracted from global publications. This work complements our other comprehensive review on COVID-19 modeling in [**Cao-Liu’22**]. However, further work includes but is not limited to:

- more extraction and analyses of full text of these global publications;
- more analyses of the COVID-19 global scientists response data;
- specific review analyses of subject areas, for example, multi-disciplinary modeling of COVID-19, and modeling inference and impact of COVID-19;
- updating the publication impact analyses per the recently released 2021 impact metrics of journals and conferences.

## Data Availability

The data of global scientific response to COVID-19 is available at https://www.kaggle.com/datasets/datascienceslab/covid19-global-scientists-response and https://datasciences.org/covid19-modeling/.

## 10 Appendices

Here, we list a few appendices. We first include the domain-driven list of disciplinary categorization, the list of modeling keywords predefined by domain knowledge. Then, we introduce the collected metadata of COVID-19 publications, the list of files corresponding to the various results, and the list of files corresponding to source codes for processing the publications in this report.

### 10.1 Appendix 1. List of disciplinary categorization

Here, we collect the list of disciplinary keywords and terminologies collected from Web of Science, SJR, and Research.com, which are used to categorize the disciplines of each collected publication on COVID-19.

Table 37 shows the categories of subject areas in three major disciplines: medical science, computer science, and social science.

**Table 37:**
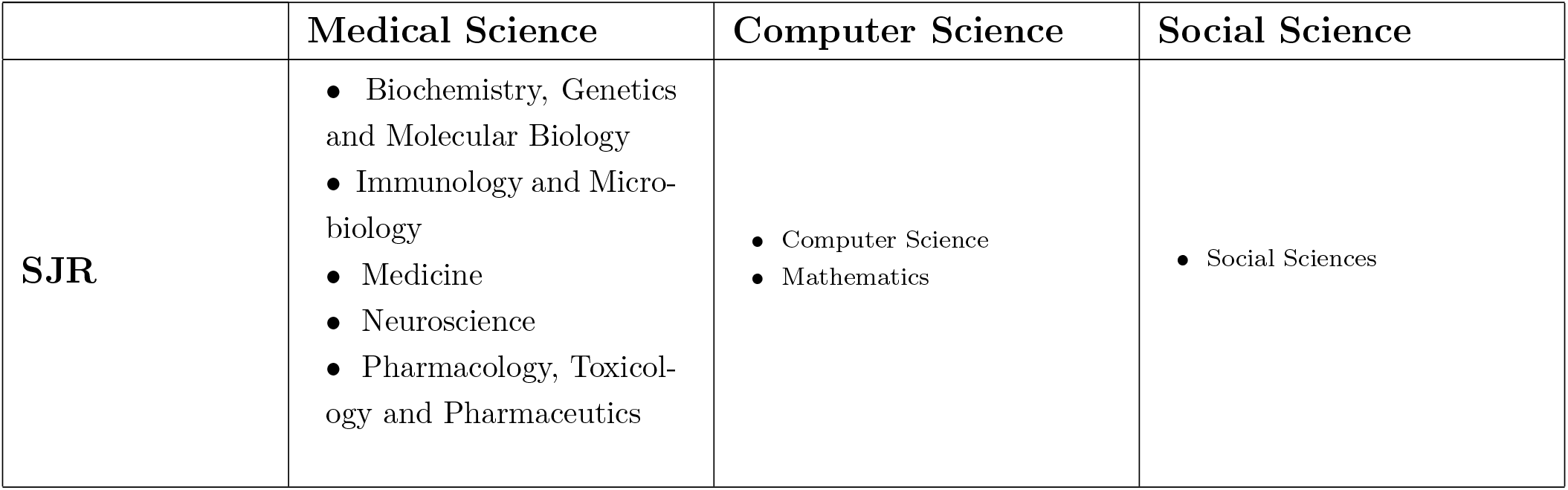
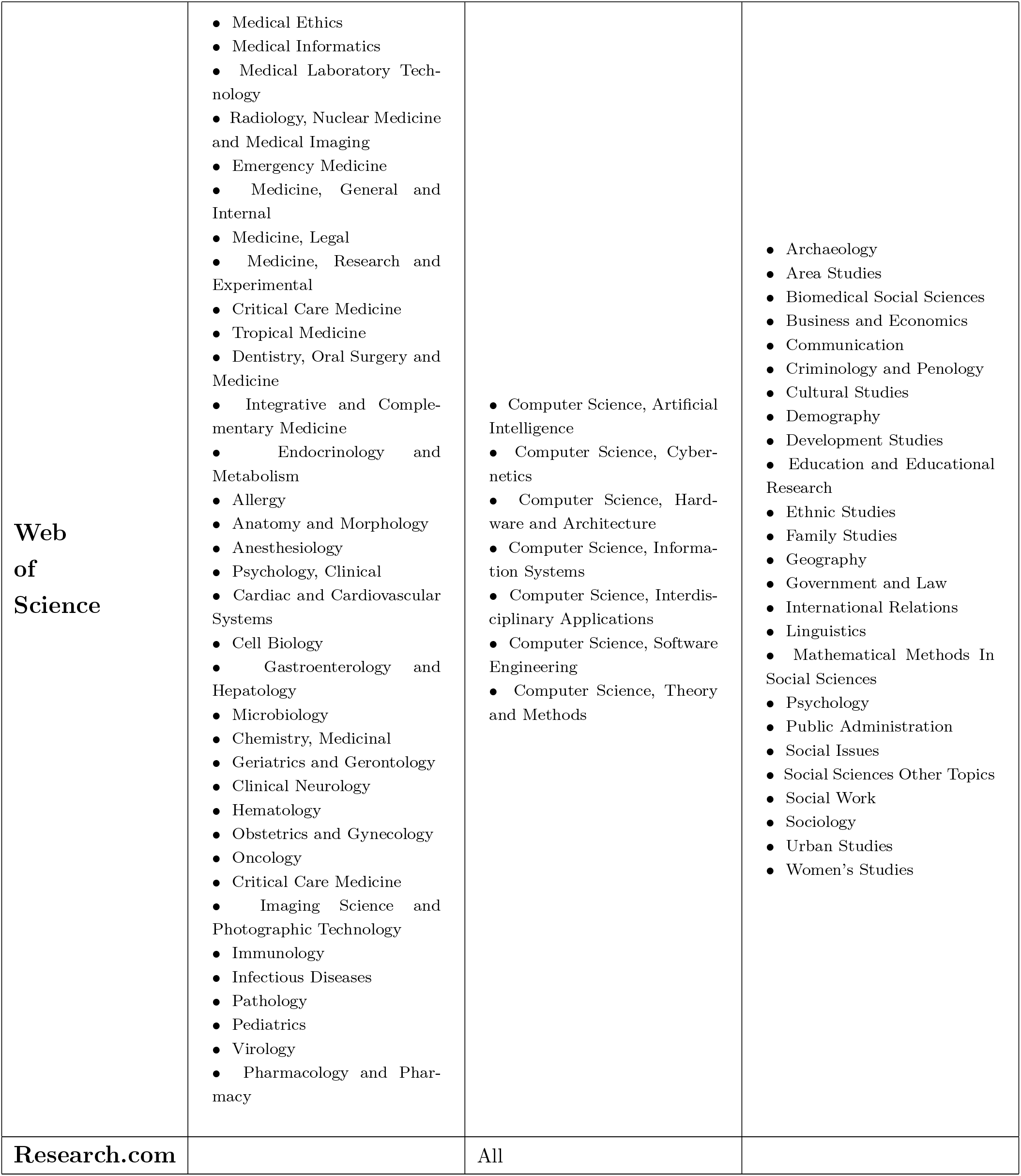
List of disciplinary categorization

|  | Medical Science | Computer Science | Social Science |
| --- | --- | --- | --- |
| **SJR** | • Biochemistry, Genetics and Molecular Biology<br>• Immunology and Microbiology<br>• Medicine<br>• Neuroscience<br>• Pharmacology, Toxicology and Pharmaceutics | • Computer Science<br>• Mathematics | • Social Sciences |
| **Web**<br>**of**<br>**Science** | • Medical Ethics<br>• Medical Informatics<br>• Medical Laboratory Technology<br>• Radiology, Nuclear Medicine and Medical Imaging<br>• Emergency Medicine<br>• Medicine, General and Internal<br>• Medicine, Legal<br>• Medicine, Research and Experimental<br>• Critical Care Medicine<br>• Tropical Medicine<br>• Dentistry, Oral Surgery and Medicine<br>• Integrative and Complementary Medicine<br>• Endocrinology and Metabolism<br>• Allergy<br>• Anatomy and Morphology<br>• Anesthesiology<br>• Psychology, Clinical<br>• Cardiac and Cardiovascular Systems<br>• Cell Biology<br>• Gastroenterology and Hepatology<br>• Microbiology<br>• Chemistry, Medicinal<br>• Geriatrics and Gerontology<br>• Clinical Neurology<br>• Hematology<br>• Obstetrics and Gynecology<br>• Oncology<br>• Critical Care Medicine<br>• Imaging Science and Photographic Technology<br>• Immunology<br>• Infectious Diseases<br>• Pathology<br>• Pediatrics<br>• Virology<br>• Pharmacology and Pharmacy | • Computer Science, Artificial Intelligence<br>• Computer Science, Cybernetics<br>• Computer Science, Hardware and Architecture<br>• Computer Science, Information Systems<br>• Computer Science, Interdisciplinary Applications<br>• Computer Science, Software Engineering<br>• Computer Science, Theory and Methods | • Archaeology<br>• Area Studies<br>• Biomedical Social Sciences<br>• Business and Economics<br>• Communication<br>• Criminology and Penology<br>• Cultural Studies<br>• Demography<br>• Development Studies<br>• Education and Educational Research<br>• Ethnic Studies<br>• Family Studies<br>• Geography<br>• Government and Law<br>• International Relations<br>• Linguistics<br>• Mathematical Methods In Social Sciences<br>• Psychology<br>• Public Administration<br>• Social Issues<br>• Social Sciences Other Topics<br>• Social Work<br>• Sociology<br>• Urban Studies<br>• Women's Studies |
| **Research.com** |  | All |  |

### 10.2 Appendix 2. List of predefined modeling keywords

The following includes the list of keywords we have collected from different sources and by our domain knowledge. They are categorized into machine learning, deep learning, mathematical modeling, epidemic modeling, and other general methods.

- Machine learning:
- Machine learning
- Sentiment analysis
- Support vector machine
- Transfer learning
- Random forest
- Decision tree
- Natural language processing
- Artificial neural network Deep learning:

Deep learning

- Convolutional neural network
- Neural network
- Long short-term memory
- Deep neural network

Mathematical modeling:

- Mathematical modeling
- Regression model linear regression
- Multivariate statistics
- Logistic regression
- Statistical model
- Bayesian
- Mathematical model
- Cox regression
- Univariate analysis
- Autoregressive integrated moving average
- Poisson regression
- Time series analysis
- Linear model
- Bivariate analysis
- Multiple linear regression
- Multivariate regression

Epidemic modeling:

- Compartmental modeling
- Susceptible-exposed-infectious-removed
- Susceptible-infectious-recovered
- Transmission model
- Epidemiological model

General and other methods and keywords:

- Compartmental modeling
- Predictive model
- Forecasting model
- Simulation
- Artificial intelligence
- Big data
- Decision making
- Data mining
- Data analytics

### 10.3 Appendix 3. List of COVID-19 publication metadata

Here, we list the metadata corresponding to the crawled publications and their supplementary information and impact metrics associated with each publication on COVID-19.

The metadata of this aggregated source data of COVID-19 publications are as follows in terms of reference information of publications, supplementary information of publications, and the impact metrics of the publication venues. There are 27 fields describing the COVID-19 publications. They are listed below.

- Pub_ID: the unique publication reference number;
- Pub_DOI: Publication DOI
- Title: Title of publication
- Abstract: Abstract of publication
- Authors: the authors of a publication
- First_author: First author of publication
- Author_affiliations: the affiliations of authors
- First_author_country: Country or region of the first author
- Coauthor_countries: the Countries of co-authors
- Pub_date: Publishing date of a publication
- Pub_source: Publication source
- Pub_source_type: Type of publication source
- Source_H5_format: H5-index format of publication source
- Source_Scopus_format: Scopus format of publication source
- Source_WOS_format: WoS format of publication source
- Keyword_from_pub: Keywords listed in the publication
- Keyword_by_OpenNLP: Keywords provided by OpenNLP
- Keyword_by_PositionRank: Keywords by PositionRank
- Source_discipline: Discipline or subject area of the publication
- Modeling_related: Is the publication related to AI, data science, machine learning, and other modeling methods?
- Modeling_technical_keywords: Modeling method keywords
- Source_Impact_Factor: Impact factor of the publication source
- Source_H5_Index: H5-index of the publication source
- Source_CiteScore: CiteScore of the publication source
- Source_SNIP: SNIP of the publication source
- Source_SJR: SJR of the publication source
- Source_Composite_Indicator: Composite Indicator of the publication source

Readers may also refer to the attributes of publication references from the following source:

- The COVID-19 Open Research Dataset (CORD-19) ([**Wang’20**])

The full publication metadata of this COVID-19 global scientist response dataset can be found in Kaggle at https://www.kaggle.com/datasets/datascienceslab/covid19-global-scientists-response.

### 10.4 Appendix 4. List of COVID-19 publication analysis results

Here, we list the files storing the analytical results of processing COVID-19 publications in the respective sections in this report.

The files are as follows:

- Correlation between monthly disciplinary publications and infections.xlsx
- G20 country’s overall research impact.xlsx
- G20 country’s research impact in computer science.xlsx
- G20 country’s research impact in medical science.xlsx
- G20 country’s research impact of social science.xlsx
- G20 paper-averaged research impact metrics.xlsx
- G20 publications and deaths.xlsx
- G20 publications and GDP per capita.xlsx
- G20 publications and infections.xlsx
- Global correlation between publications, infections and deaths.xlsx
- Global countries’ publications and GDP per capita.xlsx
- Global country’s research impact.xlsx
- Global disciplinary research impact.xlsx
- Global publication distribution.xlsx
- Global research collaborations.xlsx
- Global research impact distribution per Composite Indicator (CI).xlsx
- Global research impact distribution per SJR.xlsx
- Global research impact distribution per CiteScore.xlsx
- Global research impact distribution per H5-index.xlsx
- Global research impact distribution per Impact Factor.xlsx
- Global research impact distribution per SNIP.xlsx
- Mapping between top-50 business problems and their modelling methods.xlsx
- Modeling keyword cloud in modeling publications.xlsx
- Modeling word cloud of modeling publications from China.xlsx
- Modeling word cloud of modeling publications from the US.xlsx
- Modeling word cloud of modeling publications in computer science.xlsx
- Modeling word cloud of modeling publications in in social science.xlsx
- Modeling word cloud of modeling publications in medical science.xlsx
- Monthly top-10 business problems in modeling publications.xlsx
- OECD publications and deaths.xlsx
- OECD publications and GDP per capita.xlsx
- OECD publications and infections.xlsx
- Top-10 business problems globally in modeling publications.xlsx
- Top-10 modeling methods globally in modeling publications.xlsx
- Top-10 mostly published country’s disciplinary research impact.xlsx
- Word cloud of global publications.xlsx

### 10.5 Appendix 5. List of source codes for processing COVID-19 publications

- Here, we list the files storing the source codes of processing COVID-19 publications in the respective sections in this report.
- crawler/WOS_crawler.py,
- crawler/WHO_crawler.py,
- crawler/RG_crawler.py,
- crawler/PMD_keyword_crawler.py,
- crawler/medrxiv_crawler.py,
- crawler/crossref_crawler_by_id.py
- crawler/WOS_crawler.py,
- crawler/WHO_crawler.py,
- crawler/RG_crawler.py,
- crawler/PMD_keyword_crawler.py,
- crawler/medrxiv_crawler.py,
- crawler/crossref_crawler_by_id.py
- dataprocessing/pre process.py
- data_processing/country process.py
- data_processing/publish date process.py
- keyword_extraction/keyword position rank/main process.py
- data_processing/domain process.py
- data_processing/keyword_process.py
- data_processing/word cloud.py

## Footnotes

1 WHO Coronavirus (COVID-19) Dashboard: https://covid19.who.int/.

2 COVID-19 Coronavirus Pandemic: https://www.worldometers.info/coronavirus/#countries

3 We would suggest interested readers to read this technical report together with our systematic review on COVID-19 modeling in [**Cao-Liu’22, Covid19-modeling**].

4 https://opennlp.apache.org/

5 scimagojr.com

6 Research.com

7 https://www.ssrn.com/index.cfm/en/

8 The tool for creating world map is available at https://pyecharts.org/#/en-us/.

9 The tool for creating multi-dimensional visualization is available at https://www.originlab.com/.

10 The tool for creating radar chart is available at https://online.visual-paradigm.com/.

11 The tool for creating contour map is available at https://www.originlab.com/.

12 https://ai2-semanticscholar-cord-19.s3-us-west-2.amazonaws.com/historical_releases.html

13 Note, COVID-19 global literature on coronavirus disease between 2019 and 2023 is available at: https://search.bvsalud.org/global-literature-on-novel-coronavirus-2019-ncov/, this resource includes literature from different countries, languages, and disciplinary venues.

14 https://apps.webofknowledge.com

15 https://www.researchgate.net/

16 https://www.ncbi.nlm.nih.gov/pmc/ and https://pubmed.ncbi.nlm.nih.gov/

17 https://www.who.int/emergencies/diseases/novel-coronavirus-2019/global-research-on-novel-coronavirus-2019-ncov

18 https://www.crossref.org/

19 https://www.medrxiv.org/

20 https://arxiv.org/, since many publications here may not be associated with author country or publication date, we do not use the preprints from arXiv in this report.

21 https://scholar.google.com/citations?view_op=top_venues

22 https://clarivate.com/webofsciencegroup/solutions/journal-citation-reports/

23 https://www.scopus.com/sources.uri?RN_AG_Sourced_400000654

24 https://github.com/ymym3412/position-rank

25 https://github.com/allenai/scibert

26 scimagojr.com

27 Research.com

28 https://worldpopulationreview.com/

29 https://ourworldindata.org/covid-cases

30 https://scholar.google.com/citations?view_op=top_venues

31 https://clarivate.com/webofsciencegroup/essays/impact-factor/#:~:text=Thus%2C%20the%20impact%20factor%20of,(or%20total)%20citation%20frequencies.

32 https://jcr.clarivate.com/

33 https://www.elsevier.com/connect/editors-update/citescore-a-new-metric-to-help-you-choose-the-right-journal

34 https://www.scopus.com/sources

35 https://www.elsevier.com/connect/editors-update/citescore-a-new-metric-to-help-you-choose-the-right-journal

36 https://www.scopus.com/sources

37 https://www.elsevier.com/connect/editors-update/citescore-a-new-metric-to-help-you-choose-the-right-journal

38 https://www.scopus.com/sources

39 https://www.scimagojr.com/journalrank.php

40 Word cloud is a visual representation of words with greater prominence to those more frequently-occurring words.

41 The G20 countries and regions can be found in https://g20.org/about-the-g20/.

42 The OECD countries and regions can be found in https://www.oecd.org/about/members-and-partners/.

43 https://www.originlab.com/

